# Influence of Personality Traits on Mental Health, Coping, and Professional Performance in Medical Residents and Physicians: Systematic Review and Meta-Analysis

**DOI:** 10.64898/2026.05.11.26351770

**Authors:** Walaa Garoot, Edouard Leaune, Brice Canada, Christian Echevarría Colón, Marc Lilot, Gilles Rode, Sophie Schlatter

**Affiliations:** Faculté de Médecine Lyon Est, Lyon 1 Université, Lyon, France; Department of Anesthesiology and Intensive Care, Hospices Civils de Lyon, Lyon, France; Research on Healthcare Performance (RESHAPE), INSERM U1290, Lyon 1 Université Claude Bernard, Lyon, France; Healthcare Simulation Centre (SIMULYON, CLESS), Hospices Civils de Lyon - Lyon 1 Université, Lyon, France; Lyon 1 Université, CNRS, INSERM, Centre de Recherche en Neurosciences de Lyon CRNL U1028 UMR5292, TRAJECTOIRES, Bron, France; Laboratoire L-ViS / UCBL, INSAVALOR, 66 boulevard Niels Bohr, 69100 Villeurbanne, France; Carey Business School, Johns Hopkins University, Baltimore, MD, USA

**Keywords:** Big Five, Burnout, Coping, Five-Factor Model, Medical education, Medical students, Mental health, Personality, Physicians, Performance, Stress

## Abstract

**Background:** Medical residents and physicians face occupational stressors that increase the risk of mental health disorders and impaired professional functioning, with potential consequences for patient care. Although personality traits have been proposed as key determinants of individual adaptation to these demands, evidence on their relationship to mental health, coping, and performance has remained fragmented.

**Objective:** This systematic review and meta-analysis synthesize associations between personality traits and mental health, coping, and professional performance in medical residents and physicians.

**Methods:** PubMed, Embase, MEDLINE (Ovid), Cochrane Library, Scopus, and Web of Science were searched from inception to November 15, 2023, with an update in January 2026 (PROSPERO CRD42023483408). Eligible studies were English-language primary research involving medical residents and/or physicians, assessing at least one personality trait (FFM) and a validated measure of mental health (burnout and psychological distress), coping, or performance. Two reviewers independently screened records following PRISMA 2020 guidelines. Risk of bias was assessed using AXIS/JBI criteria and Cochrane Risk of Bias 2. Pearson r was pooled when 3 or more studies used the same instrument. Statistical significance was set at p < 0.05 (two-tailed). This review received no external funding.

**Results:** Of 4967 records identified, 34 studies (21,379 participants; 17 countries) were included. Neuroticism was associated with poorer mental health, including higher distress (r = 0.52; 95%CI: 0.18-0.87), emotional exhaustion (r = 0.42; 95% CI 0.22-0.62), and depersonalization (r = 0.30; 95% CI 0.17-0.44), and lower personal accomplishment (r = −0.24; 95%CI: −0.39 to −0.09; all k ≥ 3). Conscientiousness, Extraversion, and Agreeableness showed small-to-moderate protective associations with mental health (emotional exhaustion/depersonalization r = −0.18 to −0.36; personal accomplishment r = 0.20–0.25; all P ≤ .018). Openness showed weak and inconsistent associations across outcomes. Heterogeneity was substantial (I² > 75%), and risk of bias was low-to-moderate.

Narrative synthesis linked Neuroticism to disengagement-oriented coping and poorer performance, and Conscientiousness and Extraversion to problem-focused coping. Conscientiousness showed the most consistent positive association to performance.

**Conclusions:** Personality traits show modest but meaningful associations with mental health, coping, and professional performance in medical residents and physicians. Neuroticism appears as the main risk factor for poorer mental health, maladaptive coping, and worse professional performance. Conscientiousness, and to a lesser extent, Extraversion, and Agreeableness show small protective patterns. These findings support personality traits assessment as a formative resource for tailored support and professional development strategies in medical education and clinical practice.

## Introduction

Residents and physicians face high workloads, time pressure, emotionally demanding encounters, placing them at elevated risk of burnout, psychological distress, and impaired professional functioning.^1,2^ Identifying the factors that influence how individuals adapt to these demands is therefore essential for informing targeted support strategies and better addressing the needs of residents and physicians with different vulnerability and resilience profiles. Among these factors, personality traits have emerged as potentially important contributors to individual differences in adaptation to medical training and practice.^3–8^

The Five-Factor Model (FFM) describes five personality trait domains that capture major dimensions of individual behavioral differences. These include: Openness to experiences (a tendency to be imaginative, curious, and receptive to novel ideas), Conscientiousness (a tendency to be organized, responsible, and goal-directed), Extraversion (a tendency to be sociable, energetic, and assertive), Agreeableness (a tendency to be trustful, cooperative, and sympathetic), and Neuroticism (the inverse of Emotional Stability, reflecting a tendency to be anxious, insecure, and emotionally reactive).^9,10^ Because personality traits are relatively stable constructs that influence patterns of emotion, motivation, cognition, and behavior,^3^ they provide a useful framework for understanding individual differences in adaptation to the demands of medical practice.

Previous studies have linked personality traits to several dimensions of physician functioning, including mental health, coping, and professional performance. Higher Neuroticism, for example, has generally been associated with poorer mental health outcomes and greater use of disengagement or emotion-focused coping strategies, whereas Conscientiousness and Extraversion have tended to show more favorable associations across several outcomes.^1,2,4–7^ However, previous syntheses have largely examined these outcome domains separately and have predominantly included general occupational populations or heterogeneous groups of healthcare professionals rather than focusing specifically on medical residents and physicians.^1,11–17^ Consequently, despite substantial evidence linking personality to individual outcomes, the literature remains fragmented across outcome domains and populations, limiting an integrated understanding of how all five dimensions of the Five-Factor Model relate to physician functioning.

To address this gap, we conducted a systematic review and meta-analysis synthesizing evidence on the associations between FFM personality traits and physician functioning across three domains: (1) mental health outcomes, including burnout and psychological distress; (2) coping strategies; and (3) professional performance and professional skills, among medical residents and physicians.

## Methods

This systematic review and meta-analysis followed PRISMA 2020 recommendations;^18^ detailed in supplementary 1-3. The protocol was registered prospectively in PROSPERO (CRD42023483408; November 15, 2023; www.crd.york.ac.uk/prospero). AI technology was not used in any part of the study methodology. AI-assisted writing tools were used only during manuscript preparation and revision. Full details are provided in the dedicated Declaration of Generative AI and AI-Assisted Technologies section.

PubMed, Embase, MEDLINE (Ovid), Cochrane Library, Scopus, and Web of Science were searched from database inception to November 15, 2023. Last search update was in January 2026 without modification of eligibility criteria. Grey literature was sought through ProQuest and Google Scholar. Reference lists of included articles were screened manually. Studies were eligible if they: (1) included medical residents and/or practicing physicians; (2) assessed at least 1 personality trait using a recognized self-reported personality traits instrument (e.g., NEO inventories, Big Five Inventory); (3) reported a validated measure of mental health (e.g., psychophysiological stress), coping, and professional performance; and (4) were primary research with full text available in English. Studies were excluded if they involved exclusively students, did not assess FFM personality traits, did not relate personality or to a prespecified outcome, or had no accessible English full text after author contact.

Two reviewers (W.G. and S.S.) independently screened titles and abstracts using the Rayyan Systematic Review Platform (2023 edition) with blinding enabled, followed by independent full-text assessment. Disagreements were resolved through consensus. Data extraction was performed by one reviewer (W.G.) using a piloted standardized form and subsequently verified by S.S. Extracted variables included study design, sample size, professional level, medical specialty, age and sex distribution, personality assessment instrument, outcome measures, and reported effect estimates.

Risk of bias was assessed independently by two reviewers (W.G. and S.S.), using the AXIS tool (20 items) supplemented by 3 Joanna Briggs Institute analytical cross-sectional items for observational studies and the Cochrane Risk of Bias 2 tool for the randomized trial.^19–21^ Items were rated Yes (1), Partly (0.5), or No (0); study-level scores were categorized as low (≥0.75), moderate (0.50-0.74), or high risk (<0.50). Disagreements were resolved through discussion until consensus was reached. Inter-reviewer agreement was categorized as high (≥75%), moderate (50–74%), or low (<50%).

Pooling for meta-analysis was undertaken when three or more comparable studies used the same outcome instrument. Pearson r correlation coefficients was the primary effect size. When alternative statistics were reported (e.g., t, F, odds ratio, or standardized β coefficients), these were converted to r using established formulas. The resulting correlation coefficients were transformed to Fisher’s z for analysis and pooled using inverse-variance weighting under random-effects models with restricted maximum likelihood (REML) estimation. Pooled estimates and their 95% confidence intervals (95% CIs) were subsequently back-transformed and reported as Pearson’s r. Ninety-five percent prediction intervals (95% PIs) were also calculated to estimate the range within which the true effect of a future comparable study would be expected to fall, accounting for between-study heterogeneity. To aid interpretation, pooled correlations were categorized as trivial (r < 0.10), small (r ≈ 0.10–0.29), moderate (r ≈ 0.30–0.49), or large (r ≥ 0.50). All statistical tests were two-tailed, with statistical significance defined as p < .05.

Heterogeneity was quantified using Cochran’s Q, τ², and I², with I² values of approximately 25%, 50%, and 75% indicating low, moderate, and high heterogeneity, respectively. Heterogeneity was further explored through post hoc moderator analyses by experience level, region, and specialty; results are provided in Supplementary Materials 7–19. Publication bias was assessed using funnel plots and Egger regression test when k ≥ 10.^22–29^ Formal certainty of evidence assessment using GRADE was not undertaken, as this review synthesizes correlational associations rather than intervention effects, for which GRADE was not designed. All analyses were performed in JASP version 0.96.0.

## Results

### Study selection

The database search yielded 4967 records; 1747 duplicates were removed. Of 3220 records screened by title and abstract, 3076 were excluded as irrelevant. Of 144 full texts assessed, 127 were excluded—most commonly for exclusive focus on medical students (n = 45) or failure to assess FFM personality traits or prespecified outcomes (n = 82). Manual reference screening and the updated search to January 2026 identified 18 additional eligible studies. In total, 34 studies met inclusion criteria for qualitative synthesis. Twelve of them also met criteria for quantitative synthesis, of whom 11 contributed to the Maslach Burnout Inventory (MBI) meta-analyses and 3 to the General Health Questionnaire–12 (GHQ-12) meta-analysis, with 2 studies contributing to both (Figure 1).

**Figure 1.**
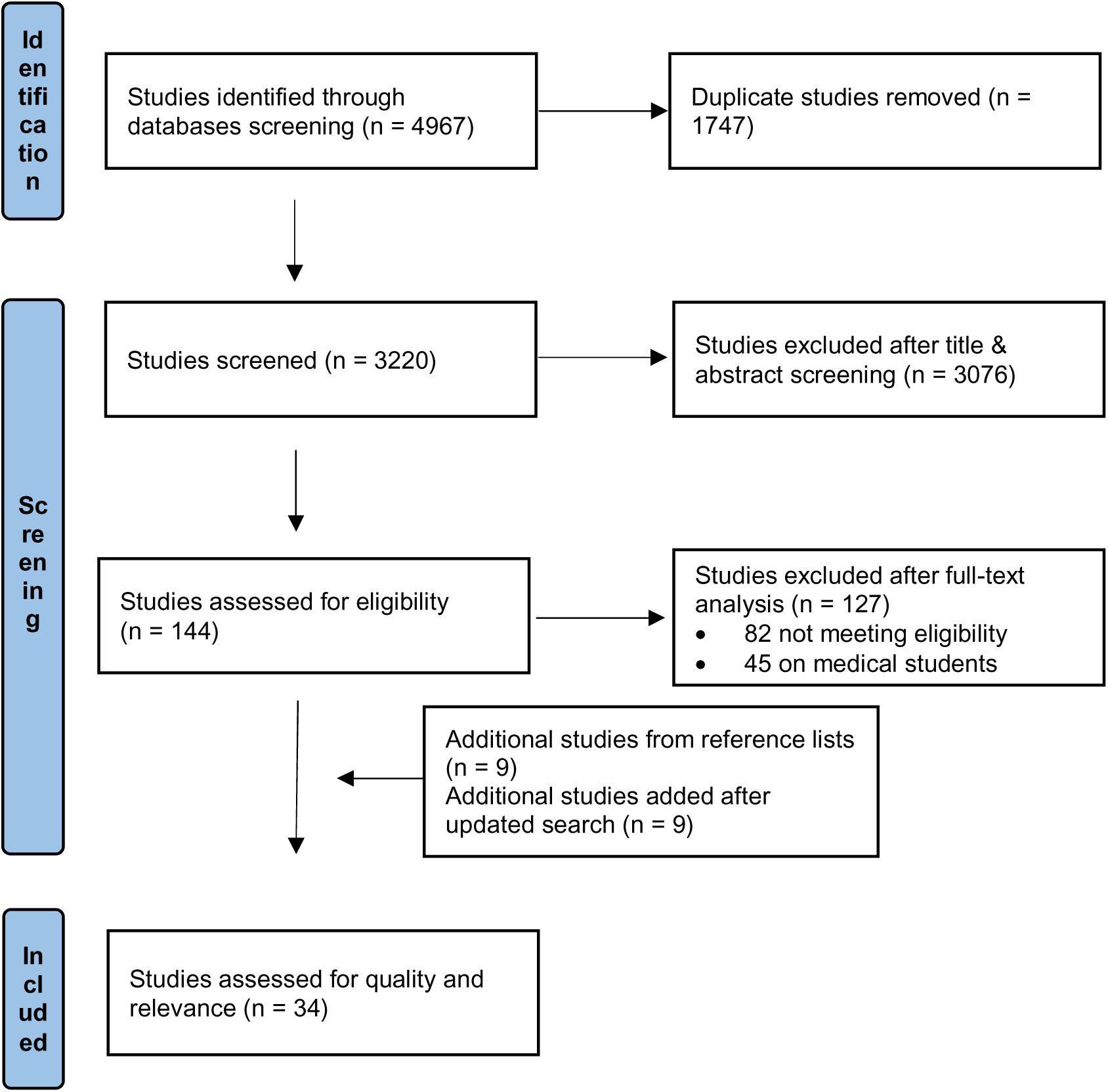
PRISMA 2020 flow diagram of study screening and selection

### Study characteristics

The 34 included studies enrolled 21,379 participants (published 1996-2025; 17 countries). Most originated from Europe (n = 18)^7,30–46^ followed by North America (n = 8)^47–54^ and Asia (n = 8)^55–62^; no eligible studies were identified from Africa, South America, or Oceania. Most were cross-sectional (n = 30); three had longitudinal components^32,53,54^ and one was a randomized trial.^7^ Sample sizes ranged from approximately 30 to more than 2000 participants. The study populations included both medical residents and fully qualified physicians; some studies focused solely on residents, others solely on attending physicians, and several combined both groups and did not distinguish them clearly. The most frequently represented specialties were surgery and related surgical fields (15 studies), internal medicine and other medical specialties (12 studies), psychiatry (9 studies), anesthesiology and critical care (7 studies), and pediatrics (6 studies). Age and gender were reported with variable details.

### Quality of the included studies

Across the 33 included observational studies, inter-rater agreement on AXIS/JBI items was greater than 75% for most studies. Quality scores ranged from 0.35 to 0.90; most fell in the low-to-moderate risk range (≥0.50); one study scored 0.36, indicating high risk. Core reporting quality and measurement validity were rated favorably; recurring concerns were incomplete adjustment for confounding, convenience sampling, and partial reporting of response rates and conflicts of interest. The randomized trial was rated “some concerns” on RoB 2. A detailed summary of individual AXIS/JBI appraisals and RoB2 is provided in Supplementary Methods 4-5.

### Quantitative and Qualitative Synthesis of the Outcomes

Personality traits were most assessed with the Big Five Inventory (n = 15 studies) or NEO measures (n = 10). The most frequently assessed outcomes were burnout (n = 16 studies) and psychological distress (n = 7). Study characteristics are presented in Table 1.

**Table 1.**

| Study ID | Country | Design | N | Personality instrument | Mental health outcomes & instruments | Coping outcomes & instruments | Professional performance outcomes & instruments |
| --- | --- | --- | --- | --- | --- | --- | --- |
| Deary IJ et al 1996<sup>33</sup> | Scotland | CSS | 189 | NEO-FFI (Only Neuroticism) | MBI; GHQ-28; SDSI | CISS | NA |
| McManus IC et al 2004<sup>32</sup> | United Kingdom | CSS + LS | 1,615 | BFI | MBI; GHQ-12 | NA | aAWQ; aWCQ; satisfaction with medicine |
| Duberstein P et al 2007<sup>52</sup> | USA | CSS | 96 | NEO-PI-R | NA | NA | PSQ |
| Lue BH et al 2010<sup>56</sup> | Taiwan | CSS | 555 | NEO-FFI | JSQ; C-CBI; PANAS | CSI | NA |
| Foulkrod KH et al 2010<sup>51</sup> | USA | CSS | 412 | IPIP | NA | NA | MSQ |
| Rosenthal R et al 2012<sup>38</sup> | Switzerland | CSS | 83 | NEO-FFI | NA | NA | VR laparoscopy simulator |
| Scheepers RA et al 2014<sup>37</sup> | Netherlands | CSS | 515 | BFI-10 | NA | NA | SETQ |
| Iorga M et al 2016<sup>31</sup> | Romania | CSS | 37 | BFI | MBI | NA | NA |
| **Kwarta P et al 2016<sup>41</sup>** | Poland | CSS | 50 | NEO-FFI | NA | CISS | NA |
| **Scheepers RA et al 2016<sup>36</sup>** | Netherlands | CSS | 515 | BFI-10 | NA | NA | SETQ; UWES-9 |
| **van der Wal R et al 2016<sup>39</sup>** | Netherlands | CSS | 655 | BFI (Dutch) | UBOS-C; GHQ-12 | NA | NA |
| **Iorga M et al 2017<sup>34</sup>** | Romania | CSS | 116 | BFI | MBI; self-reported depression | Self-reported pill use | NA |
| **Lindeman B et al 2017<sup>53</sup>** | USA | CSS + LS | 55 | TIPI | MBI (EE subscale) | NA | NA |
| **Sharma A et al 2017<sup>59</sup>** | India | CSS | 100 | BFI | MBI | NA | NA |
| **Mirzabeigi M et al 2018<sup>58</sup>** | Iran | CSS | 161 | NEO PI-R | MBI | NA | NA |
| **Brown PA et al 2019<sup>49</sup>** | Canada and Jamaica | CSS | 48 | BFI | MBI-HSS | NA | NA |
| **Hughes BD et al 2019<sup>48</sup>** | USA | CSS | 34 | TIPI | NA | NA | CAS; Grit scale |
| **Mullola S et al 2019<sup>45</sup>** | Finland | CSS | 2,815 | BFI-S | GHQ-12; suicidal ideation | NA | WAI |
| **Prins DJ et al 2019<sup>40</sup>** | Netherlands | CSS | 1,231 | BFI (Dutch) | UBOS-C | NA | NA |
| **Kheirkhah F et al 2020<sup>60</sup>** | Iran | CSS | 168 | NEO-FFI | NA | NA | SDM-Q-Doc |
| **Michelet D et al 2020<sup>46</sup>** | France | CSS | 48 | BFI-Fr | NA | NA | Customized technical performance checklist |
| **Gębska-Kuczerowska A et al 2021<sup>42</sup>** | Poland | CSS | 62 | NEO-FFI (Polish) | BOS | NA | NA |
| **Guo B et al 2021<sup>55</sup>** | China | CSS | 1,509 | NEO-PI | NA | NA | JPS; AMS; OCQ |
| **Gupta S et al 2021<sup>61</sup>** | India | CSS | 140 | NEO-FFI | NA | CSS-scale | GQ-6 |
| **Краснов E et al 2021<sup>30</sup>** | Russia | CSS | 122 | TIPI | NA | NA | JSE; ITE; TEIQue; REI; NTN-33 |
| **Martin SR et al 2022<sup>47</sup>** | USA | CSS | 69 | BFI | MBI | NA | BTPS |
| **Muntean LM et al 2022<sup>35</sup>** | Romania | CSS | 280 | DECAS | ASSET | NA | NA |
| **Schlatter S et al 2022<sup>7</sup>** | France | RCT | 147 | BFI-Fr | STAI-Y; cardiac reactivity score | Cardiac coherence score | NA |
| **Diaz E et al 2024<sup>43</sup>** | France | CSS | 1,020 | BFI-S | Wish to die | NA | NA |
| **Freeman B et al 2025<sup>50</sup>** | USA | CSS | 38 | IPIP | MBI-HSS | NA | NA |
| **Li Z et al 2025<sup>62</sup>** | China | CSS | 8,071 | CBF-PI-B (Neuroticism only) | MBI-HSS | CSS-scale | MSQ |
| **Morishita S et al 2025<sup>57</sup>** | Japan | CSS | 43 | TIPI-J | NA | NA | SDM-Q-9 |
| **Żak J et al 2025<sup>44</sup>** | Poland | CSS | 297 | FFM (50-item) | NA | NA | JSS |
| **Zastrow R et al 2025<sup>54</sup>** | USA | CSS + LS | 83 | BFP | aMBI; PSS | NA | NA |

#### 1. Mental health

• **Meta-analyses.** Eleven studies reporting all three Maslach Burnout Inventory dimensions (emotional exhaustion, depersonalization, and personal accomplishment) were included in the main burnout meta-analyses (55 effect sizes).^31,32,34,39,40,47,49,50,54,58,59^ Neuroticism showed the clearest and most consistent adverse pattern. Higher Neuroticism was associated with worst emotional exhaustion (pooled r = 0.42; 95%CI, 0.22-0.62; 95% prediction interval [PI], -0.23 to 1.06; P < .001), worst depersonalization (pooled r = 0.30; 95%CI, 0.17-0.44; 95% PI, -0.12 to 0.73; P < .001), and lower personal accomplishment (pooled r = -0.24; 95%CI, -0.39 to -0.09; 95% PI, -0.71 to 0.22; P = .005). Three GHQ-12 studies (10 effect sizes)^32,39,45^ showed that Neuroticism was associated with greater psychological distress (pooled r = 0.52; 95%CI, 0.18-0.87; 95% PI, -0.15 to 1.20; P = .02). Two prediction intervals exceeded the valid range for Pearson r (1.06 and 1.20), a known limitation of multilevel PI estimation reflecting high heterogeneity and a small number of studies; they are retained for transparency.

Conscientiousness, Extraversion, and Agreeableness each showed small-to-moderate, consistent protective patterns across burnout dimensions (emotional exhaustion and depersonalization: r = -0.181 to -0.364; 95%CI, -0.449 to -0.086; all P ≤ .003; personal accomplishment: r = 0.204 to 0.247; 95%CI, 0.049 to 0.413; all P ≤ .018).

Openness showed a weak, selective pattern: no significant association with emotional exhaustion (pooled r = -0.029; 95% CI, -0.091 to 0.034; P = .331), a small negative association with depersonalization (pooled r = -0.134; 95% CI, -0.248 to -0.020; P = .026), and a small positive association with personal accomplishment (pooled r = 0.168; 95%CI, 0.070 to 0.266; P = .003). Heterogeneity was substantial in all models (I^2^ > 75%). Funnel plot inspection and Egger test across burnout meta-analyses did not suggest clear evidence of small-study effects. Full qualitative and quantitative synthesis of the outcomes are presented in the Supplement 6. Full details of exploratory moderator analyses are presented in Supplementary Methods 7-19.

• **Narrative synthesis.** Narrative synthesis of studies not eligible for pooling was broadly concordant with the quantitative findings. Neuroticism consistently predicted higher burnout, while other traits showed modest and often inconsistent protective associations. Across studies examining psychological distress,^7,33–35,43,45,54,56^ Neuroticism was the most consistent vulnerability marker, higher Neuroticism correlate with higher distress, negative affectivity, and anticipatory stress. Conscientiousness, Extraversion, and Agreeableness showed small protective associations.

#### 2. Coping

Across studies examining coping (n = 7),^7,33,34,41,56,61,62^ Neuroticism consistently associated with emotion-focused and disengagement strategies. Conscientiousness and, to lesser extent, Extraversion were linked to problem-focused approaches. Agreeableness was associated with a prosocial coping profile. Openness showed minimal and direction-inconsistent associations.

#### 3. Professional performance

Across professional performance (n = 17 studies),^30,32,36–38,44–48,51,52,55,57,60–62^ including teaching performance, technical skills, job satisfaction, and non-technical aptitudes, Conscientiousness showed the most consistent positive pattern and Neuroticism the most consistent negative one, though findings were heterogeneous.

## Discussion

These findings provide a comprehensive synthesis of FFM personality trait associations with mental health, coping, and professional performance outcomes, in medical residents and physicians. Personality traits show modest but meaningful associations. Neuroticism emerged as the most consistent risk factor whereas Conscientiousness was the most consistent protective factor.

### 1. Mental health

Neuroticism showed the clearest and most consistent association with poorer mental health. Higher Neuroticism was associated with greater emotional exhaustion and depersonalization, lower personal accomplishment, and greater psychological distress. The magnitude and direction of these findings are broadly consistent with those reported in general occupational samples and other healthcare-specific reviews,^1,11–13^ supporting Neuroticism as a relatively consistent correlate of poorer occupational mental health across different occupational contexts. One possible explanation is that individuals higher in Neuroticism may be more likely to appraise workplace demands as threatening and to experience stronger negative emotional responses to stress.^12^

Conscientiousness showed smaller but consistently favorable associations across burnout dimensions, with lower emotional exhaustion and depersonalization and higher personal accomplishment. This pattern is consistent with the broader protective associations observed for Conscientiousness in occupational and healthcare populations.^11–13^ However, its association with broader psychological stress was not statistically significant, suggesting that its protective association may be more evident for specific dimensions of occupational burnout than for psychological distress more generally.

Extraversion and Agreeableness also showed small protective associations across the three burnout dimensions, although these associations were less pronounced than the adverse associations observed for Neuroticism. In particular, the association between Extraversion and personal accomplishment was weaker in our physician sample (r=0.20) than that previously reported in general working populations (r=0.36).^11–13^ Although this difference may suggest that the relevance of Extraversion varies across occupational contexts, direct comparisons between meta-analytic estimates should be interpreted cautiously given differences in study populations, measurement instruments, and study characteristics. Features of medical practice, including high workload, time pressure, and hierarchical or team-based working environments, could potentially modify the extent to which the interpersonal and positive-affect characteristics associated with Extraversion translate into greater personal accomplishment; however, this interpretation remains speculative.

Agreeableness showed its strongest favorable association with depersonalization, consistent with previous evidence suggesting that its interpersonal characteristics—such as cooperation, empathy, and positive interpersonal orientation—may be particularly relevant to this relational dimension of burnout.^11–13^

Openness showed the weakest and most selective pattern. It was not significantly associated with emotional exhaustion or broader psychological distress but was weakly associated with lower depersonalization and higher personal accomplishment. Rather than indicating a generalized favorable association with mental health, these findings suggest that Openness may relate selectively to particular dimensions of burnout. This pattern is broadly consistent with previous evidence showing relatively weak or inconsistent associations between Openness and occupational mental health outcomes.^11,12^

### 2. Coping

To our knowledge, this is the first systematic review to synthesize personality– coping associations specifically among medical residents and physicians. Examining these relationships may be particularly relevant in medicine, where physicians are repeatedly exposed to occupational stressors and coping strategies may influence how these demands are managed.^5^ Consistent with the aforementioned association between Neuroticism and poorer mental health outcomes, Neuroticism was associated predominantly with disengagement and emotion-focused coping strategies. This pattern is broadly consistent with findings in the general population, where Neuroticism has been associated with greater use of strategies such as withdrawal, wishful thinking, and emotion-focused coping.^14^ These findings raise the possibility that differences in coping may represent one pathway linking Neuroticism to adverse occupational outcomes; however, the available evidence does not establish a mediating or causal role of coping.

Conscientiousness showed the clearest adaptive coping pattern, with positive associations with problem-focused approaches. This is consistent with its organized and goal-directed characteristics and with evidence from the general population linking Conscientiousness particularly to greater use of problem-solving and cognitive restructuring strategies.^14^

Extraversion was also associated, although less consistently, with problem-focused coping, broadly aligning with general-population evidence linking Extraversion to problem-solving, cognitive restructuring, and support-seeking strategies.^14^ Agreeableness showed a weaker, predominantly prosocial coping profile, whereas associations with Openness were minimal and varied in direction across coping measures.^14^ These findings should nevertheless be interpreted in the context of the generally modest associations between personality and coping reported in the broader literature. The heterogeneity of coping constructs and measurement approaches across the studies included in this review may obscure trait-specific associations and limit direct comparisons across studies.

### 3. Professional performance

The relationship between personality and professional performance remains comparatively underexplored in medicine, and interpretation is complicated by substantial heterogeneity in how professional performance and related work outcomes were conceptualized and measured across the included studies which limits firm conclusions. This is particularly relevant given evidence from organizational research that personality–performance associations vary according to the performance criterion examined.^15^ Nevertheless, Conscientiousness showed the most consistently favorable pattern across performance, technical skills, and other work-related outcomes, whereas Neuroticism showed the most consistently unfavorable pattern. This pattern is consistent with evidence from broader occupational populations, in which Conscientiousness emerges as the strongest and most consistent Big Five correlate of job performance, whereas Neuroticism generally shows smaller unfavorable associations with performance.^15,17^

Extraversion and Agreeableness showed less consistent but generally favorable associations with selected professional and work-related outcomes, broadly compatible with evidence from broader occupational populations showing relatively small overall associations for these traits and suggesting that their relevance may vary according to the performance criterion and occupational context.^15,17^ Openness demonstrated the weakest and most inconsistent pattern, consistent with evidence suggesting comparatively limited associations with conventional job-performance criteria and a particularly weak association with overall job performance.^15,17^ A broadly similar trait pattern has also been reported for job satisfaction, with Neuroticism, Conscientiousness, and Extraversion showing stronger associations than Agreeableness and Openness.^16^ The broad direction of associations observed among physicians resembles that reported in general occupational populations. However, the heterogeneity of outcomes, measurement instruments, and study designs, together with the limited physician-specific evidence, precludes firm conclusions regarding the relative importance of individual personality traits for professional performance.^15–17^

## Strengths and limitations

This review has several notable strengths. It is the first systematic review and meta-analysis to examine the associations between FFM personality traits and mental health, coping, and performance outcomes among medical residents and physicians. By synthesizing data from 34 studies across 17 countries (21,379 participants), it provides a broad perspective across specialties, career stages, and nearly three decades of research that individual studies cannot achieve. Methodological rigor was maintained throughout the review process. Moreover, the convergence of meta-analytic and narrative findings across studies with diverse designs and assessment instruments enhances confidence in the consistency and robustness of the main conclusions.

However, several limitations must be acknowledged. As almost all included studies were observational, this meta-analyze estimates correlations rather than causal effects; this is inherent to personality research, where traits cannot be experimentally manipulated. Substantial heterogeneity limits precision, as two prediction intervals exceeded the mathematically valid range for Pearson r, a known artifact of multilevel prediction-interval estimation under extreme heterogeneity and small k. These intervals are retained for transparency but should not be interpreted as precise bounds. This review was restricted to English-language publications, which may have excluded eligible non-English studies and introduced a form of language bias not captured by our within-sample bias assessments. However, among the burnout meta-analyses (k ≥ 10), funnel-plot inspection and Egger’s test did not reveal clear evidence of small-study effects, though this assessment was not feasible for the psychological distress meta-analysis or for the narratively synthesized coping and performance outcomes.

## Implications for policy and practice

Implications for policy and practice include incorporating structured feedback based on personality profiles into medical trainees (undergraduate students and postgraduate residents) mentoring and/or and professional development programs, and shifting from reactive stress management toward proactive, personality-informed support.^2,5,63–65^ Neuroticism was the trait most consistently linked to maladaptive coping and worse performance outcomes, while Conscientiousness was most consistently linked to adaptive coping and better performance. These patterns point to two complementary targets for support—one at the individual level and one at the organizational level.

^•^ At the individual level, support may focus on strengthening emotional regulation, coping skills, planning, goal setting, and accountability according to individual needs. Occupational health follow-up may offer an additional setting to promote self-awareness of individual personality traits, metacognitive reflection on responses to stress, and access to tailored coping tools or additional psychological support when needed.^5–7^ More broadly, programs that normalize conversations about mental health and facilitate access to support may help medical residents and physicians with different personality profiles thrive without judgment.^2,5,63,64^

^•^ At the organizational level, personality-sensitive interventions are likely to be most effective when combined with structural changes to workload, supervision, and team climate. Structured educational programs in medical training have also shown the value of formal supervision, regular feedback, and longitudinal follow-up in supporting trainee engagement and development.^66^ Organization-directed interventions have been shown to produce meaningfully larger improvements in mental health than individual-directed interventions alone.^67,68^

## Future research

Further research is needed to clarify how personality traits relate to coping strategies and professional performance among physicians and medical residents, particularly given the limited and heterogeneous evidence in these domains. Larger longitudinal studies should examine whether personality traits predict changes in mental health, coping, and professional performance over time. Future research should also investigate sources of heterogeneity in these associations, including: specialty, training stage, workload, organizational culture, and other contextual factors, and evaluate whether personality-informed interventions can strengthen adaptive coping and resilience among individuals with different personality profiles.^5^ Future should prespecify subgroup analyses, improve adjustment for potential confounding factors, including: age, gender, workload, and organizational context, and transparently report funding sources and conflicts of interest. Studies from under-represented regions and specialties is also needed to assess the generalizability of these associations and inform context-appropriate interventions.

## Conclusions

Personality traits have modest but consistent associations with mental health, coping, and professional performance outcomes in medical residents and physicians. Neuroticism appears as the main risk factor for poorer mental health, maladaptive coping, and worse professional performance. Conscientiousness, and to lesser extent, Extraversion, and Agreeableness show small protective patterns. These findings support personality traits assessment as a formative resource within medical education and workforce development through targeted mentoring and call for structural organizational interventions alongside individual support.

## Declaration of Generative AI and AI-Assisted Technologies in the Writing Process

During manuscript preparation and revision, the authors used ChatGPT (OpenAI), Claude (Anthropic), Microsoft Copilot (Microsoft), and Paperpal (Cactus Communications) to support language refinement and clarification of selected concepts. ChatGPT and Claude were used to assist with clarification of concepts, sentence rephrasing, and improvements in clarity and coherence, while Microsoft Copilot and Paperpal were used primarily for grammar, language editing, and stylistic refinement. These tools were not treated as independent sources of scientific evidence or as autonomous decision-makers and were not used for study design, literature screening, data extraction, statistical analysis, selection or verification of results, or independent interpretation of the findings. All scientific decisions, citations, interpretations, and conclusions were made and critically reviewed by the authors. All AI-assisted text was subsequently reviewed and edited by the authors, who take full responsibility for the accuracy, integrity, and final content of the manuscript.

## Supporting information

Meta-analysis Output(7-19 Supp)

## Acknowledgments

The authors thank Steven Clocuh for his assistance with the study selection process.

## Data Availability

This is a systematic review and meta-analysis of previously published literature; no individual participant-level data were generated or collected by the authors. The extracted summary data and analysis code (JASP, version 0.96.0) supporting the results reported in this article are included in this published article and its supplementary files. Any additional materials are available from the W.G. and the corresponding author (S.S.) upon reasonable request.

## Author Contributions

Conceptualization: W.G. (lead), S.S. (equal)

Project administration: W.G. (lead), S.S. (equal)

Methodology: W.G. (lead), S.S. (equal), E.L. (supporting)

Data curation: W.G. (lead), S.S. (equal)

Investigation: W.G. (lead), S.S. (supporting)

Formal analysis: W.G (lead), C.EC. (equal), S.S. (supporting)

Supervision: E.L. (lead), B.C. (equal), M.L. (equal), G.R. (equal)

Writing – original draft: W.G. (lead), S.S. (supporting)

Writing – manuscript review & editing: W.G. (lead), S.S. (equal), E.L. (supporting), C.EC. (supporting), B.C. (supporting), M.L. (supporting), G.R. (supporting)

## Supplementary material

Appendix: Full PRISMA 2020 checklist (Supplementary Methods 1); PRISMA 2020 for Abstracts checklist (Supplementary Methods 2); complete PubMed search strategy (Supplementary Methods 3); AXIS/JBI and RoB2 risk-of-bias appraisals (Supplementary Methods 4-5); detailed qualitative and quantitative synthesis of the outcomes (Supplementary Methods 6); meta-analysis and exploratory moderator analyses results (Supplementary Methods 7-19).

