## Supplementary material for "Influence of Personality Traits on Mental Health, Coping, and Professional Performance in Medical Residents and Physicians: Systematic Review and Meta-Analysis": 1-6 Supp

### **Supplemental Methods 1. PRISMA 2020 Main Checklist**

| **Section and Topic** | **Item #** | **Checklist item** | **Location where item is reported** |
| --- | --- | --- | --- |
| **TITLE** | | |  |
| Title | 1 | Identify the report as a systematic review. | Title page |
| **ABSTRACT** | | |  |
| Abstract | 2 | See the PRISMA 2020 for Abstracts checklist. | Abstract section |
| **INTRODUCTION** | | |  |
| Rationale | 3 | Describe the rationale for the review in the context of existing knowledge. | Introduction |
| Objectives | 4 | Provide an explicit statement of the objective(s) or question(s) the review addresses. | Introduction |
| **METHODS** | | |  |
| Eligibility criteria | 5 | Specify the inclusion and exclusion criteria for the review and how studies were grouped for the syntheses. | Methods |
| Information sources | 6 | Specify all databases, registers, websites, organisations, reference lists and other sources searched or consulted to identify studies. Specify the date when each source was last searched or consulted. | Methods |
| Search strategy | 7 | Present the full search strategies for all databases, registers and websites, including any filters and limits used. | Supplementary Methods 3 |
| Selection process | 8 | Specify the methods used to decide whether a study met the inclusion criteria of the review, including how many reviewers screened each record and each report retrieved, whether they worked independently, and if applicable, details of automation tools used in the process. | Methods |
| Data collection process | 9 | Specify the methods used to collect data from reports, including how many reviewers collected data from each report, whether they worked independently, any processes for obtaining or confirming data from study investigators, and if applicable, details of automation tools used in the process. | Methods |
| Data items | 10a | List and define all outcomes for which data were sought. Specify whether all results that were compatible with each outcome domain in each study were sought (e.g. for all measures, time points, analyses), and if not, the methods used to decide which results to collect. | Methods |
|  | 10b | List and define all other variables for which data were sought (e.g. participant and intervention characteristics, funding sources). Describe any assumptions made about any missing or unclear information. | Methods |
| Study risk of bias assessment | 11 | Specify the methods used to assess risk of bias in the included studies, including details of the tool(s) used, how many reviewers assessed each study and whether they worked independently, and if applicable, details of automation tools used in the process. | Methods |
| Effect measures | 12 | Specify for each outcome the effect measure(s) (e.g. risk ratio, mean difference) used in the synthesis or presentation of results. | Methods |
| Synthesis methods | 13a | Describe the processes used to decide which studies were eligible for each synthesis (e.g. tabulating the study intervention characteristics and comparing against the planned groups for each synthesis (item #5)). | Methods |
|  | 13b | Describe any methods required to prepare the data for presentation or synthesis, such as handling of missing summary statistics, or data conversions. | Methods |
|  | 13c | Describe any methods used to tabulate or visually display results of individual studies and syntheses. | Methods; Forest plots in Supplement 7-19 |
|  | 13d | Describe any methods used to synthesize results and provide a rationale for the choice(s). If meta-analysis was performed, describe the model(s), method(s) to identify the presence and extent of statistical heterogeneity, and software package(s) used. | Methods |
|  | 13e | Describe any methods used to explore possible causes of heterogeneity among study results (e.g. subgroup analysis, meta-regression). | Methods + Results; Supplementary Methods 7-19 |
|  | 13f | Describe any sensitivity analyses conducted to assess robustness of the synthesized results. | Methods |
| Reporting bias assessment | 14 | Describe any methods used to assess risk of bias due to missing results in a synthesis (arising from reporting biases). | Methods |
| Certainty assessment | 15 | Describe any methods used to assess certainty (or confidence) in the body of evidence for an outcome. | Methods |
| **RESULTS** | | |  |
| Study selection | 16a | Describe the results of the search and selection process, from the number of records identified in the search to the number of studies included in the review, ideally using a flow diagram. | Results + Figure 1 |
|  | 16b | Cite studies that might appear to meet the inclusion criteria, but which were excluded, and explain why they were excluded. | Not applicable (no study was excluded if they meet the inclusion criteria) |
| Study characteristics | 17 | Cite each included study and present its characteristics. | Results + Table 1 |
| Risk of bias in studies | 18 | Present assessments of risk of bias for each included study. | Results; Supplementary Methods 4-5 |
| Results of individual studies | 19 | For all outcomes, present, for each study: (a) summary statistics for each group (where appropriate) and (b) an effect estimate and its precision (e.g. confidence/credible interval), ideally using structured tables or plots. | Results; Supplementary Methods 7-19 |
| Results of syntheses | 20a | For each synthesis, briefly summarise the characteristics and risk of bias among contributing studies. | Results |
|  | 20b | Present results of all statistical syntheses conducted. If meta-analysis was done, present for each the summary estimate and its precision (e.g. confidence/credible interval) and measures of statistical heterogeneity. If comparing groups, describe the direction of the effect. | Results |
|  | 20c | Present results of all investigations of possible causes of heterogeneity among study results. | Results; Supplementary Methods 7-19 |
|  | 20d | Present results of all sensitivity analyses conducted to assess the robustness of the synthesized results. | Not applicable |
| Reporting biases | 21 | Present assessments of risk of bias due to missing results (arising from reporting biases) for each synthesis assessed. | Results |
| Certainty of evidence | 22 | Present assessments of certainty (or confidence) in the body of evidence for each outcome assessed. | Not applicable (GRADE not applied; see Methods) |
| **DISCUSSION** | | |  |
| Discussion | 23a | Provide a general interpretation of the results in the context of other evidence. | Discussion |
|  | 23b | Discuss any limitations of the evidence included in the review. | Discussion |
|  | 23c | Discuss any limitations of the review processes used. | Discussion |
|  | 23d | Discuss implications of the results for practice, policy, and future research. | Discussion |
| **OTHER INFORMATION** | | |  |
| Registration and protocol | 24a | Provide registration information for the review, including register name and registration number, or state that the review was not registered. | Methods |
|  | 24b | Indicate where the review protocol can be accessed, or state that a protocol was not prepared. | Methods (PROSPERO CRD42023483408; www.crd.york.ac.uk/prospero) |
|  | 24c | Describe and explain any amendments to information provided at registration or in the protocol. | Methods |
| Support | 25 | Describe sources of financial or non-financial support for the review, and the role of the funders or sponsors in the review. | Funding Statement |
| Competing interests | 26 | Declare any competing interests of review authors. | Conflict of Interest section |
| Availability of data, code and other materials | 27 | Report which of the following are publicly available and where they can be found: template data collection forms; data extracted from included studies; data used for all analyses; analytic code; any other materials used in the review. | Data Availability |

### **Supplemental Methods 2. PRISMA 2020 Abstract Checklist**

| Section and topic | Item # | Checklist item | Location where item is reported |
| --- | --- | --- | --- |
| Title |  |  |  |
| Title | 1 | Identify the report as a systematic review. | Title page |
| Background |  |  |  |
| Objectives | 2 | Provide an explicit statement of the main objective(s) or question(s) the review addresses. | Objective |
| Methods |  |  |  |
| Eligibility criteria | 3 | Specify the inclusion and exclusion criteria for the review. | Methods |
| Information sources | 4 | Specify the information sources (e.g. databases, registers) used to identify studies and the date when each was last searched. | Methods |
| Risk of bias | 5 | Specify the methods used to assess risk of bias in the included studies. | Methods |
| Synthesis of results | 6 | Specify the methods used to present and synthesise results. | Methods |
| Results |  |  |  |
| Included studies | 7 | Give the total number of included studies and participants and summarise relevant characteristics of studies. | Results |
| Synthesis of results | 8 | Present results for main outcomes, preferably indicating the number of included studies and participants for each. If meta-analysis was done, report the summary estimate and confidence/credible interval. If comparing groups, indicate the direction of the effect (i.e. which group is favoured). | Results |
| Discussion |  |  |  |
| Limitations of evidence | 9 | Provide a brief summary of the limitations of the evidence included in the review (e.g. study risk of bias, inconsistency and imprecision). | Conclusions |
| Interpretation | 10 | Provide a general interpretation of the results and important implications. | Conclusions |
| Other |  |  |  |
| Funding | 11 | Specify the primary source of funding for the review. | Conclusions |
| Registration | 12 | Provide the register name and registration number. | Methods |

**Supplemental Methods 3. Details of the Search Strategy**

| **Key Concept/**  **Research Q** | **Concept 1**  personality traits/ psychophysiological stress/medical students and doctors | **Concept 2**  personality traits/coping skills/medical students and doctors | **Concept 3**  personality traits/ performances, professional skills, and aptitudes of medical students and doctors | **All concepts together** |
| --- | --- | --- | --- | --- |
| **Basic Search** | (“personality traits” OR ”big five” OR “big five personality traits“ OR “Big Five Inventory”) **AND** (stress OR “psychological stress” OR “physiological stress”) **AND** (“medical students” OR “medical trainee” OR doctors OR physician) | (“personality traits” OR ”big five” OR “big five personality traits“ OR “Big Five Inventory”) **AND** (“coping mechanisms” OR “coping skills” OR “stress management”) **AND** (“medical students” OR “medical trainee” OR doctors OR physician) | (“personality traits” OR ”big five” OR “big five personality traits“ OR “Big Five Inventory”) **AND** (performance OR “professional skills” OR “professional aptitudes”) **AND** (“medical students” OR “medical trainee” OR doctors OR physician) | (“personality traits” OR ”big five” OR “big five personality traits“ OR “Big Five Inventory”) **AND** (stress OR “psychological stress” OR “physiological stress”) **AND** (“coping mechanisms” OR “coping skills” OR “stress management”) **AND** (performance OR “professional skills” OR “professional aptitudes”) **AND** (“medical students” OR “medical trainee” OR doctors OR physician) |
| **Database Search Strategy** | | | | |
| **PubMed** | (("personality traits"[Title/Abstract] OR "big five"[Title/Abstract] OR "big five personality traits"[Title/Abstract]) AND (stress[Title/Abstract] OR "psychological stress"[Title/Abstract] OR "physiological stress"[Title/Abstract])) AND ("medical students"[Title/Abstract] OR "medical trainee"[Title/Abstract] OR doctors[Title/Abstract] OR physician[Title/Abstract]) | (("personality traits"[Title/Abstract] OR "big five"[Title/Abstract] OR "big five personality traits"[Title/Abstract]) AND ("coping mechanisms"[Title/Abstract] OR "coping skills"[Title/Abstract] OR "stress management"[Title/Abstract])) AND ("medical students"[Title/Abstract] OR "medical trainee"[Title/Abstract] OR doctors[Title/Abstract] OR physician[Title/Abstract]) | (("personality traits"[Title/Abstract] OR "big five"[Title/Abstract] OR "big five personality traits"[Title/Abstract]) AND (performance[Title/Abstract] OR "professional skills"[Title/Abstract] OR "professional aptitudes"[Title/Abstract])) AND ("medical students"[Title/Abstract] OR "medical trainee"[Title/Abstract] OR doctors[Title/Abstract] OR physician[Title/Abstract]) |  |
| **Embase** | ('personality traits' OR 'big five' OR 'big five personality traits') AND ('stress'/exp OR stress OR 'psychological stress'/exp OR 'psychological stress' OR 'physiological stress'/exp OR 'physiological stress') AND ('medical students'/exp OR 'medical students' OR 'medical trainee' OR doctors OR 'physician'/exp OR physician) | ('personality traits' OR 'big five' OR 'big five personality traits' OR 'big five inventory'/exp OR 'big five inventory') AND ('coping mechanisms' OR 'coping skills' OR 'stress management'/exp OR 'stress management') AND ('medical students'/exp OR 'medical students' OR 'medical trainee' OR doctors OR 'physician'/exp OR physician) | ('personality traits' OR 'big five' OR 'big five personality traits' OR 'big five inventory'/exp OR 'big five inventory') AND ('performance'/exp OR performance OR 'professional skills' OR 'professional aptitudes') AND ('medical students'/exp OR 'medical students' OR 'medical trainee' OR doctors OR 'physician'/exp OR physician) | ('medical student'/exp OR 'med school student' OR 'med student' OR 'medical school student' OR 'medical student' OR 'medical students' OR 'student, medical' OR 'students, medical' OR 'physician'/exp OR 'doctor' OR 'medical doctor' OR 'medical practitioner' OR 'physician' OR 'physician associate' OR 'physicians' OR 'practitioner' OR 'private physician' OR 'medical trainee') AND ('personality'/exp OR 'personality' OR 'personality characteristic' OR 'personality pattern' OR 'personality structure' OR 'personality type' OR 'psychologic structure') AND (stress OR 'coping behavior'/exp OR 'behavior, coping' OR 'behavior, coping' OR 'coping' OR 'coping ability' OR 'coping behavior' OR 'coping behavior' OR 'coping mechanism' OR 'coping strategy' OR 'coping style' OR 'performance'/exp OR 'performance' OR 'performance test' OR 'progressive ratio performance' OR 'professional skills' OR 'professional aptitudes') |
| **Cochrane Library [Cochrane Central Register of Controlled Trials (CENTRAL)]** | (#1 OR #2) AND (#3 OR #4 OR #5) AND (#10 OR #11 OR #12)"  #1 MeSH descriptor: [Personality Tests] explode all trees  #2 ((“personality traits” OR ”big five” OR “big five personality traits“ OR “Big Five Inventory”)):ti,ab,kw  #3 MeSH descriptor: [Stress, Psychological] explode all trees  #4 MeSH descriptor: [Stress, Physiological] explode all trees  #5 ((stress OR “psychological stress” OR “physiological stress”)):ti,ab,kw  #10 MeSH descriptor: [Students, Medical] explode all trees  #11 MeSH descriptor: [Physicians] explode all trees  #12 ((“medical students” OR “medical trainee” OR doctors OR physician)):ti,ab,kw | (#1 OR #2) AND (#6 OR #7) AND (#10 OR #11 OR #12)  #1 MeSH descriptor: [Personality Tests] explode all trees  #2 ((“personality traits” OR ”big five” OR “big five personality traits“ OR “Big Five Inventory”)):ti,ab,kw  #6 MeSH descriptor: [Adaptation, Psychological] explode all trees  #7 ((“coping mechanisms” OR “coping skills” OR “stress management”)):ti,ab,kw  #10 MeSH descriptor: [Students, Medical] explode all trees  #11 MeSH descriptor: [Physicians] explode all trees  #12 ((“medical students” OR “medical trainee” OR doctors OR physician)):ti,ab,kw | (#1 OR #2) AND (#8 OR #9) AND (#10 OR #11 OR #12)  #1 MeSH descriptor: [Personality Tests] explode all trees  #2 ((“personality traits” OR ”big five” OR “big five personality traits“ OR “Big Five Inventory”)):ti,ab,kw  #8 MeSH descriptor: [Work Performance] explode all trees  #9 ((performance OR “professional skills” OR “professional aptitudes”)):ti,ab,kw  #10 MeSH descriptor: [Students, Medical] explode all trees  #11 MeSH descriptor: [Physicians] explode all trees  #12 ((“medical students” OR “medical trainee” OR doctors OR physician)):ti,ab,kw | (#1 OR #2) AND (#3 OR #4 OR #5) AND (#6 OR #7) AND (#8 OR #9) AND (#10 OR #11 OR #12) |
| **MEDLINE (Ovid)** | - **Basic search:**   personality traits/ psychophysiological stress/medical students and doctors {Including Related Terms}   - **Advanced search:** Personality/ or Personality Inventory/ AND Stress, Psychological/ or Stress, Physiological/ AND (medical students and doctors).mp. [mp=title, book title, abstract, original title, name of substance word, subject heading word, floating sub-heading word, keyword heading word, organism supplementary concept word, protocol supplementary concept word, rare disease supplementary concept word, unique identifier, synonyms, population supplementary concept word, anatomy supplementary concept word] | - **Basic search:**   personality traits/coping skills/medical students and doctors {Including Related Terms}   - **Advanced search:** Personality/ or Personality Inventory/ AND "behavior and behavior mechanisms"/ AND (medical students and doctors).mp. [mp=title, book title, abstract, original title, name of substance word, subject heading word, floating sub-heading word, keyword heading word, organism supplementary concept word, protocol supplementary concept word, rare disease supplementary concept word, unique identifier, synonyms, population supplementary concept word, anatomy supplementary concept word] | - **Basic search:**   personality traits/ performances, professional skills, and aptitudes of medical students and doctors {Including Related Terms}   - **Advanced search:** Personality/ or Personality Inventory/ AND (performances, professional skills and aptitudes).mp. [mp=title, book title, abstract, original title, name of substance word, subject heading word, floating sub-heading word, keyword heading word, organism supplementary concept word, protocol supplementary concept word, rare disease supplementary concept word, unique identifier, synonyms, population supplementary concept word, anatomy supplementary concept word] AND (medical students and doctors).mp. [mp=title, book title, abstract, original title, name of substance word, subject heading word, floating sub-heading word, keyword heading word, organism supplementary concept word, protocol supplementary concept word, rare disease supplementary concept word, unique identifier, synonyms, population supplementary concept word, anatomy supplementary concept word] |  |
| **Scopus** | ( TITLE-ABS-KEY ( "personality traits" OR "big five" OR "big five personality traits" OR "Big Five Inventory" ) AND TITLE-ABS-KEY ( stress OR "psychological stress" OR "physiological stress" ) AND TITLE-ABS-KEY ( "medical students" OR "medical trainee" OR doctors OR physician ) ) | ( TITLE-ABS-KEY ( "personality traits" OR "big five" OR "big five personality traits" OR "Big Five Inventory" ) AND TITLE-ABS-KEY ( "coping mechanisms" OR "coping skills" OR "stress management" ) AND TITLE-ABS-KEY ( "medical students" OR "medical trainee" OR doctors OR physician ) ) | ( TITLE-ABS-KEY ( "personality traits" OR "big five" OR "big five personality traits" OR "Big Five Inventory" ) AND TITLE-ABS-KEY ( performance OR "professional skills" OR "professional aptitudes" ) AND TITLE-ABS-KEY ( "medical students" OR "medical trainee" OR doctors OR physician ) ) |  |
| **Web of Science** | “personality traits” OR ”big five” OR “big five personality traits“ OR “Big Five Inventory” (Topic) and (stress OR “psychological stress” OR “physiological stress”) (Topic) and (“medical students” OR “medical trainee” OR doctors OR physician) (Topic) | “personality traits” OR ”big five” OR “big five personality traits“ OR “Big Five Inventory” (Topic) and (“coping mechanisms” OR “coping skills” OR “stress management”) (Topic) and (“medical students” OR “medical trainee” OR doctors OR physician) (Topic) | “personality traits” OR ”big five” OR “big five personality traits“ OR “Big Five Inventory” (Topic) and (performance OR “professional skills” OR “professional aptitudes”) (Topic) and (“medical students” OR “medical trainee” OR doctors OR physician) (Topic) | “personality traits” OR ”big five” OR “big five personality traits“ OR “Big Five Inventory” (Topic) and (stress OR “psychological stress” OR “physiological stress”) (Topic) and (“coping mechanisms” OR “coping skills” OR “stress management”) (Topic) and (performance OR “professional skills” OR “professional aptitudes”) (Topic) and (“medical students” OR “medical trainee” OR doctors OR physician) (Topic) |
| **ProQuest** | (("personality traits" OR "big five" OR "big five personality traits" OR "Big Five Inventory") AND (stress OR "psychological stress" OR "physiological stress") AND ("medical students" OR "medical trainee" OR doctors OR physician)) AND la.exact("ENG") | (("personality traits" OR "big five" OR "big five personality traits" OR "Big Five Inventory") AND ("coping mechanisms" OR "coping skills" OR "stress management" OR "coping strategies" OR BCI OR cope) AND ("medical students" OR "medical trainee" OR doctors OR physician)) AND la.exact("ENG") | (("personality traits" OR "big five" OR "big five personality traits" OR "Big Five Inventory") AND (performance OR "professional skills" OR "professional aptitudes") AND ("medical students" OR "medical trainee" OR doctors OR physician)) AND la.exact("ENG") | ((("personality traits" OR "big five" OR "big five personality traits" OR"Big Five Inventory") AND (performance OR "professional skills" OR "professional aptitudes") AND ("medical students" OR "medical trainee" OR doctors OR physician)) AND la.exact("ENG")) AND ((("personality traits" OR "big five" OR "big five personality traits" OR "Big Five Inventory") AND ("coping mechanisms" OR "coping skills" OR "stress management" OR "coping strategies" OR BCI OR cope) AND ("medical students" OR "medical trainee" OR doctors OR physician)) AND la.exact("ENG")) AND ((("personality traits" OR "big five" OR "big five personality traits" OR "Big Five Inventory") AND (stress OR "psychological stress" OR "physiological stress") AND ("medical students" OR "medical trainee" OR doctors OR physician)) AND la.exact("ENG")) |

**Supplemental Methods 4. AXIS - JBI**

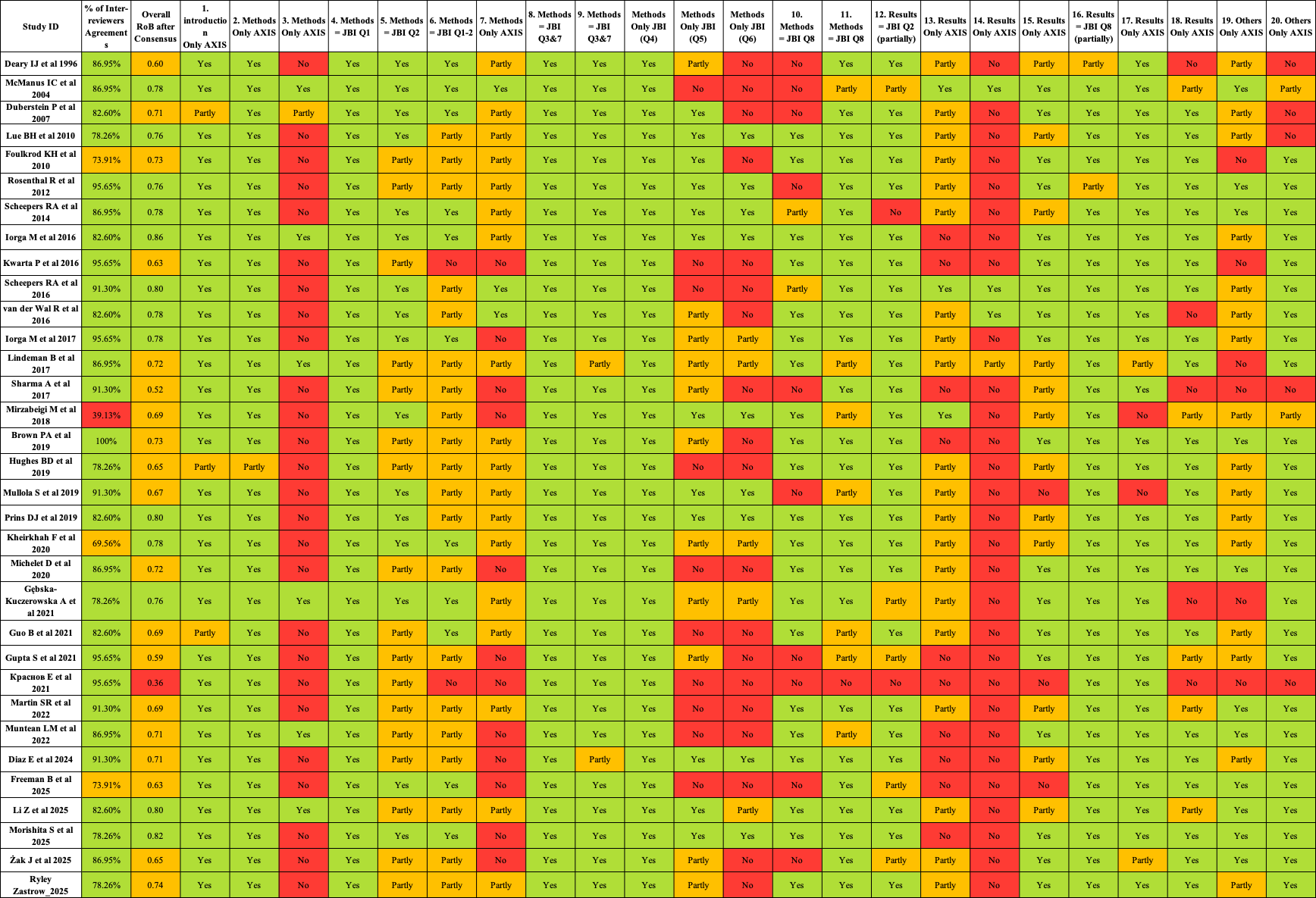

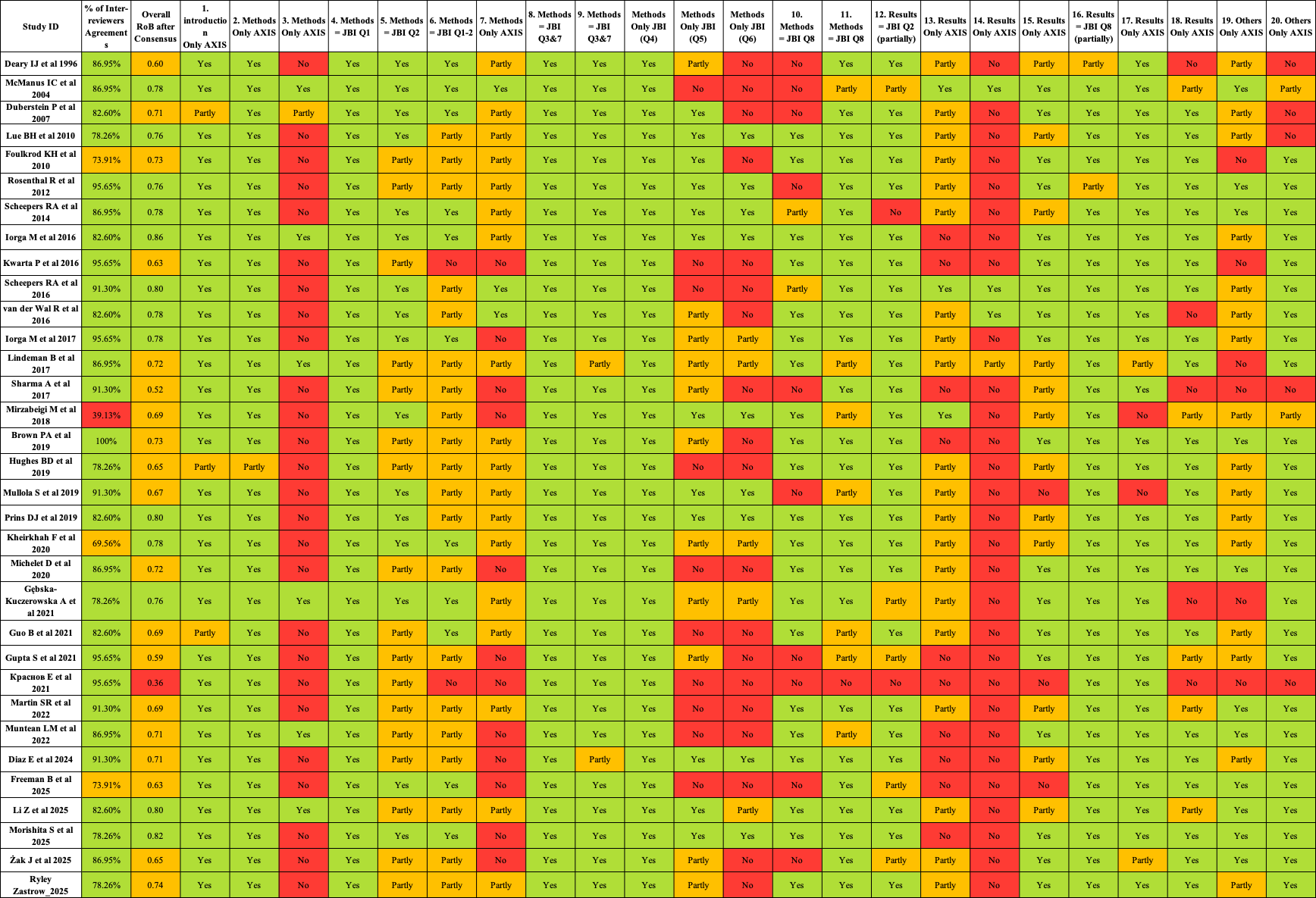

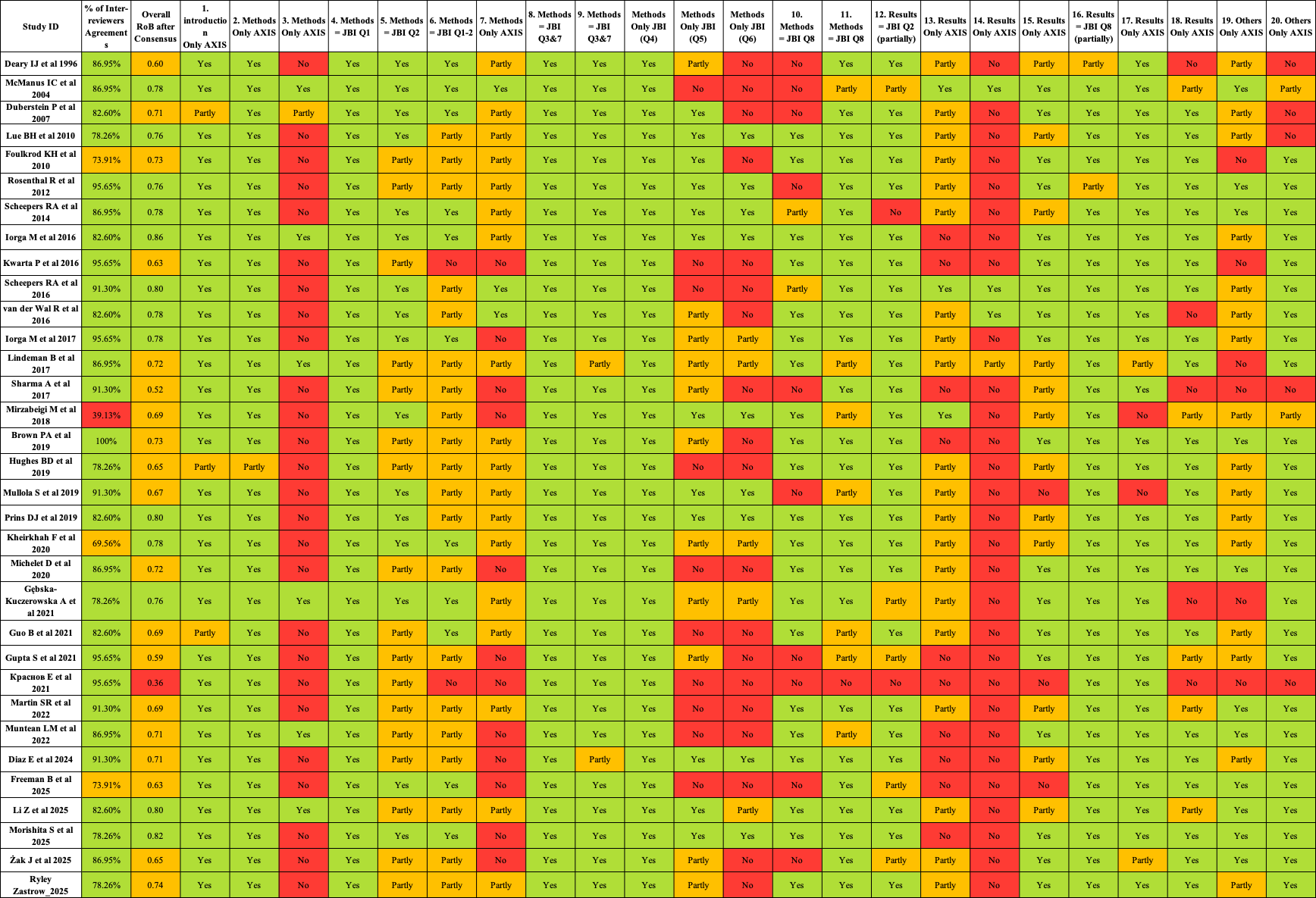

**Supplemental Methods 5. Revised Cochrane risk-of-bias tool for randomized trials (RoB 2)**

**Study details**

| **Reference** | **Schlatter S, Louisy S, Canada B, et al.**  *Personality traits affect anticipatory stress vulnerability and coping effectiveness in occupational critical care situations.*  Scientific Reports. 2022;12:20965. doi:10.1038/s41598-022-24905-z |
| --- | --- |

**Study design**

| X | Individually-randomized parallel-group trial |
| --- | --- |
| ⬜ | Cluster-randomized parallel-group trial |
| ⬜ | Individually randomized cross-over (or other matched) trial |

**For the purposes of this assessment, the interventions being compared are defined as**

| Experimental: | 2 groups (Relaxation breathing and cardiac biofeedback for 5 min) | Comparator: | Usual professional activity (reviewing printed laboratory test results for 5 min) |
| --- | --- | --- | --- |

| **Specify which outcome is being assessed for risk of bias** | Cardiac coherence score during the 5-minute coping intervention |
| --- | --- |

| **Specify the numerical result being assessed.** In case of multiple alternative analyses being presented, specify the numeric result (e.g. RR = 1.52 (95% CI 0.83 to 2.77) and/or a reference (e.g. to a table, figure or paragraph) that uniquely defines the result being assessed. | Linear regression analysis was used to predict psychological anticipatory stress (STAI-Y), cardiac reactivity, and cardiac coherence by personality trait. For all regression models, the β (i.e., estimate the effect on the outcome of each 1-unit increase in the independent variable) and the adjusted coefficients R2 (i.e., percentage of variance explained) were provided. |
| --- | --- |

**Is the review team’s aim for this result…?**

| X | to assess the effect of *assignment to intervention* (the ‘intention-to-treat’ effect) |
| --- | --- |
| ⬜ | to assess the effect of *adhering to intervention* (the ‘per-protocol’ effect) |

**If the aim is to assess the effect of *adhering to intervention***, select the deviations from intended intervention that should be addressed (at least one must be checked): N/A

⬜	occurrence of non-protocol interventions

⬜	failures in implementing the intervention that could have affected the outcome

⬜	non-adherence to their assigned intervention by trial participants

**Which of the following sources were obtained to help inform the risk-of-bias assessment? (tick as many as apply)**

X	Journal article(s) with results of the trial

⬜	Trial protocol

⬜	Statistical analysis plan (SAP)

⬜	Non-commercial trial registry record (e.g. ClinicalTrials.gov record)

⬜	Company-owned trial registry record (e.g. GSK Clinical Study Register record)

⬜	“Grey literature” (e.g. unpublished thesis)

⬜	Conference abstract(s) about the trial

⬜	Regulatory document (e.g. Clinical Study Report, Drug Approval Package)

⬜	Research ethics application

⬜	Grant database summary (e.g. NIH RePORTER or Research Councils UK Gateway to Research)

⬜	Personal communication with trialist

⬜	Personal communication with the sponsor

Risk of bias assessment

Responses underlined in green are potential markers for low risk of bias, and responses in red are potential markers for a risk of bias. Where questions relate only to sign posts to other questions, no formatting is used.

**Domain 1: Risk of bias arising from the randomization process**

**Bias arising from the randomization process**

- **Evidence**: The study states that 'Participants were then randomly assigned to three groups' [1] and 'Following the briefing, participants were randomly assigned to one of the three 5-min interventions' [4]. The flowchart shows a 'Randomized (n=147)' box leading to the three intervention groups [10]. However, details regarding the method of sequence generation (e.g., computer-generated list, random numbers) and allocation concealment (e.g., opaque envelopes, central randomization) are not provided. Baseline demographic and psychometric parameters are presented in Table 2, showing mean ± SD or % for age, weight, size, sports per week, and scores for the five personality traits [11]. No significant baseline imbalances are reported or discussed.
- **Risk of Bias Judgment**: Some concerns
- **Justification**: The authors report that participants were “randomly assigned” to three intervention groups and present a flow chart and baseline characteristics table, but they do not describe how the random sequence was generated or how allocation was concealed. No major baseline imbalances are reported.
  **Judgment:** *Some concerns* – randomization is plausible, but insufficient detail on sequence generation and concealment prevents a low-risk rating.

| **Signalling questions** | **Comments** | **Response options** |
| --- | --- | --- |
| **1.1 Was the allocation sequence random?** | Following the briefing, participants were randomly assigned to one of the three 5-min interventions. But details regarding the method of sequence generation (e.g., computer-generated list, random numbers) and allocation concealment (e.g., opaque envelopes, central randomization) are not provided. | Y / PY / PN / N / NI |
| **1.2 Was the allocation sequence concealed until participants were enrolled and assigned to interventions?** | Interventions took place in the same isolated and silent room and were all guided by the same experimenter (S.S.), but the paper gives no information on how allocation was concealed from recruiters or participants before assignment. | Y / PY / PN / N / NI |
| **1.3 Did baseline differences between intervention groups suggest a problem with the randomization process?** | Baseline characteristics are reported only for the total sample, not broken down by randomized group. | Y / PY / PN / N / NI |
| **Risk-of-bias judgement** | While randomization is stated, the lack of detail on the random sequence generation method and allocation concealment makes it impossible to determine if the process was truly unpredictable and if participants or personnel could foresee assignments. The baseline characteristics are presented, and no major imbalances are highlighted, which is positive. However, the absence of explicit methods for randomization and concealment introduces some uncertainty. | Low / High / Some concerns |
| Optional: What is the predicted direction of bias arising from the randomization process? | No clear reason to favour either the experimental or the control group. | NA / Favours experimental / Favours comparator / Towards null /Away from null / Unpredictable |

**Domain 2: Risk of bias due to deviations from the intended interventions (*effect of assignment to intervention*)**

**Bias due to deviations from intended interventions**

- **Evidence**: The interventions involved specific tasks (relaxation breathing, biofeedback, or reviewing lab results) performed by participants [4]. The study mentions that 'All the interventions were conducted in an isolated and silent room, and were all guided by the same experimenter (S.S)' [4]. This indicates that participants and the experimenter guiding the interventions were aware of the assigned intervention, meaning there was no blinding of participants or personnel. The analysis approach is not explicitly stated as intention-to-treat or per-protocol, but the results discuss the effectiveness of interventions based on the groups they were assigned to, implying an intention-to-treat approach for effectiveness analysis, although missing data was handled by exclusion for some analyses [7] [8].
- **Risk of Bias Judgment**: Some concerns

**Justification**: All interventions were delivered in a standardized way by the same experimenter in an isolated room. Participants and the experimenter were aware of group assignment, but there is no indication of systematic deviations from the assigned interventions or differential co-interventions, and analyses were conducted according to randomized groups.

| **Signalling questions** | **Comments** | **Response options** |
| --- | --- | --- |
| **2.1. Were participants aware of their assigned intervention during the trial?** | Participants clearly knew what they were doing. | Y / PY / PN / N / NI |
| **2.2. Were carers and people delivering the interventions aware of participants' assigned intervention during the trial?** | They were necessarily aware of the group assignment. | Y / PY / PN / N / NI |
| **2.3. If Y/PY/NI to 2.1 or 2.2: Were there deviations from the intended intervention that arose because of the trial context?** | No deviations are reported. | NA / Y / PY / PN / N / NI |
| **2.4 If Y/PY to 2.3: Were these deviations likely to have affected the outcome?** |  | NA / Y / PY / PN / N / NI |
| **2.5. If Y/PY/NI to 2.4: Were these deviations from intended intervention balanced between groups?** |  | NA / Y / PY / PN / N / NI |
| **2.6 Was an appropriate analysis used to estimate the effect of assignment to intervention?** |  | Y / PY / PN / N / NI |
| **2.7 If N/PN/NI to 2.6: Was there potential for a substantial impact (on the result) of the failure to analyse participants in the group to which they were randomized?** |  | NA / Y / PY / PN / N / NI |
| **Risk-of-bias judgement** | Participants and the experimenter were not blinded, but interventions were delivered per protocol in identical conditions. No important deviations due to the trial context are reported. | Low / High / Some concerns |
| Optional: What is the predicted direction of bias due to deviations from intended interventions? | No clear reason to favour either the experimental or the control group. | NA / Favours experimental / Favours comparator / Towards null /Away from null / Unpredictable |

Domain 2: Risk of bias due to deviations from the intended interventions (*effect of adhering to intervention*)

| **Signalling questions** | **Comments** | **Response options** |
| --- | --- | --- |
| **2.1. Were participants aware of their assigned intervention during the trial?** | Participants clearly knew what they were doing. | Y / PY / PN / N / NI |
| **2.2. Were carers and people delivering the interventions aware of participants' assigned intervention during the trial?** | They were necessarily aware of the group assignment. | Y / PY / PN / N / NI |
| **2.3. [If applicable:] If Y/PY/NI to 2.1 or 2.2: Were important non-protocol interventions balanced across intervention groups?** | No important non-protocol interventions are reported. | NA / Y / PY / PN / N / NI |
| **2.4. [If applicable:] Were there failures in implementing the intervention that could have affected the outcome?** |  | NA / Y / PY / PN / N / NI |
| **2.5. [If applicable:] Was there non-adherence to the assigned intervention regimen that could have affected participants’ outcomes?** |  | NA / Y / PY / PN / N / NI |
| **2.6. If N/PN/NI to 2.3, or Y/PY/NI to 2.4 or 2.5: Was an appropriate analysis used to estimate the effect of adhering to the intervention?** | All groups were guided & did only their assigned 5-min task in the same room. | NA / Y / PY / PN / N / NI |
| **Risk-of-bias judgement** |  | Low / High / Some concerns |
| Optional: What is the predicted direction of bias due to deviations from intended interventions? |  | NA / Favours experimental / Favours comparator / Towards null /Away from null / Unpredictable |

**Domain 3: Missing outcome data**

**Bias due to missing outcome data**

- **Evidence**: The initial sample size was 147 participants [3]. However, the analysis for personality and STAI-Y (psychological anticipatory stress) included 125 participants, with 22 excluded due to missing data [7]. For personality and cardiac reactivity (physiological anticipatory stress), 120 participants were included, with 27 excluded due to missing data [7]. For personality and cardiac coherence (coping effectiveness), 118 participants were included, with 29 excluded due to missing data [8]. The reasons for these exclusions are stated as 'missing data' but are not elaborated upon (e.g., participant withdrawal, technical issues). The methods used to handle missing data appear to be complete-case analysis (exclusion).
- **Risk of Bias Judgment**: Some concerns
- **Justification**: Of 147 randomized participants, 15–20% were excluded from analyses for different outcomes because of “missing data”, with no further breakdown of reasons. Numbers retained per arm remained broadly similar. Complete-case analyses were used.

| **Signalling questions** | **Comments** | **Response options** |
| --- | --- | --- |
| **3.1 Were data for this outcome available for all, or nearly all, participants randomized?** | 147 residents were randomized, but only about 118 were included in the cardiac-coherence analyses (≈80%). | Y / PY / PN / N / NI |
| **3.2 If N/PN/NI to 3.1: Is there evidence that the result was not biased by missing outcome data?** | A flow diagram shows exclusions, but the paper does not clearly describe reasons for all missing cardiac-coherence data or compare participants with and without outcome data by group or baseline characteristics. There is no direct evidence that missingness is harmless. | NA / Y / PY / PN / N |
| **3.3 If N/PN to 3.2: Could missingness in the outcome depend on its true value?** | With no detailed information on patterns of missingness, it is unclear whether participants with more (or less) stress-related physiological responses were more likely to have missing data & whether it is likely that missingness depended on the true cardiac-coherence values. | NA / Y / PY / PN / N / NI |
| **3.4 If Y/PY/NI to 3.3: Is it likely that missingness in the outcome depended on its true value?** | With no detailed information on patterns of missingness, it is unclear whether participants with more (or less) stress-related physiological responses were more likely to have missing data & whether it is likely that missingness depended on the true cardiac-coherence values. | NA / Y / PY / PN / N / NI |
| **Risk-of-bias judgement** | Around 20% of randomized participants lack cardiac-coherence data, and the reasons and patterns of missingness are insufficiently described. | Low / High / Some concerns |
| Optional: What is the predicted direction of bias due to missing outcome data? | It’s not possible to predict whether missing outcome data would tend to favour the experimental or control group or move results towards/away from the null. | NA / Favours experimental / Favours comparator / Towards null /Away from null / Unpredictable |

**Domain 4: Risk of bias in measurement of the outcome**

**Bias in measurement of the outcome**

- **Evidence**: Psychological anticipatory stress was measured by the STAI-Y questionnaire [5], a validated tool [12]. Physiological anticipatory stress was measured by cardiac reactivity, calculated from mean heart rate [5]. Cardiac coherence, an indicator of coping effectiveness, was measured using an ear pulse sensor and computed as (Peak Power / [Total Power - Peak Power]) on a min-by-min basis [6]. The STAI-Y is a self-report measure, and heart rate measurements are objective. Outcome assessors for the STAI-Y (self-report) were not blinded, and for cardiac reactivity and coherence, the measurements are objective and automated (ear pulse sensor, EmWave software), reducing assessor bias [6]. However, the experimenter (S.S.) who guided the interventions was not blinded [4], which could potentially influence how data was collected or handled, even for objective measures.
- **Risk of Bias Judgment**: Some concerns

**Justification**: Anticipatory stress was measured using the validated STAI-Y questionnaire, and physiological outcomes (cardiac reactivity and cardiac coherence) were derived from automated heart-rate recordings via ear-pulse sensors and dedicated software. The experimenter and participants were unblinded, which could influence self-reported stress, but objective HRV measures are unlikely to be affected.

| **Signalling questions** | **Comments** | **Response options** |
| --- | --- | --- |
| **4.1 Was the method of measuring the outcome inappropriate?** |  | Y / PY / PN / N / NI |
| **4.2 Could measurement or ascertainment of the outcome have differed between intervention groups?** |  | Y / PY / PN / N / NI |
| **4.3 If N/PN/NI to 4.1 and 4.2: Were outcome assessors aware of the intervention received by study participants?** | They were necessarily aware of the group assignment. | NA / Y / PY / PN / N / NI |
| **4.4 If Y/PY/NI to 4.3: Could assessment of the outcome have been influenced by knowledge of intervention received?** | Outcome values are generated automatically by the software; the experimenter cannot easily manipulate cardiac-coherence scores during recording. | NA / Y / PY / PN / N / NI |
| **4.5 If Y/PY/NI to 4.4: Is it likely that assessment of the outcome was influenced by knowledge of intervention received?** |  | NA / Y / PY / PN / N / NI |
| **Risk-of-bias judgement** |  | Low / High / Some concerns |
| Optional: What is the predicted direction of bias in measurement of the outcome? |  | NA / Favours experimental / Favours comparator / Towards null /Away from null / Unpredictable |

**Domain 5: Risk of bias in selection of the reported result**

**Bias in selection of the reported result**

- **Evidence**: The study clearly states its aim: 'to investigate the influence of personality on both anticipatory stress vulnerability and the effectiveness of coping strategies' [1]. The introduction outlines hypotheses related to neuroticism, extraversion, and conscientiousness affecting anticipatory stress vulnerability, and openness affecting coping effectiveness [2]. The results section directly addresses these hypotheses, presenting findings for personality and anticipatory stress vulnerability, and personality and coping intervention effectiveness [13]. The paper does not mention a pre-registered protocol or trial registration number. There is no explicit evidence of selective reporting of outcomes or analyses, as the reported results align with the stated objectives and hypotheses. The main outcomes (STAI-Y, cardiac reactivity, cardiac coherence) are consistently reported throughout the methods and results sections.
- **Risk of Bias Judgment**: Low risk
- **Justification**: The prespecified aims were to examine the influence of personality on anticipatory stress vulnerability and coping effectiveness. All main outcomes (STAI-Y, cardiac reactivity and cardiac coherence) and the corresponding personality analyses are reported, and there is no evidence of unreported primary outcomes. The study was not formally registered.

| **Signalling questions** | **Comments** | **Response options** |
| --- | --- | --- |
| **5.1 Were the data that produced this result analysed in accordance with a pre-specified analysis plan that was finalized before unblinded outcome data were available for analysis?** | Statistical analysis methods are described in the paper, and ethics approval is reported, but there is no protocol, registry entry, or statement that the analysis plan was finalized before unblinded outcome data were available. | Y / PY / PN / N / NI |
| **Is the numerical result being assessed likely to have been selected, on the basis of the results, from...** |  |  |
| **5.2. ... multiple eligible outcome measurements (e.g. scales, definitions, time points) within the outcome domain?** | The outcome is a single, clearly defined HRV metric (“cardiac coherence”) with its computation taken from prior literature. Alternative analysis sets are not mentioned | Y / PY / PN / N / NI |
| **5.3 ... multiple eligible analyses of the data?** | The paper presents one main set of regression models for cardiac coherence. Alternative analysis sets are not mentioned | Y / PY / PN / N / NI |
| **Risk-of-bias judgement** |  | Low / High / Some concerns |
| Optional: What is the predicted direction of bias due to selection of the reported result? |  | NA / Favours experimental / Favours comparator / Towards null /Away from null / Unpredictable |

**Overall risk of bias**

**Overall Risk of Bias Assessment**

Considering the assessments across all domains, the overall risk of bias for this study is Some concerns.

The primary drivers for this judgment are:

- **Some concerns due to the** **lack of blinding for both participants and the experimenter**.
- **Some concerns due to randomization process**: While randomization was stated, the insufficient detail on sequence generation and allocation concealment means that selection bias cannot be definitively ruled out.
- **Some concerns due to missing outcome data**: The moderate attrition rates with unspecified reasons and the use of complete-case analysis without further justification or sensitivity analysis introduce uncertainty.

| **Risk-of-bias judgement** | Domain 1: Some concerns  Domain 2: Low risk  Domain 3: Some concerns  Domain 4: Low risk  Domain 5: Some concerns | Low / High / Some concerns |
| --- | --- | --- |
| Optional: What is the overall predicted direction of bias for this outcome? | It’s not possible to confidently say any bias would push results towards or away from the null or favour either group. | NA / Favours experimental / Favours comparator / Towards null /Away from null / Unpredictable |

**Recommendations**

To improve the robustness and reduce the risk of bias in future studies of this nature, the following recommendations are made:

- **Implement robust blinding**: For interventions where blinding of participants is difficult (e.g., active vs. control), blinding of outcome assessors and personnel involved in data collection should be prioritized. If possible, use a sham or placebo control to blind participants to their intervention status.
- **Detail randomization and allocation**: Clearly describe the methods used for random sequence generation (e.g., computer-generated random numbers) and allocation concealment (e.g., sequentially numbered, opaque, sealed envelopes) to ensure transparency and minimize selection bias.
- **Address missing data comprehensively**: Report detailed reasons for all missing data. If attrition occurs, employ appropriate statistical methods for handling missing data (e.g., multiple imputation) and conduct sensitivity analyses to assess the impact of missing data on the results.
- **Pre-register protocols**: Registering the study protocol in a public registry (e.g., ClinicalTrials.gov) before participant enrollment would enhance transparency by documenting the study design, outcomes, and analysis plan, thereby mitigating concerns about selective reporting.

**Supplemental Methods 6. Descriptive and Quantitative Synthesis of the Outcomes**

1. **Stress-related outcomes**

Consistent with our prespecified primary outcome, stress-related measures were operationalized heterogeneously across the included studies and covered several related constructs, including burnout, broader psychological distress, perceived work stress, acute stress responses, and severe stress-related outcomes. Sixteen studies assessed burnout using Maslach Burnout Inventory (MBI) configurations or related instruments, including the Maslach Burnout Inventory-Human Services Survey (MBI-HSS), Dutch adaptations of the MBI (UBOS-C), the Polish Burnout Scale (BOS), and the Copenhagen Burnout Inventory. ^1–16^ Of these, eleven studies (55 effect sizes) reported data for all three subscales and were therefore eligible for quantitative synthesis. ^1–8,11–13^ The remaining five burnout studies were retained for narrative synthesis because they reported only total burnout scores or used non-comparable burnout measures.

Broader psychological distress-related outcomes were also assessed with a range of instruments. Four studies used the General Health Questionnaire (GHQ) in either the 12-item or 28-item format;^2,3,7,17^ among these, the three GHQ-12 studies were sufficiently comparable to be included in the quantitative synthesis (10 effect sizes).^2,7,17^ Other studies assessed related mental health constructs using self-reported depression^4^ or emotional affect measured with the Positive and Negative Affect Schedule (PANAS).^10^ Five studies specifically addressed perceived work stress or acute stress responses, using instruments such as the Specialist Doctors Stress Inventory,^2^ the Shortened Stress Evaluation Tool (ASSET),^18^ the Job Stress Questionnaire,^10^ a combination of state anxiety measures (STAI-Y) and physiological indicators of cardiac reactivity,^19^ or Perceived Stress Scale (PSS).^16^ Two studies investigated more severe outcomes related to work, specifically suicidal ideation or the wish to die.^17,20^

In the meta-analysis of the three burnout subscales, Neuroticism showed the clearest and most consistent adverse pattern across burnout dimensions and stress. Forest plot inspection revealed considerable variation in effect sizes across studies, with confidence intervals ranging from near-zero to moderate positive correlations, though the overall direction was consistently positive except for personal accomplishment. Higher Neuroticism was associated with greater emotional exhaustion (pooled r =0.42, 95% CI 0.22–0.62, 95% PI −0.23 to 1.06, p<0.0001) and depersonalization (pooled r =0.30, 95% CI 0.17–0.44, 95% PI −0.12 to 0.73, p<0.0001), and with lower personal accomplishment (pooled r =−0.24, 95% CI −0.39 to −0.09, 95% PI −0.71 to 0.22, p=0.005). The separate meta-analysis of psychological distress likewise showed a positive association with Neuroticism (pooled r =0.52, 95% CI 0.18–0.87, 95% PI −0.15 to 1.2, p=0.022).

Compared with Neuroticism, Conscientiousness showed a small protective pattern, with inverse associations for emotional exhaustion (pooled r =−0.19, 95% CI −0.26 to −0.12, 95% PI −0.35 to −0.03, p<0.001) and depersonalization (pooled r =−0.24, 95% CI −0.34 to −0. 14, 95% PI −0.51 to 0.04, p<0.001), and a positive association with personal accomplishment (pooled r=0.23, 95% CI 0.05 to 0.41, 95% PI −0.35 to 0.82, p=0.018). Its pooled association with stress was also small but not statistically significant (r=−0.12, 95% CI −0.49 to 0.26, 95% PI −0.86 to 0.62, p=0.307).

Extraversion also appeared protective for burnout dimensions; emotional exhaustion (pooled r=−0.26, 95% CI −0.34 to −0.15, 95% PI −0.57 to 0.06, p<0.001) and depersonalization (pooled r=−0.21, 95% CI −0.32 to −0.09, 95% PI −0.55 to 0.14, p=0.003) and was positively associated with personal accomplishment (pooled r=0.204, 95% CI 0.09 to 0.32, 95% PI −0.11 to 0.52, p=0.002). However, as with Conscientiousness, the pooled association with stress was not significant (r=−0.16, 95% CI −0.79 to 0.47, 95% PI −1.42 to 1.09, p=0.386).

Agreeableness was similarly associated with lower emotional exhaustion (pooled r=−0.18, 95% CI −0.28 to −0.09, 95% PI −0.44 to 0.07, p=0.002) and lower depersonalization (pooled r=−0.36, 95% CI −0.45 to −0.28, 95% PI −0.58 to −1.44, p<0.001), and with higher personal accomplishment (pooled r=0.25, 95% CI 0.12 to 0.38, 95% PI −0.14 to 0.64, p=0.002), although its association with psychological stress was not significant (pooled r=−0.09, 95% CI −0.55 to 0.36, 95% PI −1 to 0.81, p=0.472).

Openness to Experience showed a selective rather than uniform pattern of association. It was not significantly associated with emotional exhaustion (pooled r=−0.03, 95% CI −0.09 to 0.03, 95% PI −0.16 to 0.11, p=0.331) or psychological stress (pooled r=0.007, 95% CI −0.15 to 0.16, 95% PI −0.28 to 0.29, p=0.854), indicating essentially null relationships for these outcomes. By contrast, Openness was associated with lower depersonalization (pooled r=−0.134, 95% CI −0.25 to −0.02, 95% PI −0.46 to 0.2, p=0.026) and higher personal accomplishment (pooled r=0.17, 95% CI 0.07 to 0.27, 95% PI −0.1 to 0.44, p=0.003), suggesting a small protective role for the more attitudinal dimensions of burnout.

To note, heterogeneity was substantial in all models (I²>75%), indicating marked between-effect variability across samples and settings. Funnel-plot asymmetry tests and Egger’s tests were generally non-significant for personality traits across burnout dimensions, suggesting no clear evidence of small-study effects. The GHQ-12 analysis (k=3) was too sparse for funnel-plot assessment. Full details of the meta-analyses are presented in Supplementary Methods 7-19.

In post-hoc moderator analyses by experience level (n = 8), region (n = 11), and specialty (n = 6), some studies were excluded because their reporting lacked sufficient details. Full details of exploratory moderator analyses are presented in Supplementary Methods 7-19.

Across moderator analyses, the Neuroticism–burnout association varies more clearly by experience and region than by specialty, and that its strongest and most consistent links are with emotional exhaustion (pooled effects r = 0.33 to 0.51), followed by depersonalization (pooled effects r = 0.27 to 0.32), whereas its association with personal accomplishment was weaker and inverse (pooled effects r =−0.33 to −0.17).

Conscientiousness showed a broadly protective pattern across burnout dimensions, with the clearest inverse associations for depersonalization (pooled effects r = −0.18 to −0.29) and emotional exhaustion (pooled effects r = −0.16 to −0.26), but an inconsistent variable positive association with personal accomplishment (pooled effects r = 0.19 to 0.24; some p-values > 0.05). Like Neuroticism, these associations varied more clearly by experience and region than by specialty.

Extraversion showed a generally protective association with burnout, most clearly for emotional exhaustion (pooled effects r = −0.22 to −0.23), followed by depersonalization (pooled effects r = −0.15 to −0.20), while its positive association with personal accomplishment was smaller and more context dependent (pooled effects r = 0.17 to 0.19; most p-values > 0.05). The clearest moderator signal came from experience, whereas region and specialty produced non-significant moderation effects.

Agreeableness showed a generally protective association with burnout, most clearly for depersonalization (pooled effects r = −0.33 to −0.36), followed by emotional exhaustion (pooled effects r = −0.11 to −0.20), and with a positive association with personal accomplishment (pooled effects r = 0.17 to 0.31; some p-values > 0.05). Compared with the other protective traits, Agreeableness showed clearest moderator signals by experience and region, while specialty-based models were non-significant.

Openness to Experience showed a weak and inconsistent pattern across burnout dimensions. Its associations with depersonalization were small and negative (pooled effects r = −0.11 to −0.14), and its associations with personal accomplishment were small and positive (pooled effects r = 0.11 to 0.20). While its associations with emotional exhaustion were non-significant. Compared with the other big five personality traits, there was little evidence that experience, region, or specialty clearly moderated these associations.

Across studies that assessed burnout with instruments not included in the main meta-analyses (including MBI total scores, the Polish Burnout Scale and the Copenhagen Burnout Inventory and variants), Neuroticism consistently predicted higher burnout across instruments, countries and specialties, typically with small-to-moderate correlations. By contrast, the other Big Five traits showed only modest and often inconsistent protective associations.

Across the other subset of studies examining psychological distress, affect, and suicidality, findings were broadly convergent despite heterogeneity in instruments and designs. Neuroticism emerged as the most consistent vulnerability marker, showing generally positive correlations with psychological distress, perceived job stress, negative affectivity and self-declared depression, and higher anticipatory stress in experimental settings. By contrast, Conscientiousness, Extraversion, and Agreeableness appeared to confer small protective effects, with complex, gender-specific moderation patterns for extreme outcomes such as wish to die or suicidal ideation. Openness remained the least consistently associated with stress-related measures.Bottom of Form

2. **Coping Outcomes**

Coping outcomes were captured using standardized questionnaires assessing coping styles and more targeted indices of behavioral or physiological coping. Two studies used the Coping Inventory for Stressful Situations (CISS),^3,21^ which distinguishes task-oriented (TOC), emotion-oriented (EOC), and avoidance-oriented coping (AOC; with distraction and social diversion as avoidance subscales), whereas one study used the Coping Strategies Inventory^10^ and two studies used Coping Strategies scale.^15,22^ Two additional studies approached coping from a more concrete angle, either via self-reported use of medication to cope with stress^4^ or via physiological parameters such as cardiac coherence during a stress-management intervention.^19^

Although there were only a few studies, and some used broad coping measures that hid the more complex links between personality and coping, the findings still pointed in the same direction. Neuroticism consistently associated with emotion-focused and disengagement strategies. Conscientiousness, and to lesser extent Extraversion, were linked to adaptive coping, with positive correlations for problem-focused approaches. Agreeableness showed a prosocial coping profile, though correlations were generally weaker. Openness displayed minimal associations with coping styles.

3. **Performance, professionalism, and other work-related outcomes**

Performance and work-related outcomes were heterogeneous and included supervisor teaching performance (System for Evaluation of Teaching Qualities, SETQ),^23,24^ technical performance on a laparoscopy simulator,^25^ job performance,^26^ work ability,^17^ customized 20-item technical performance checklist,^27^ job satisfaction,^2,15,28,29^ work engagement,^23^ specific motivational constructs (e.g. perfectionism,^5^ grit,^30^ achievement motivation,^26^ gratitude questionnaire^22^), organizational commitment^26^ and workplace climate.^2^ Non-technical aptitudes outcomes included empathy (Jefferson Scale of Empathy), trait emotional intelligence and related constructs, tolerance–intolerance of uncertainty,^31^ shared decision making (physician SDM scales^32^ and patient-rated SDM-Q-9^33^), and patient satisfaction questionnaire^34^.

Studies examining links between the Big Five personality traits and outcomes such as performance, technical skills, and professionalism were highly heterogeneous. Many studies rely on a single instrument or dataset, and most report small, inconsistent associations. As a result, these findings should be interpreted as suggestive rather than conclusive. Across the literature, the most consistent pattern is that Conscientiousness—and, to a lesser extent, Extraversion and Agreeableness—is associated with more favorable outcomes, whereas Neuroticism tends to show negative associations. Openness demonstrates the weakest and most inconsistent relationships. Studies investigating sleep problems similarly reveal variable and sometimes conflicting results.
