## Supplementary material for "Influence of Personality Traits on Mental Health, Coping, and Professional Performance in Medical Residents and Physicians: Systematic Review and Meta-Analysis": Meta-analysis Output(7-19 Supp): Appendices_Meta-analysis_16082026.docx

**Supplementary Methods 7-19. Full details of exploratory moderator analyses**

**Supplementary Methods 7a (Agreeableness vs Depersonalization)**

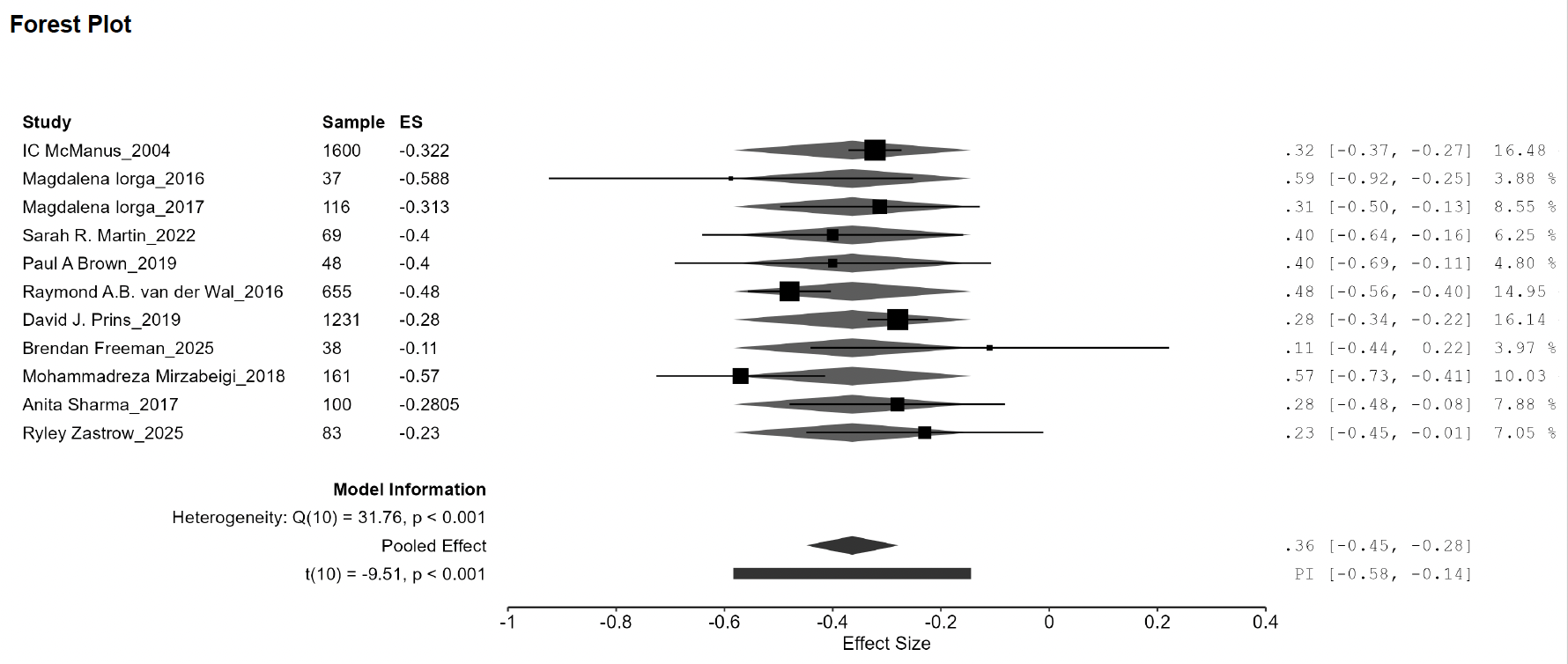

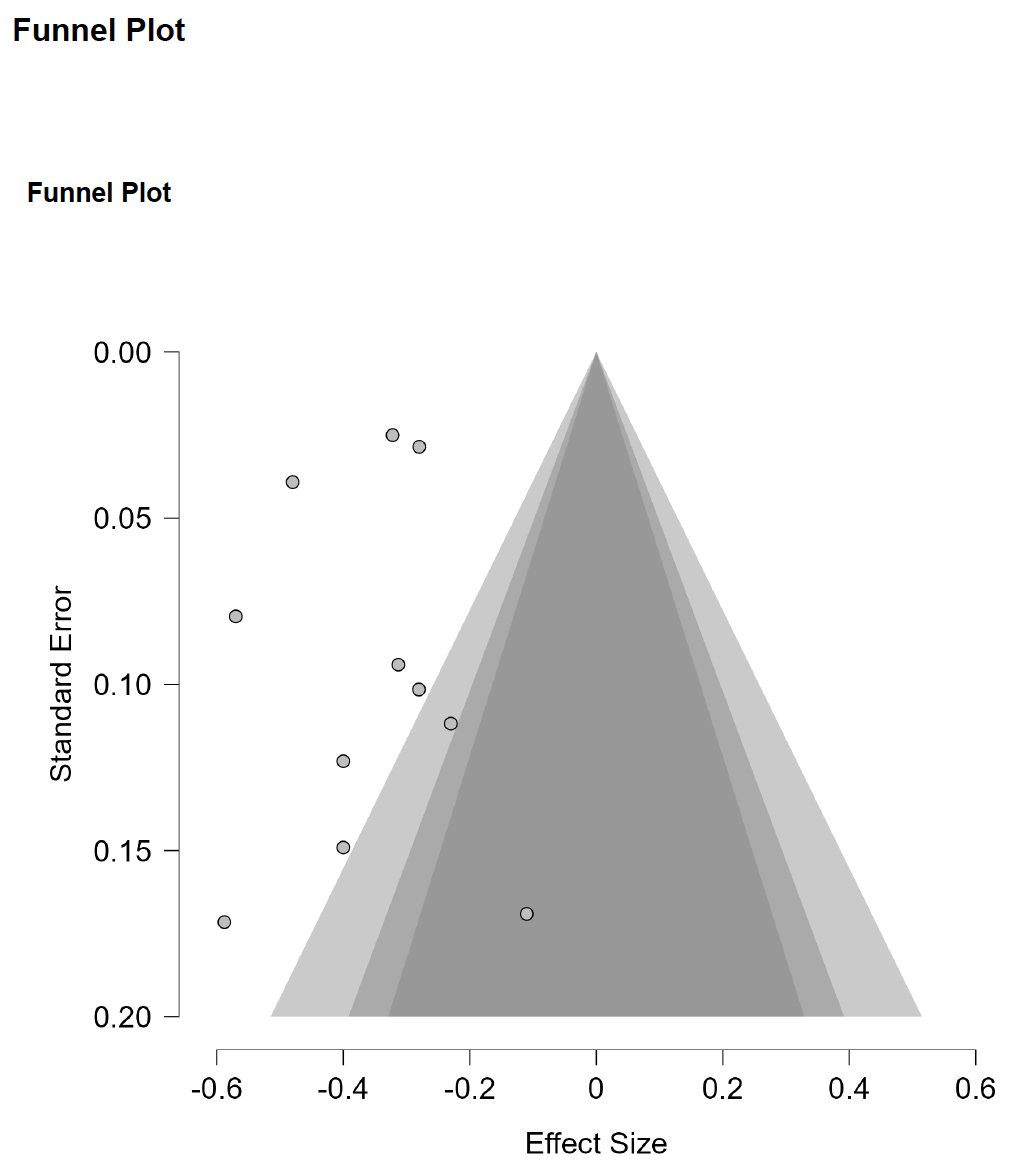

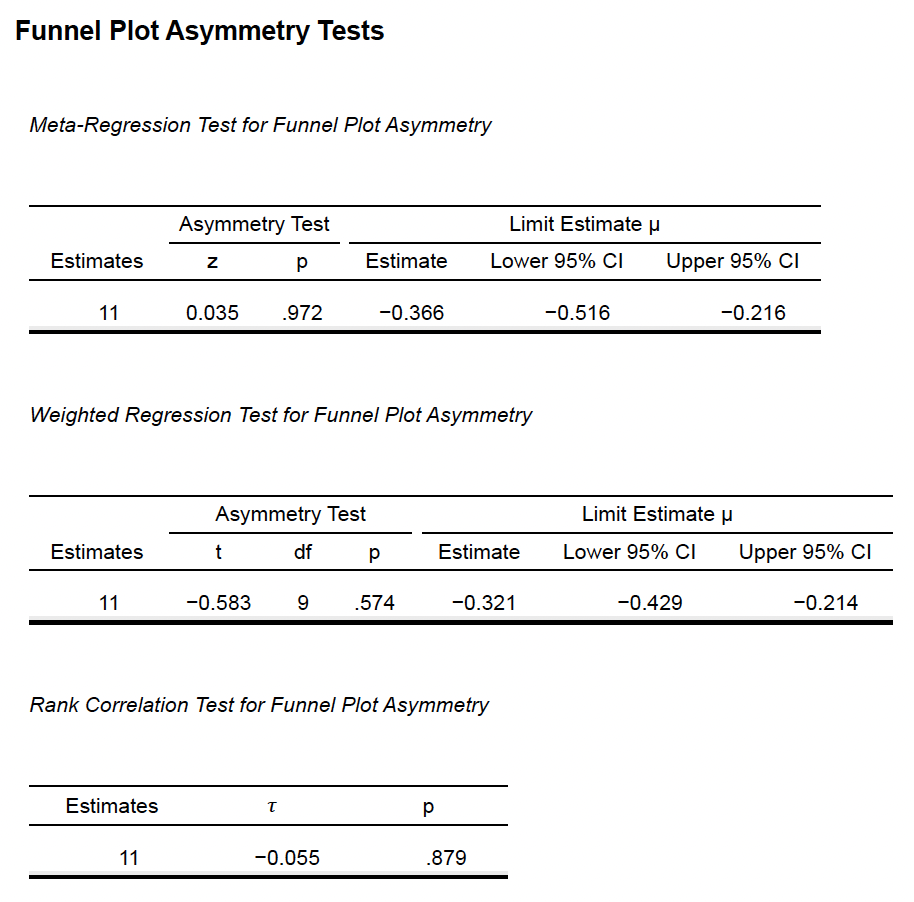

**Supplementary Methods 7b (Conscientiouness vs Depersonalization)**

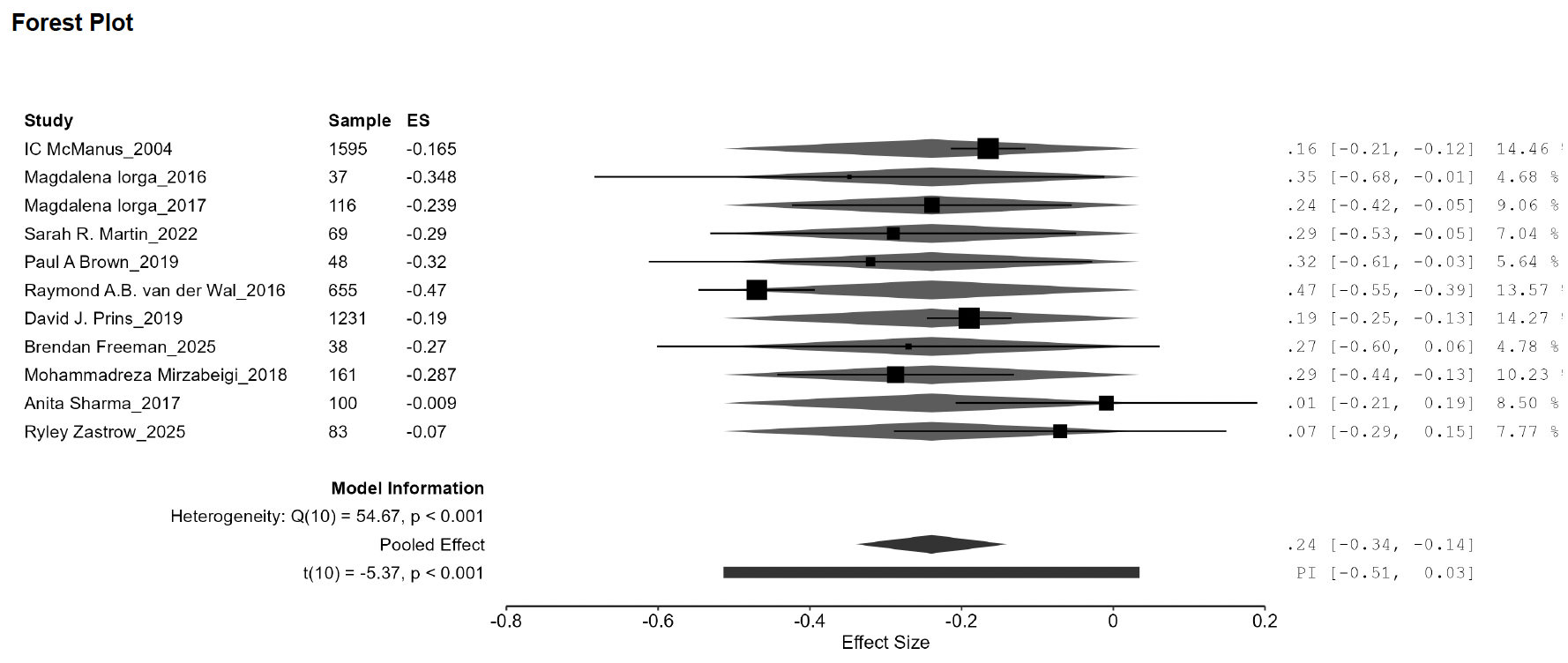

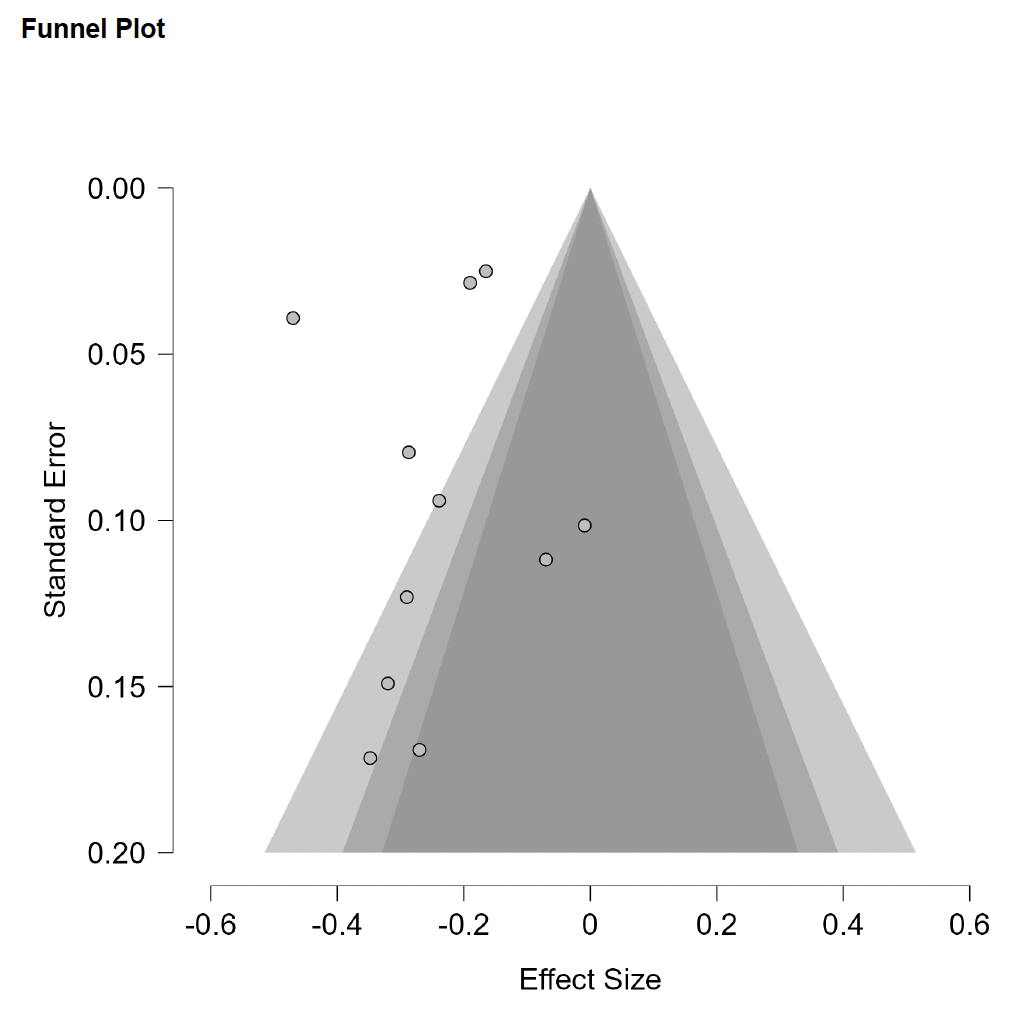

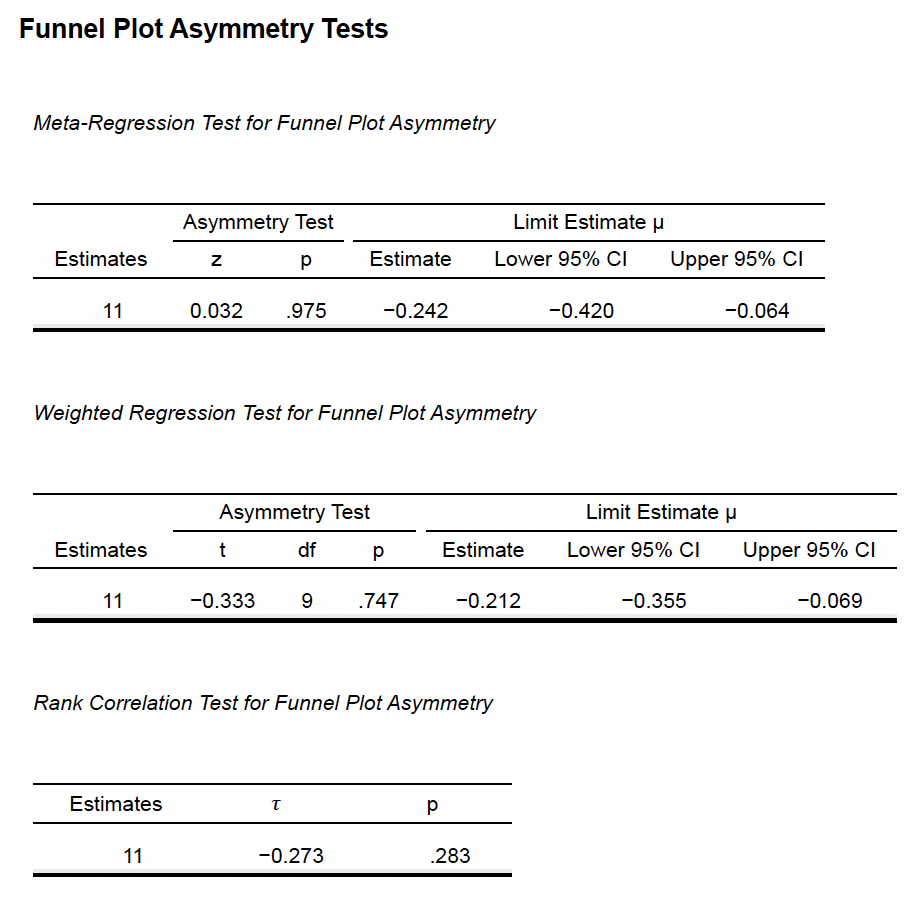

**Supplementary Methods 7c (Extraversion vs Depersonalization)**

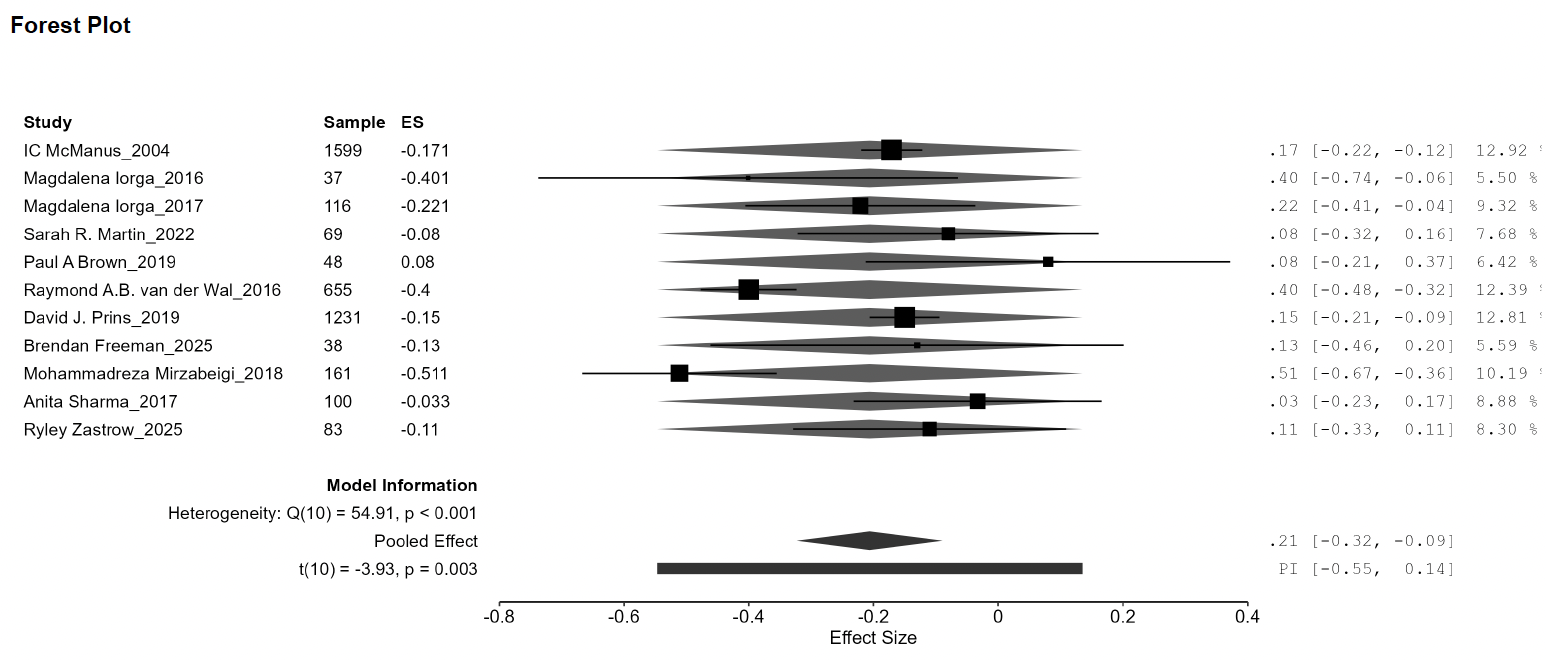

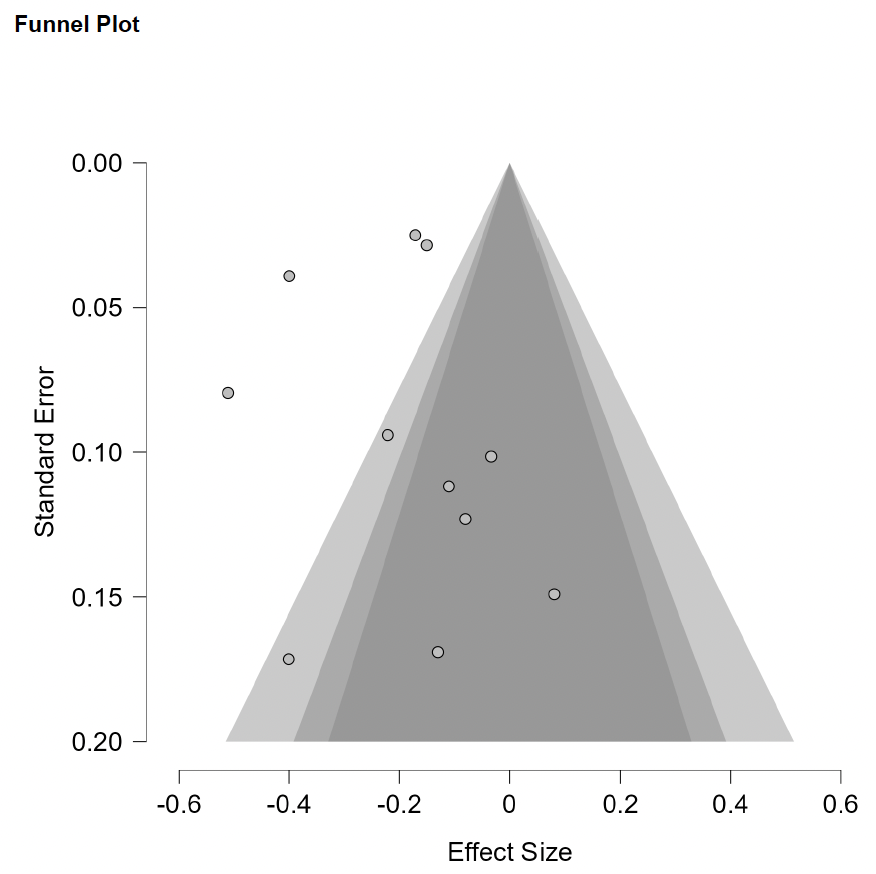

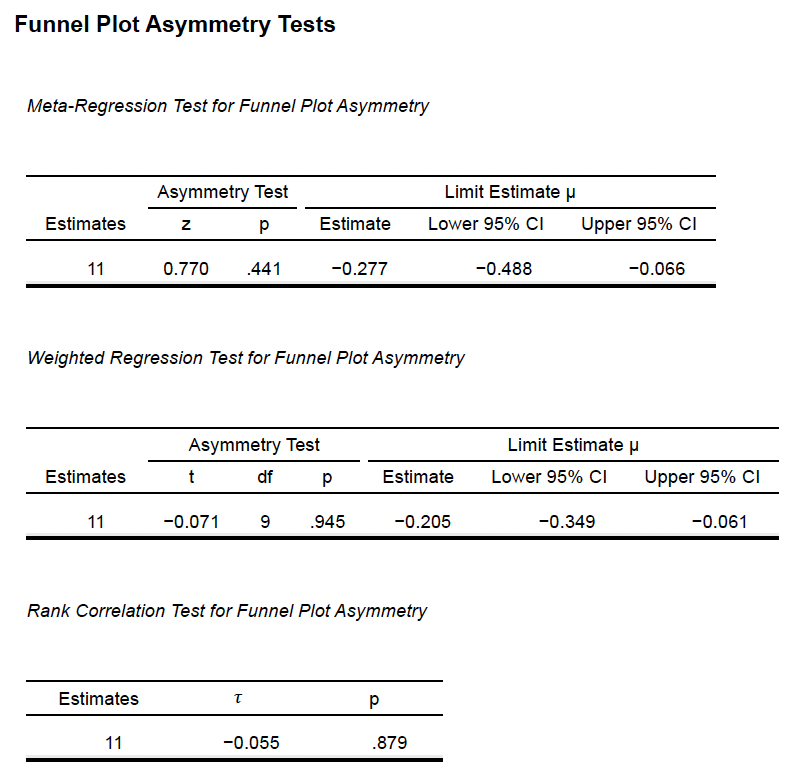

**Supplementary Methods 7d (Neuroticism vs Depersonalization)**

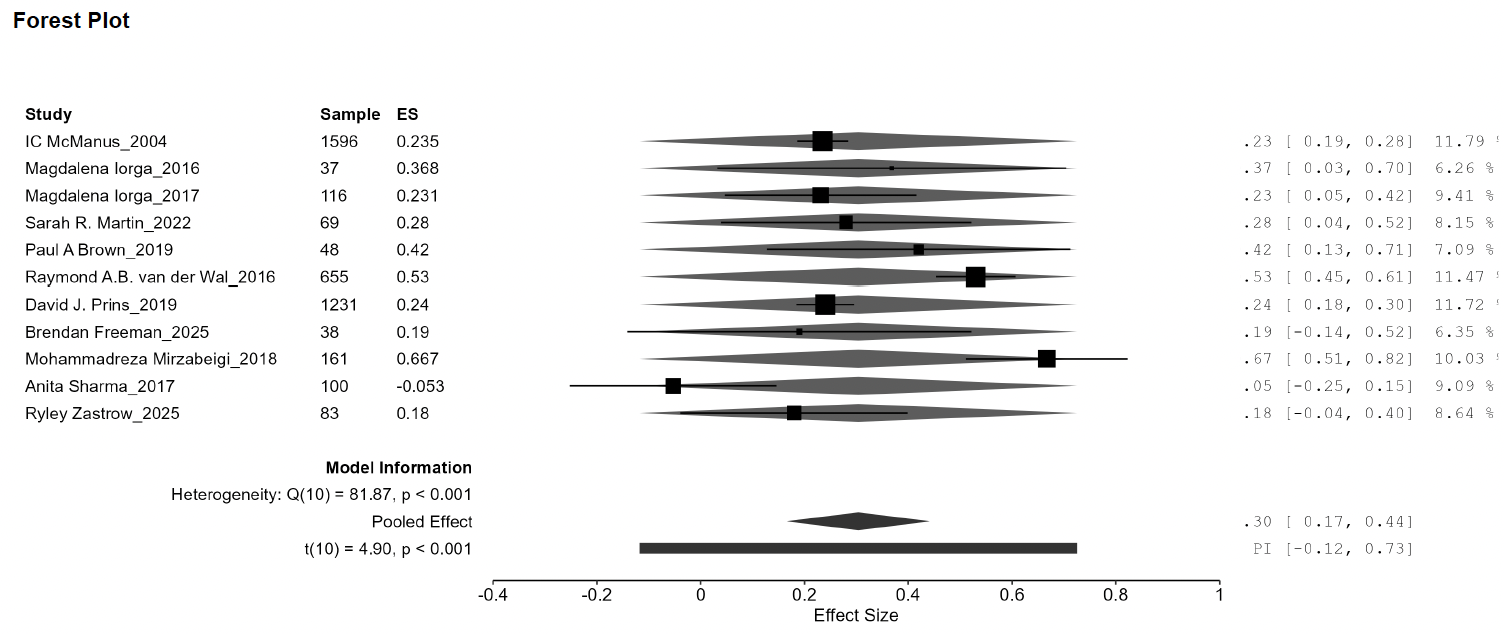

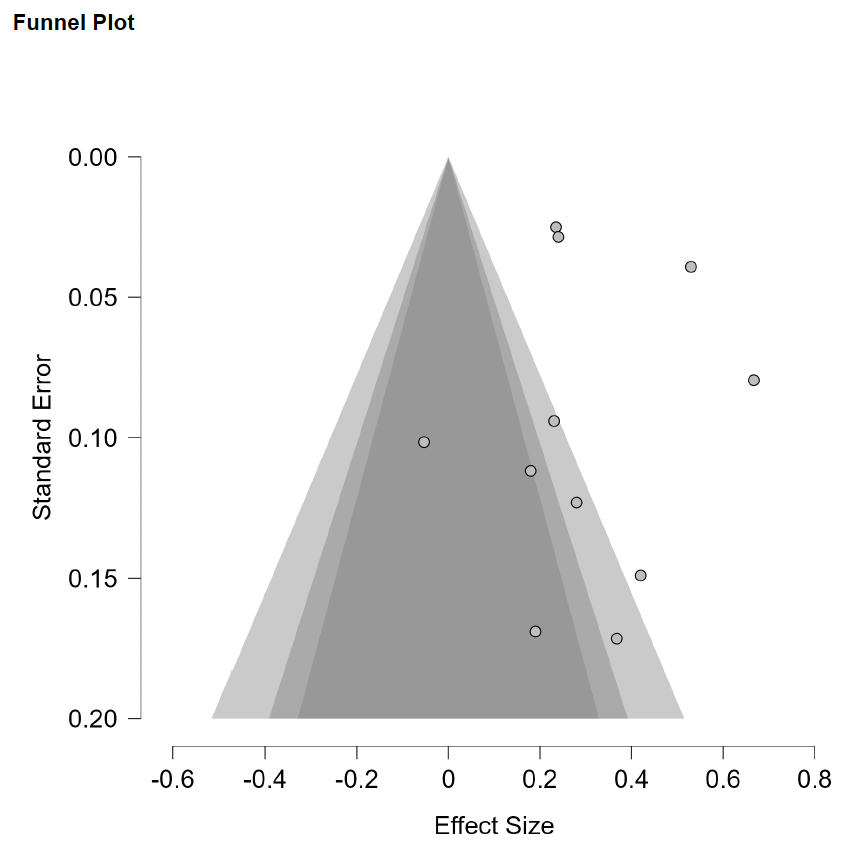

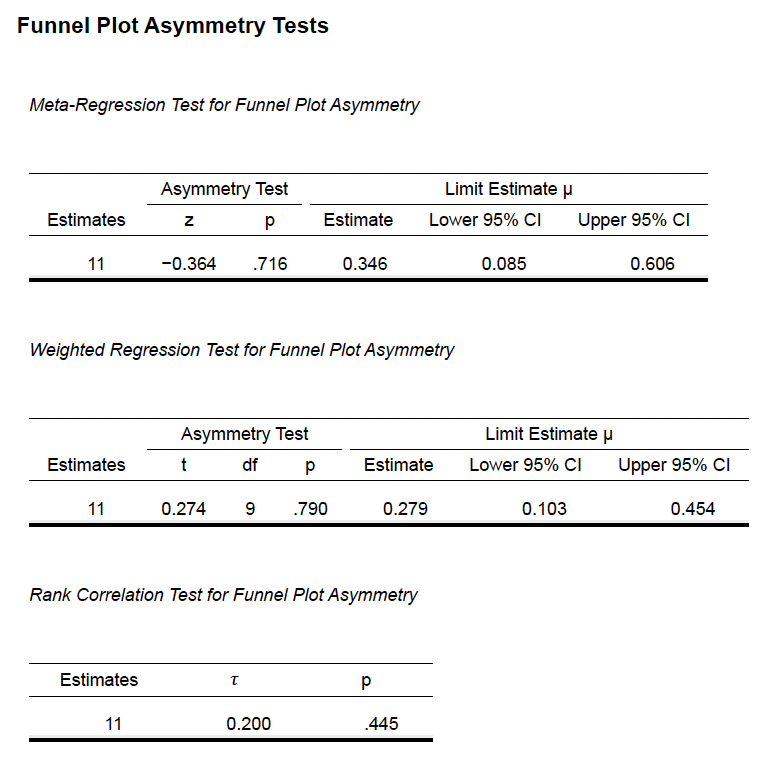

**Supplementary Methods 7e (Openness vs Depersonalization)**

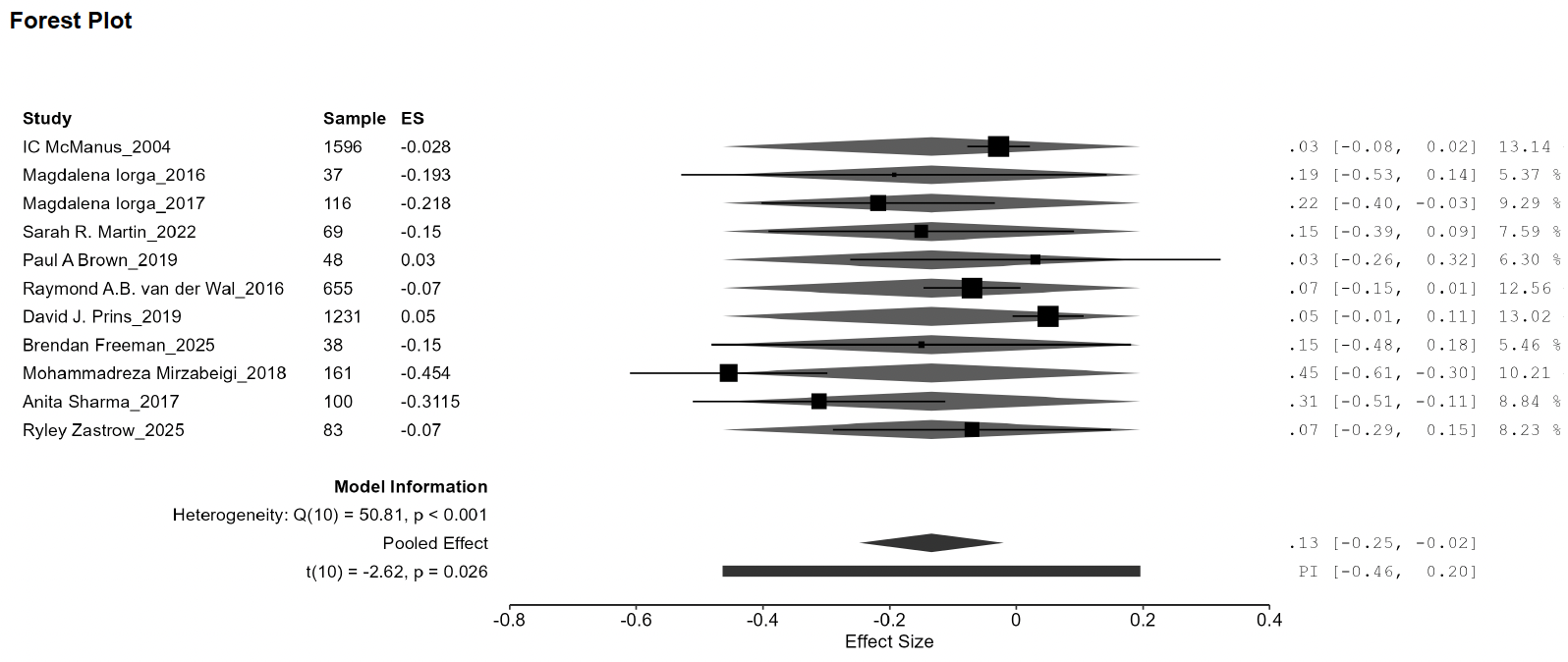

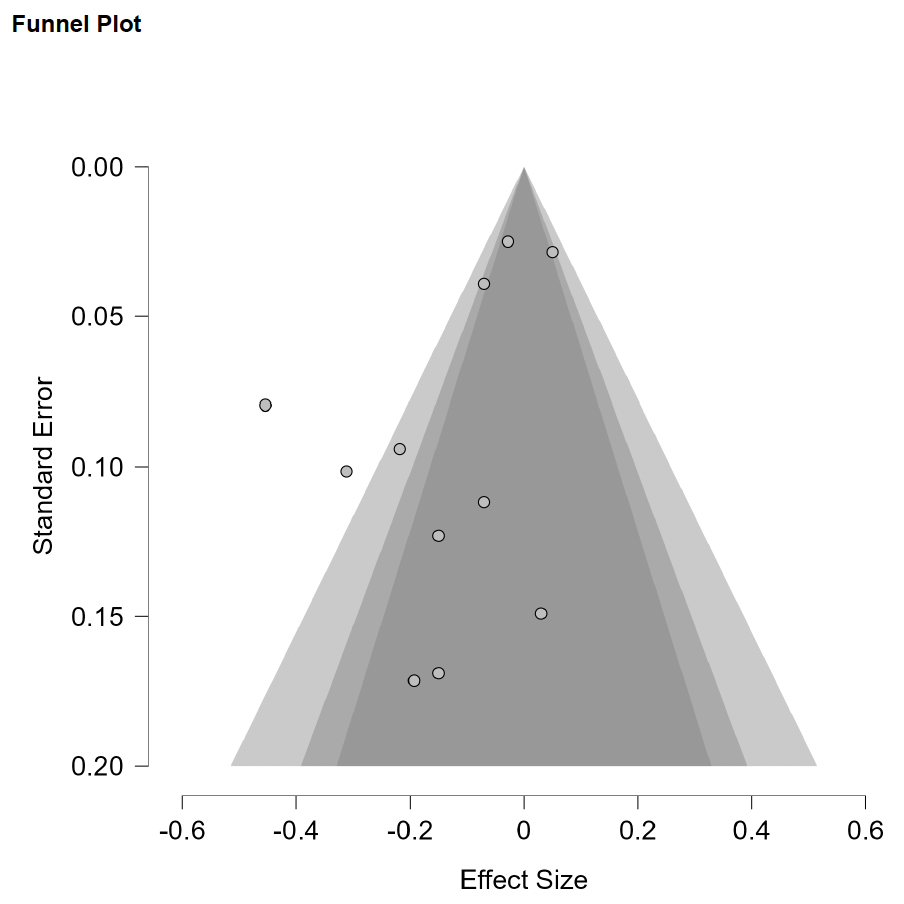

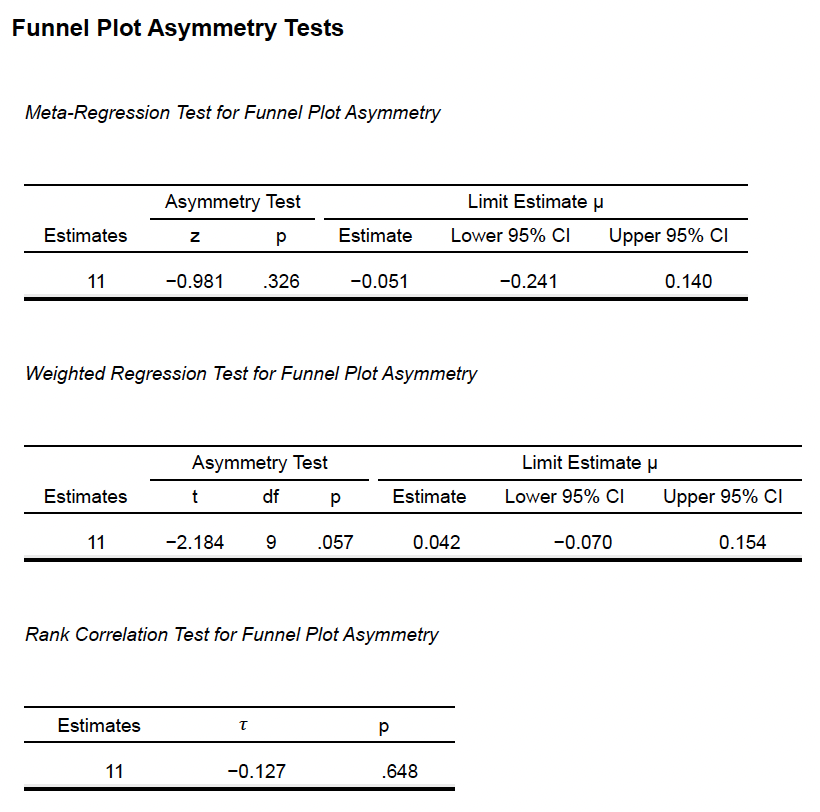

**Supplementary Methods 8a (Agreeableness vs Emotional Exhaustion)**

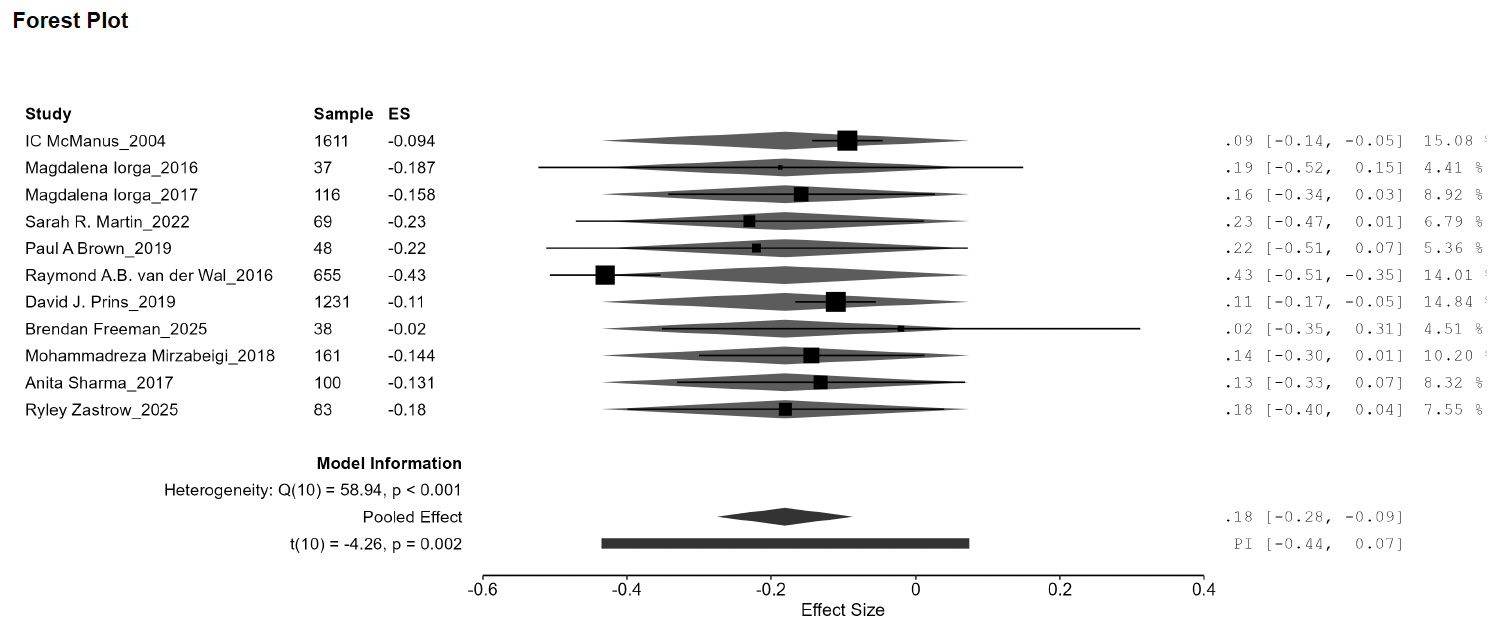

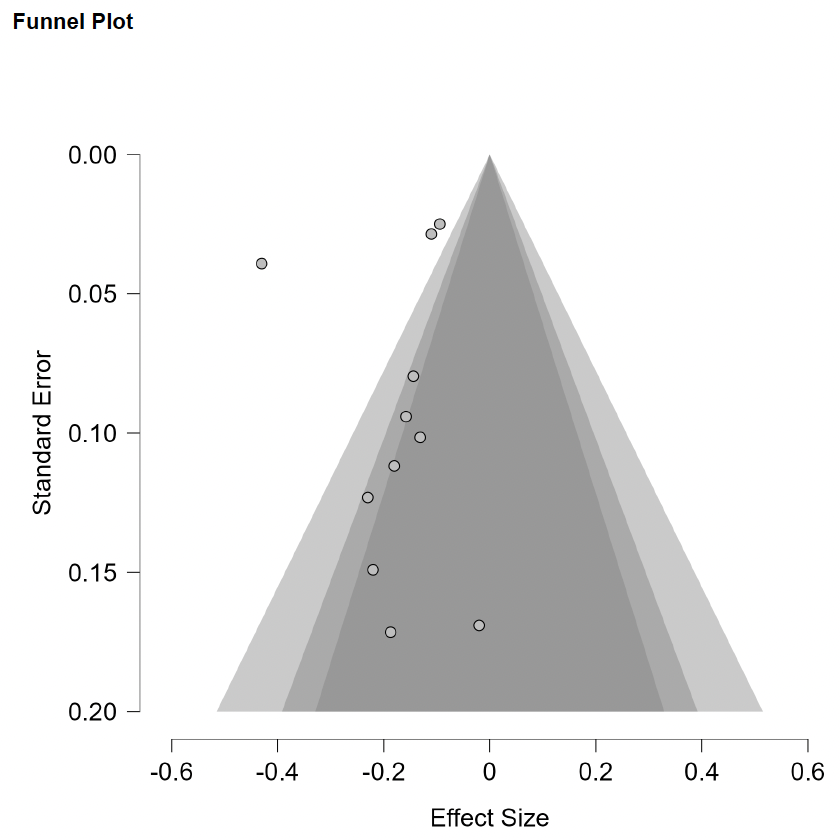

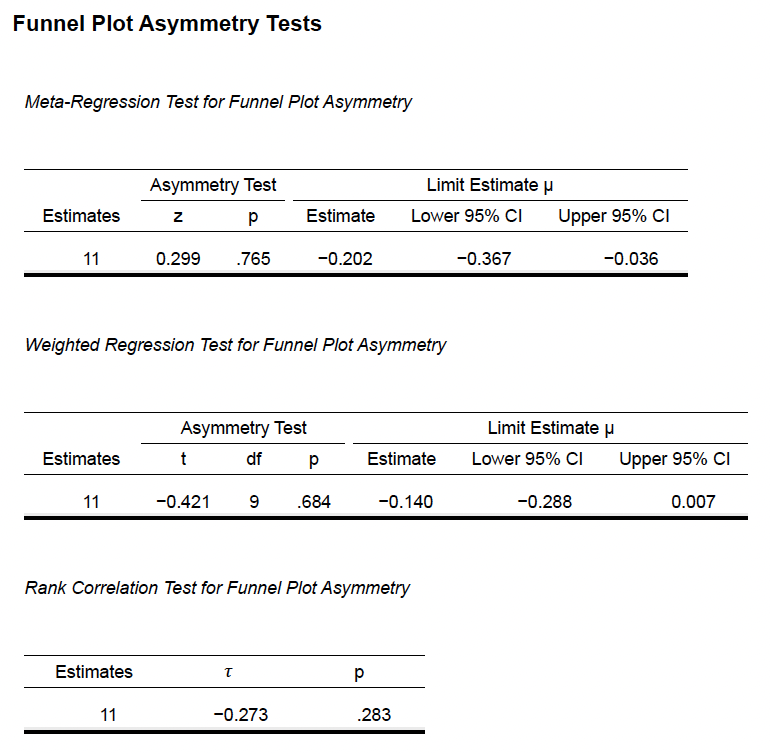

**Supplementary Methods 8b (Conscientiousness vs Emotional Exhaustion)**

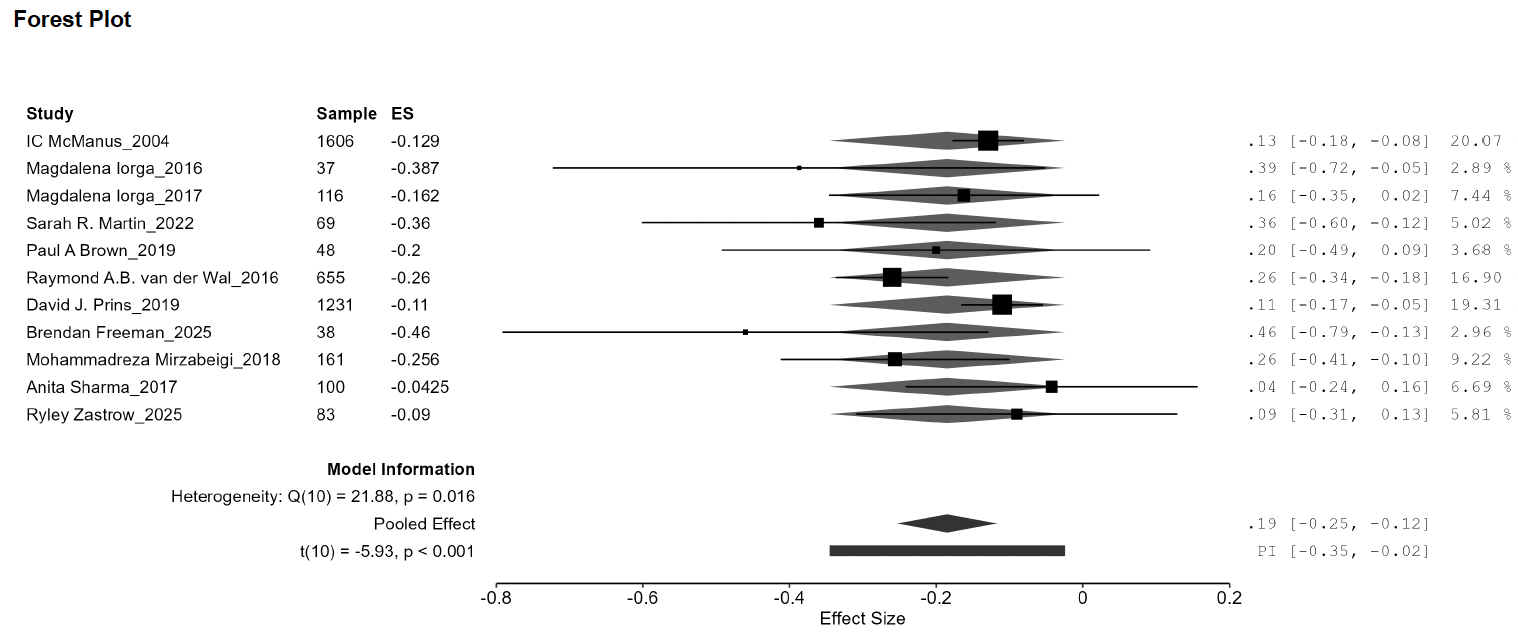

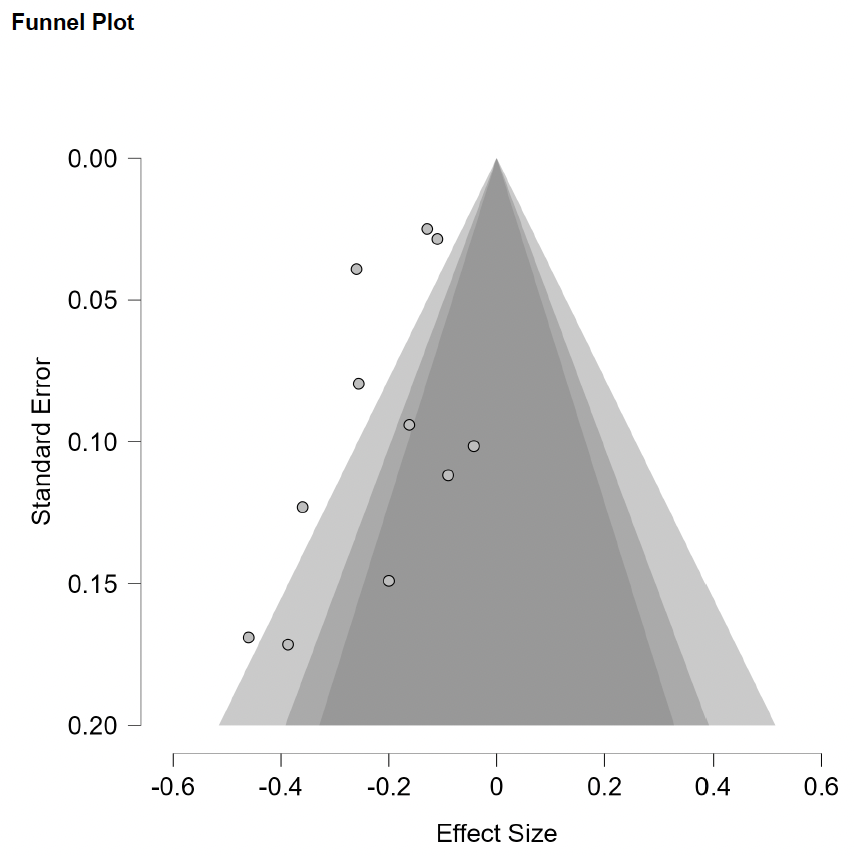

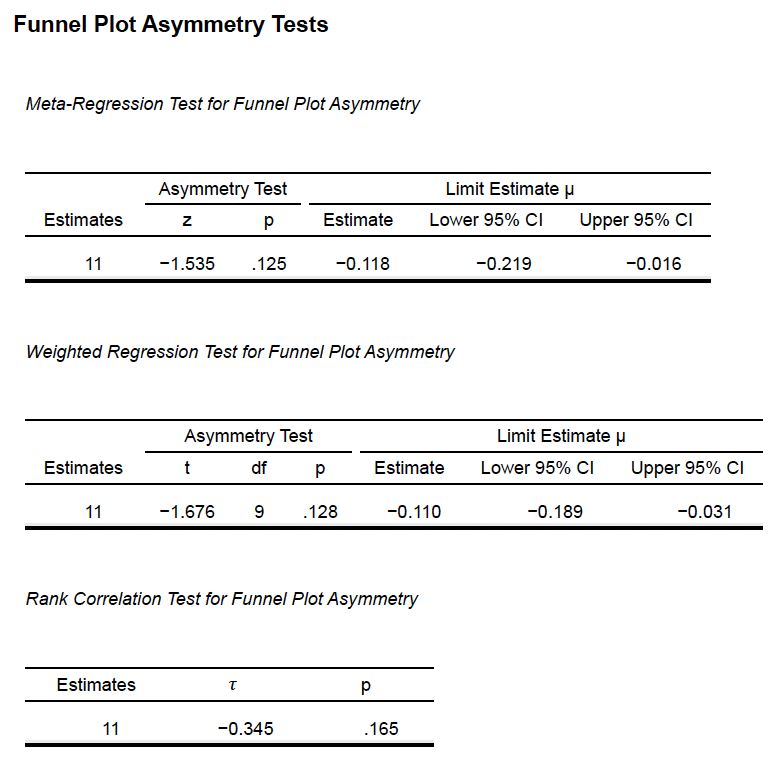

**Supplementary Methods 8c (Extraversion vs Emotional Exhaustion)**

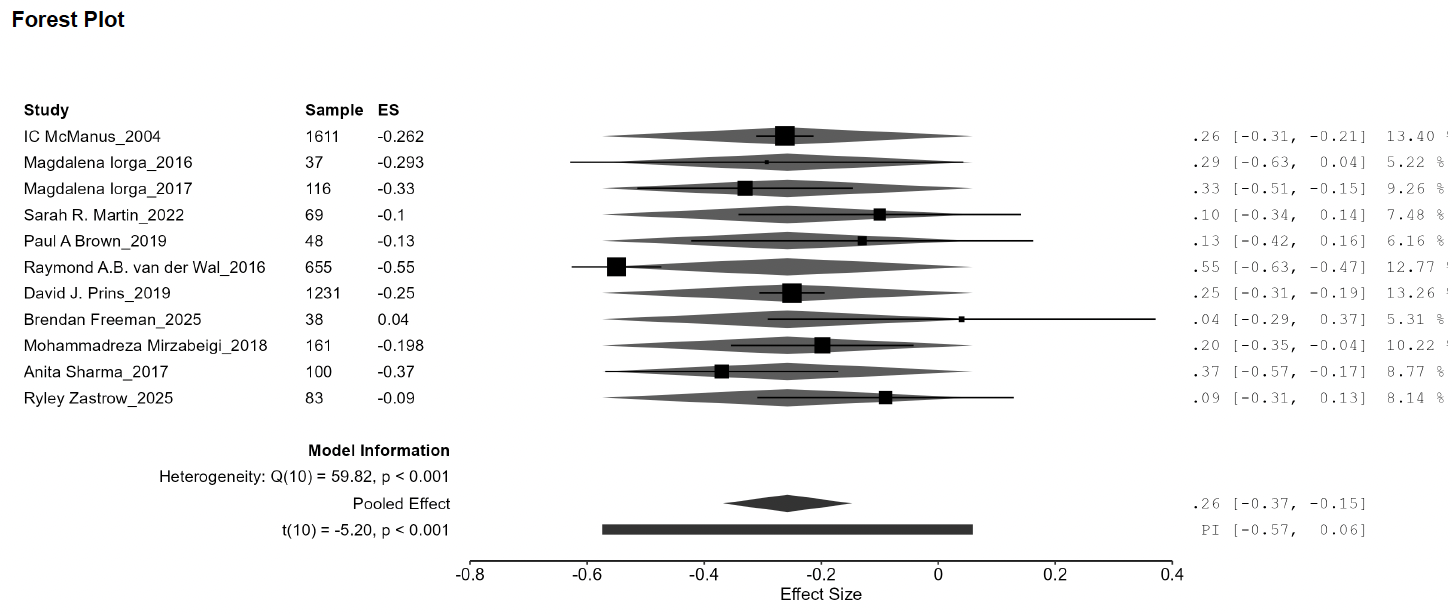

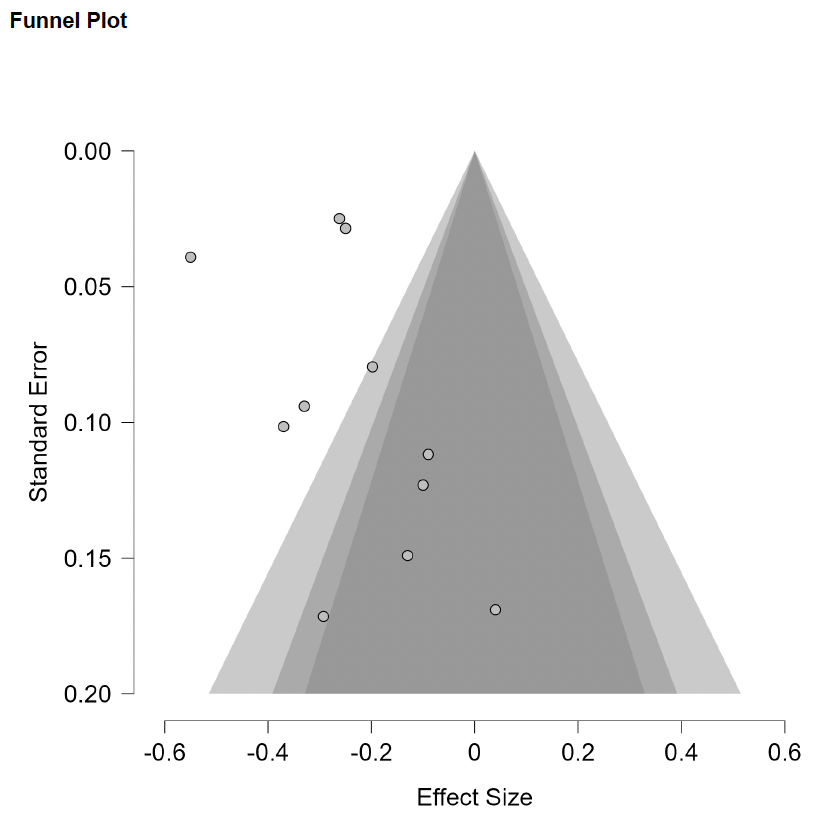

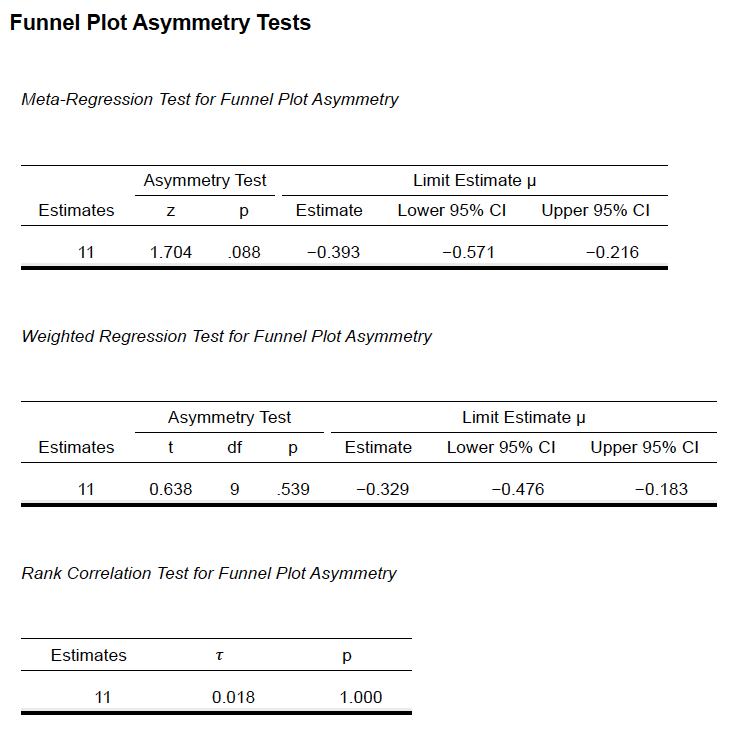

**Supplementary Methods 8d (Neuroticism vs Emotional Exhaustion)**

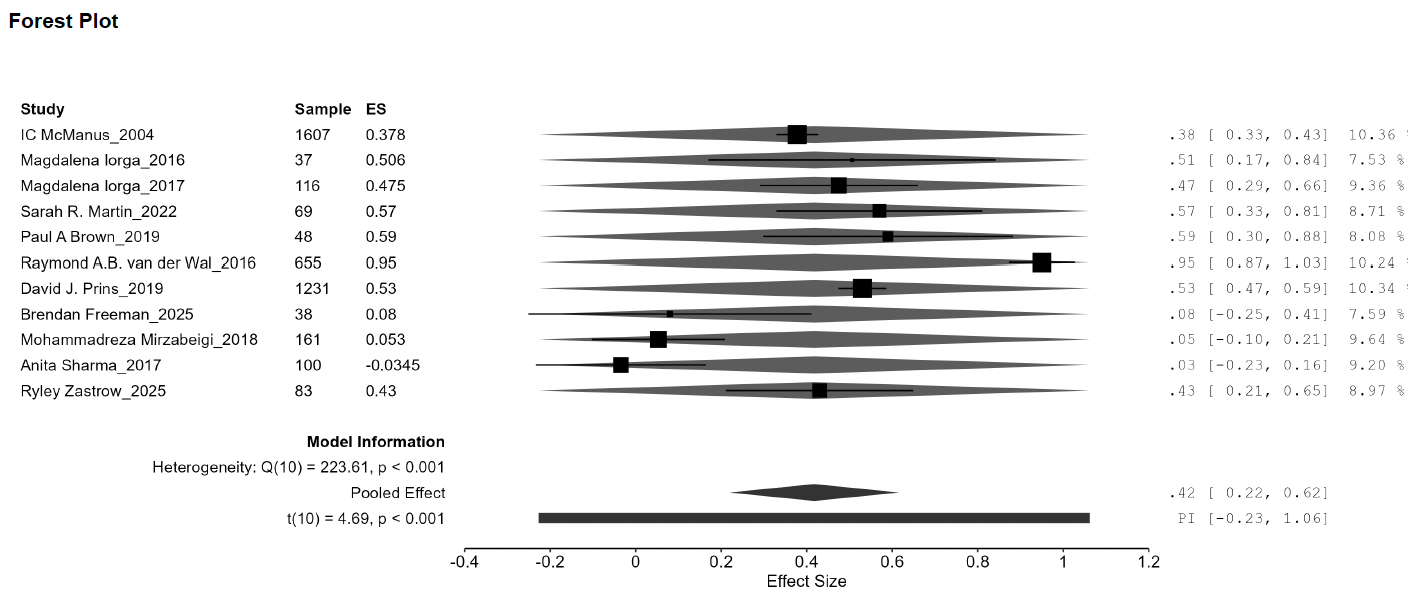

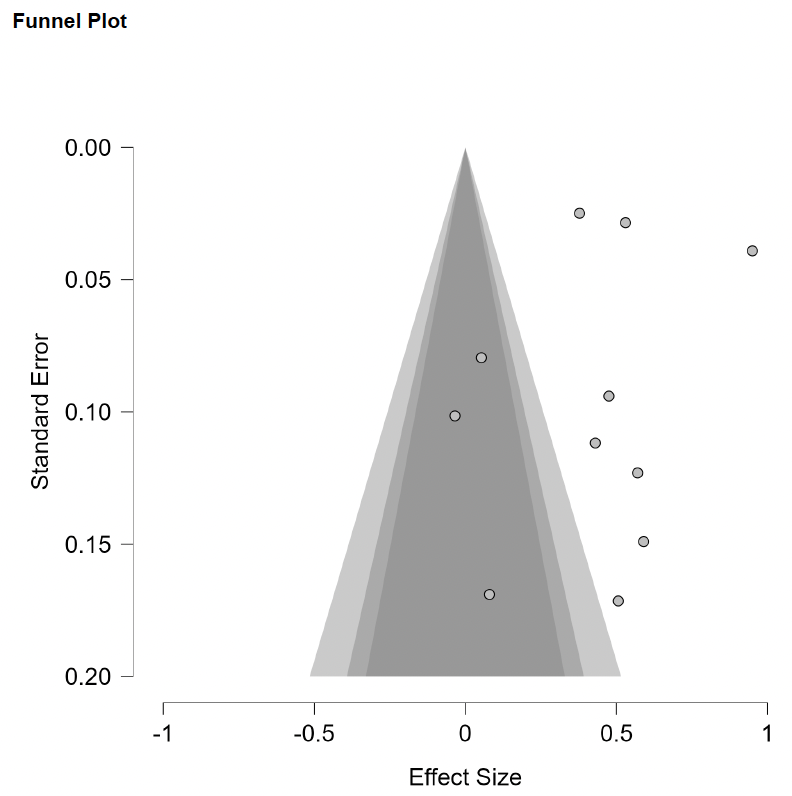

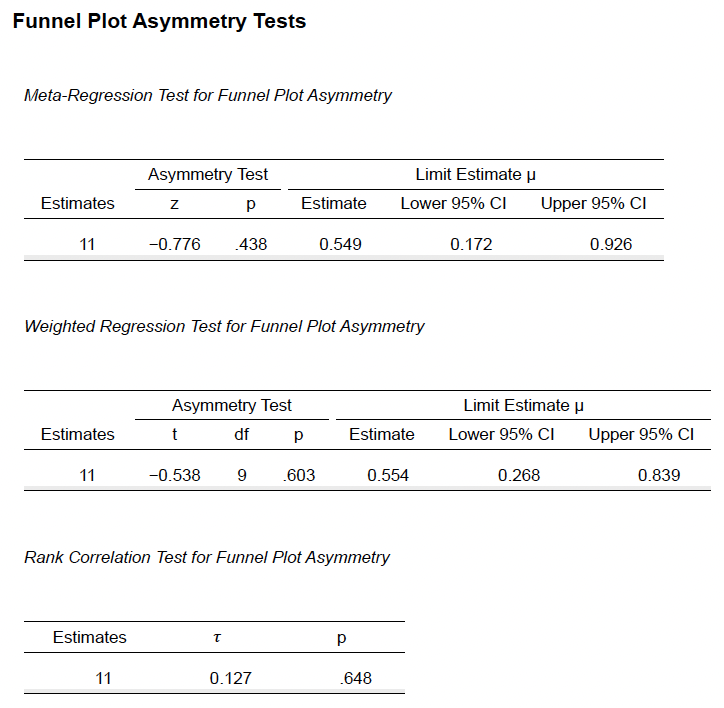

**Supplementary Methods 8e (Openness vs Emotional Exhaustion)**

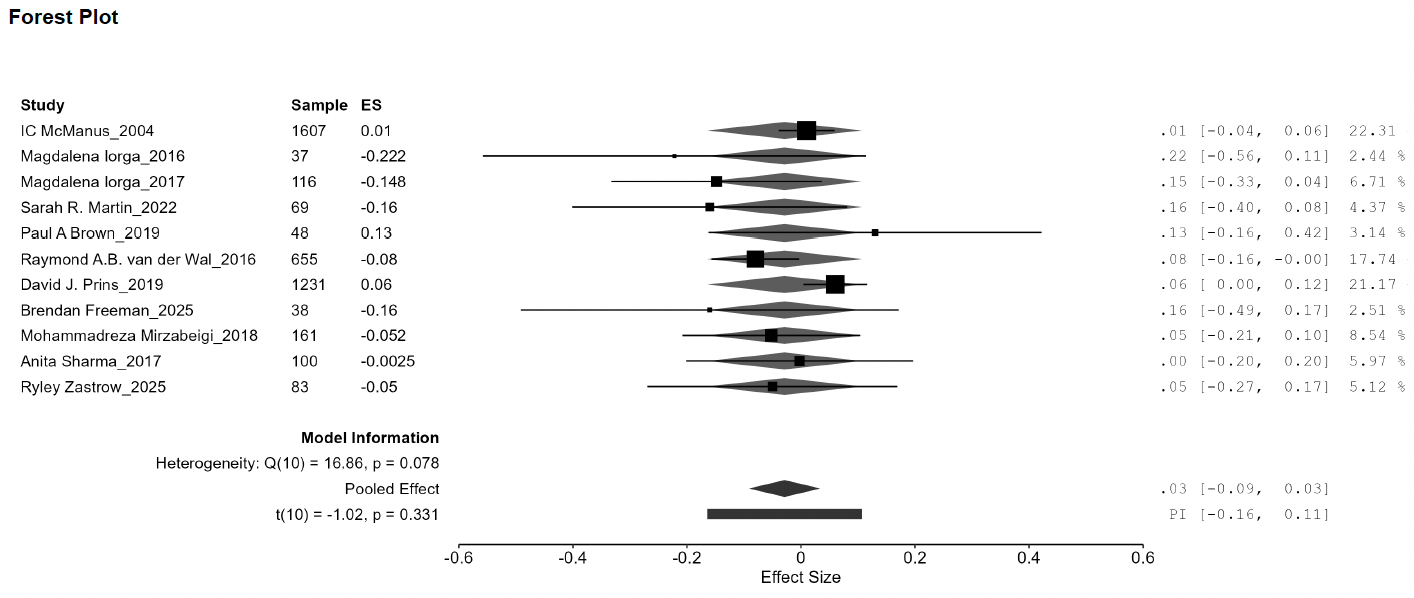

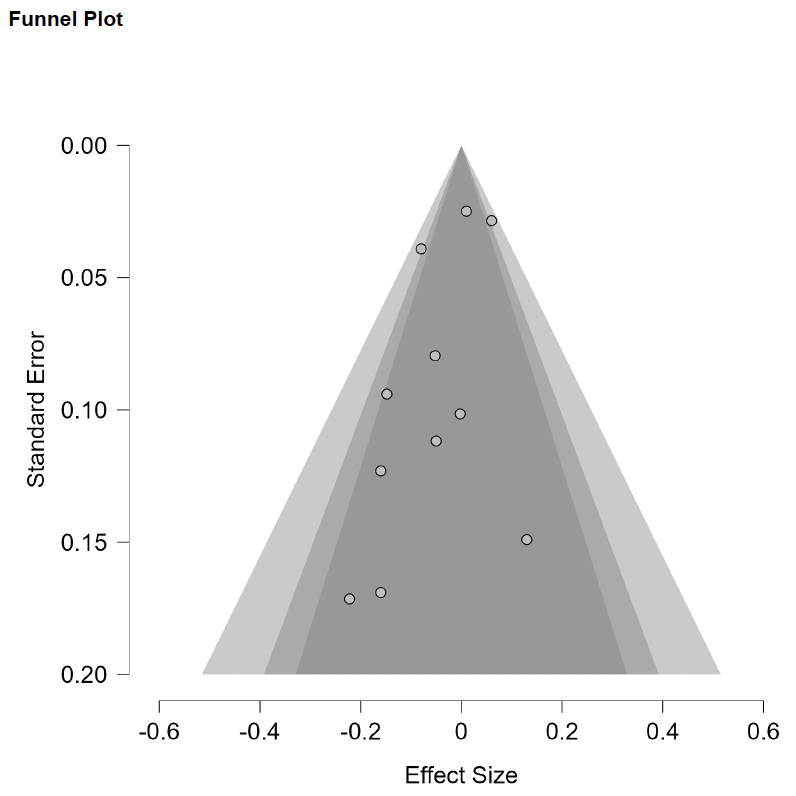

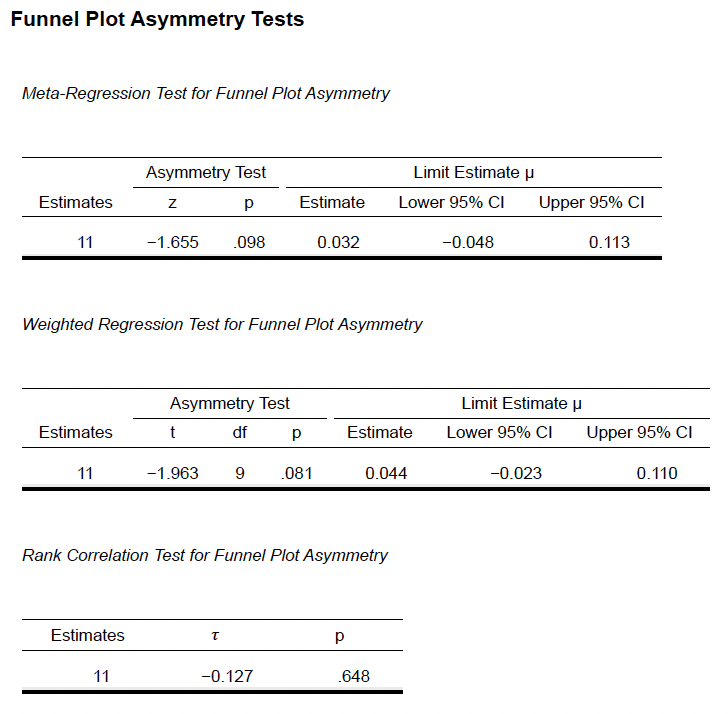

**Supplementary Methods 9a (Agreeableness vs Personal Accomplishment)**

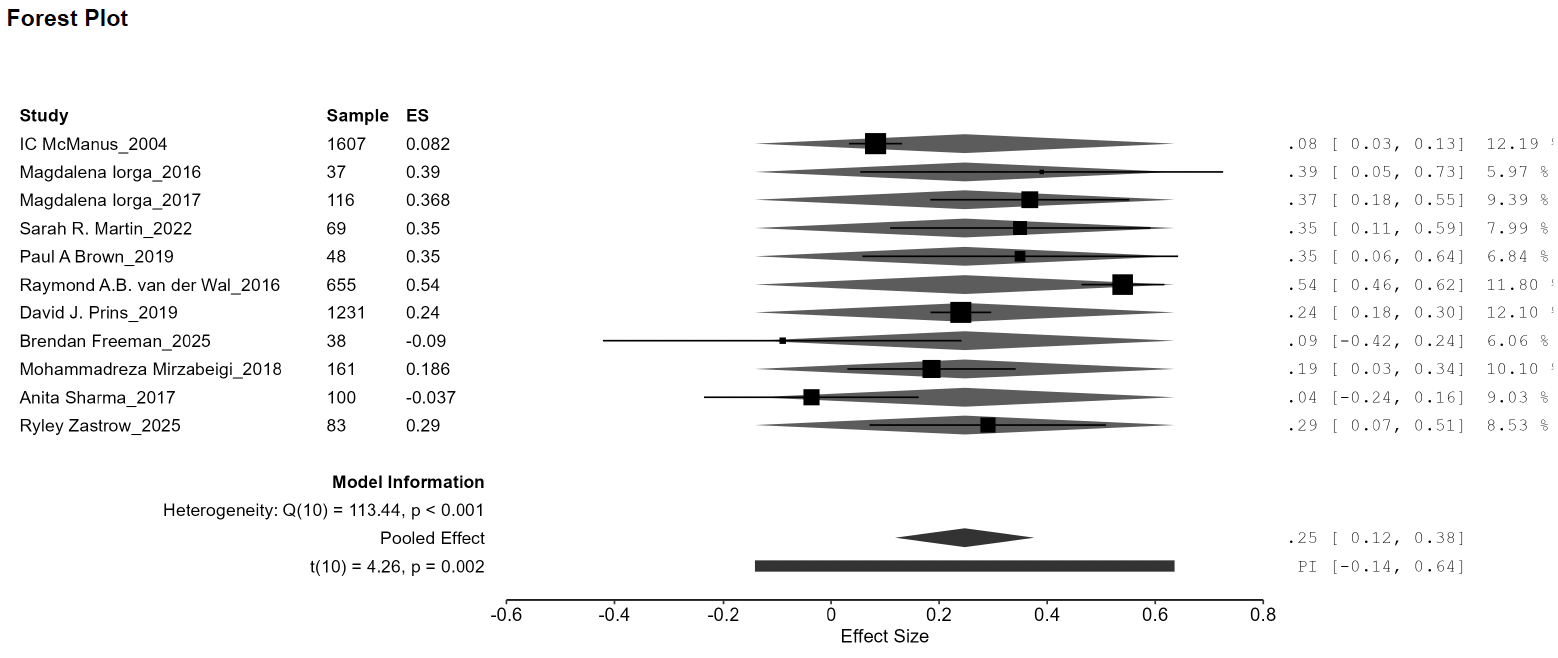

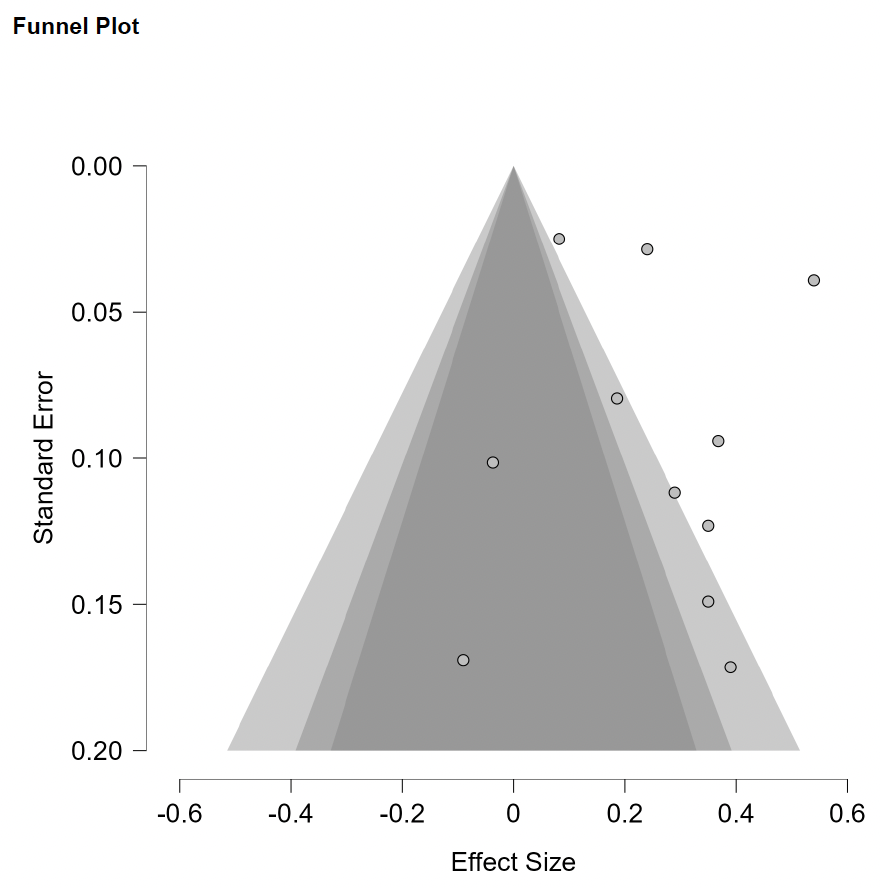

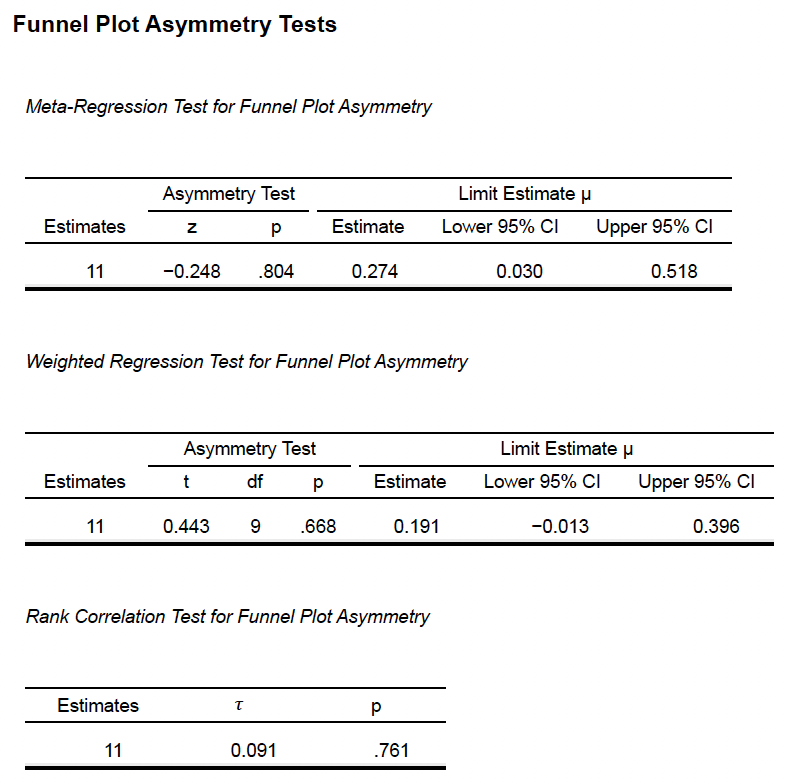

**Supplementary Methods 9b (Conscientiousness vs Personal Accomplishment)**

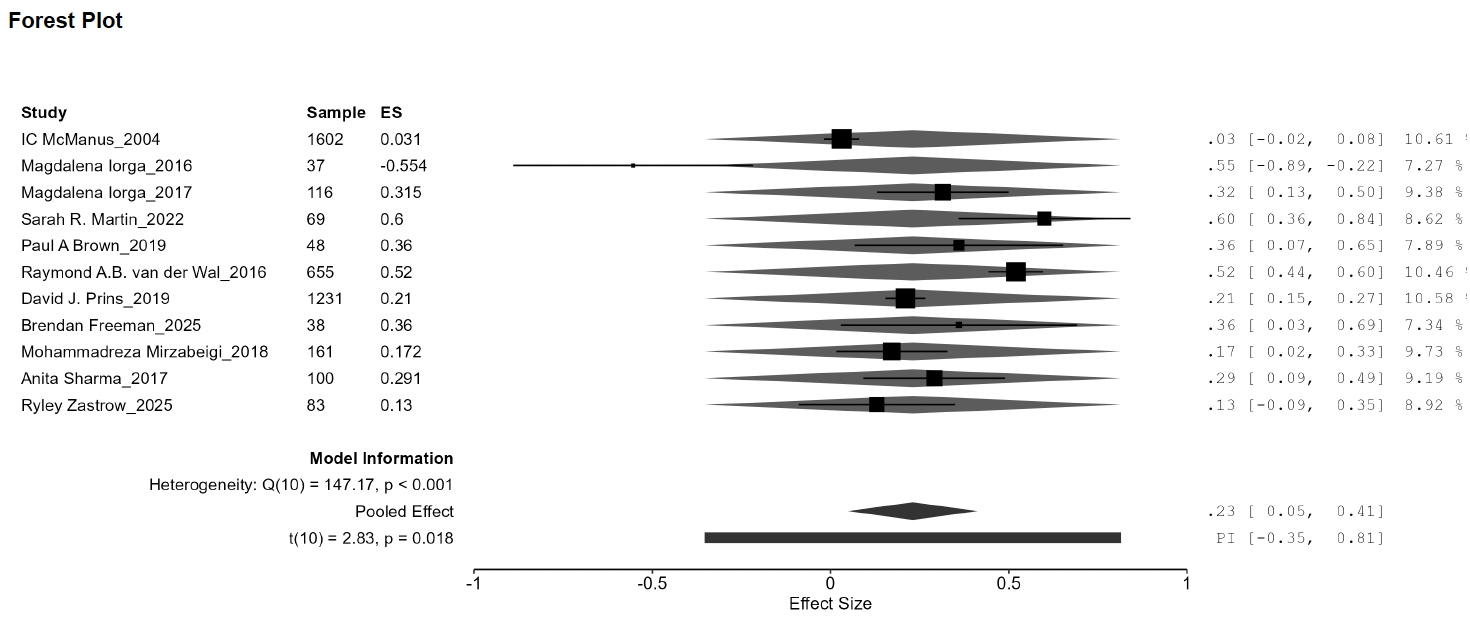

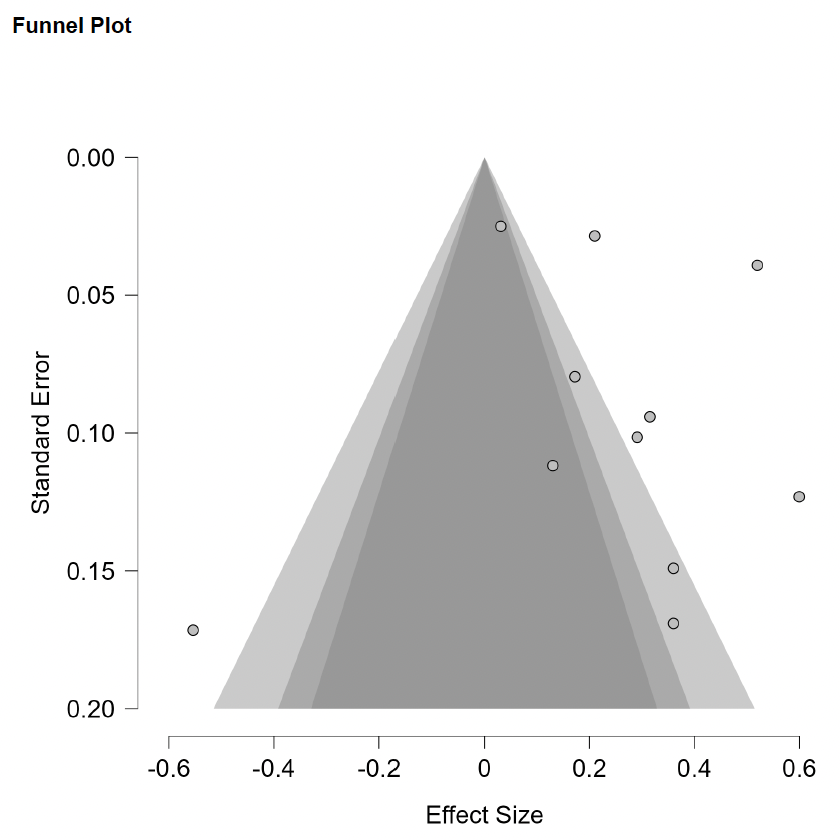

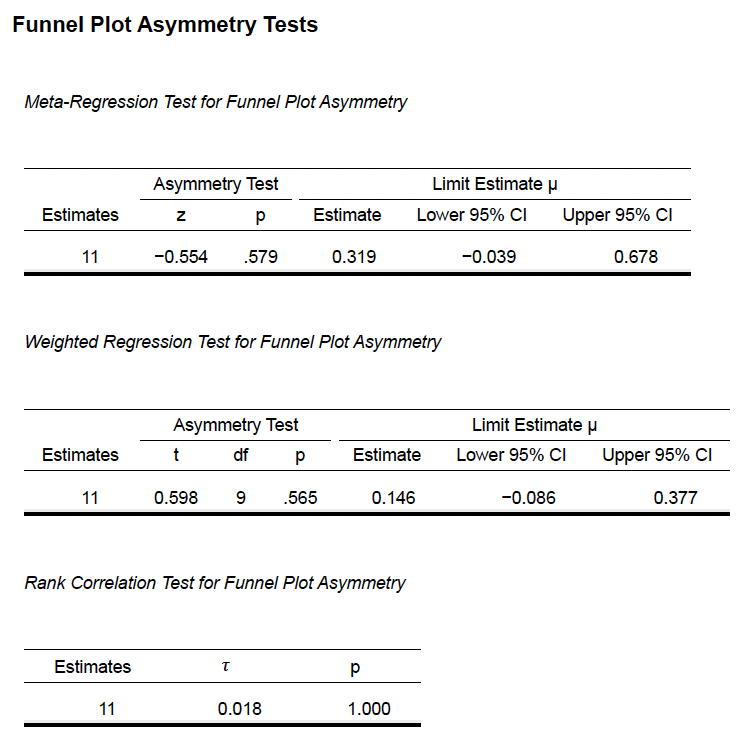

**Supplementary Methods 9c (Extraversion vs Personal Accomplishment)**

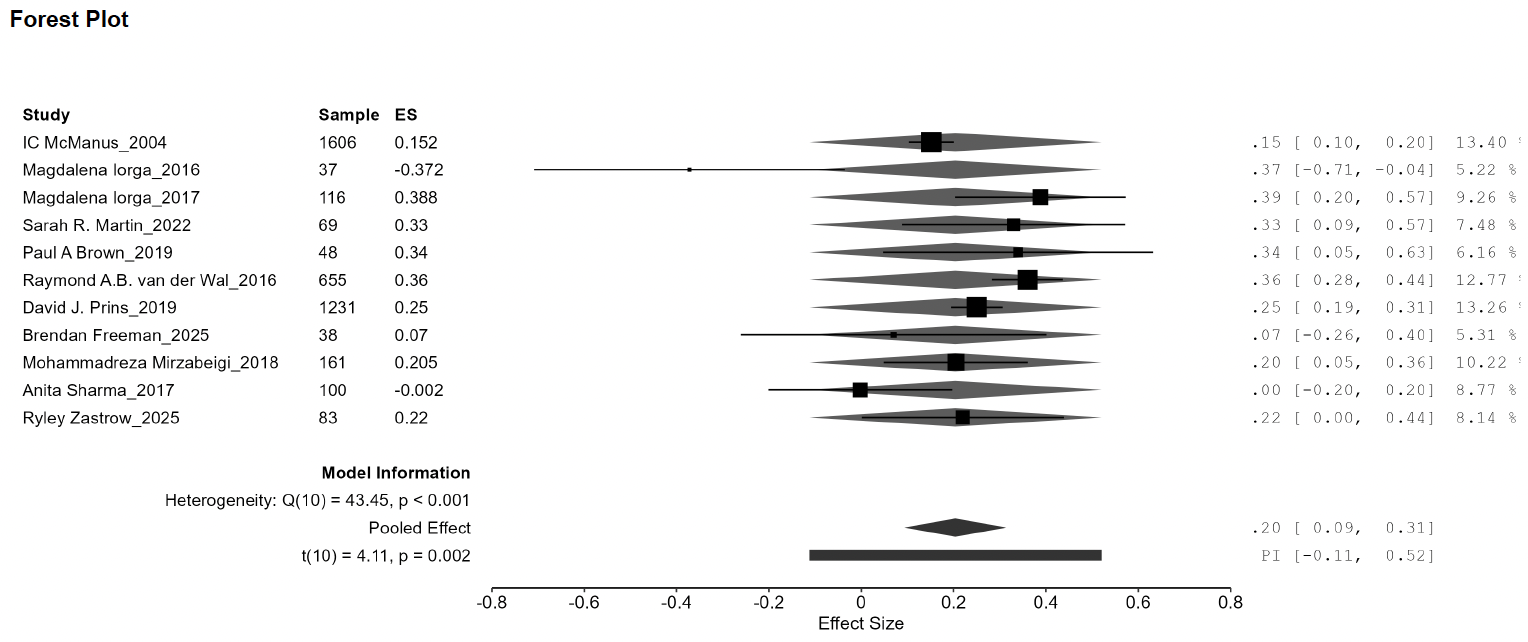

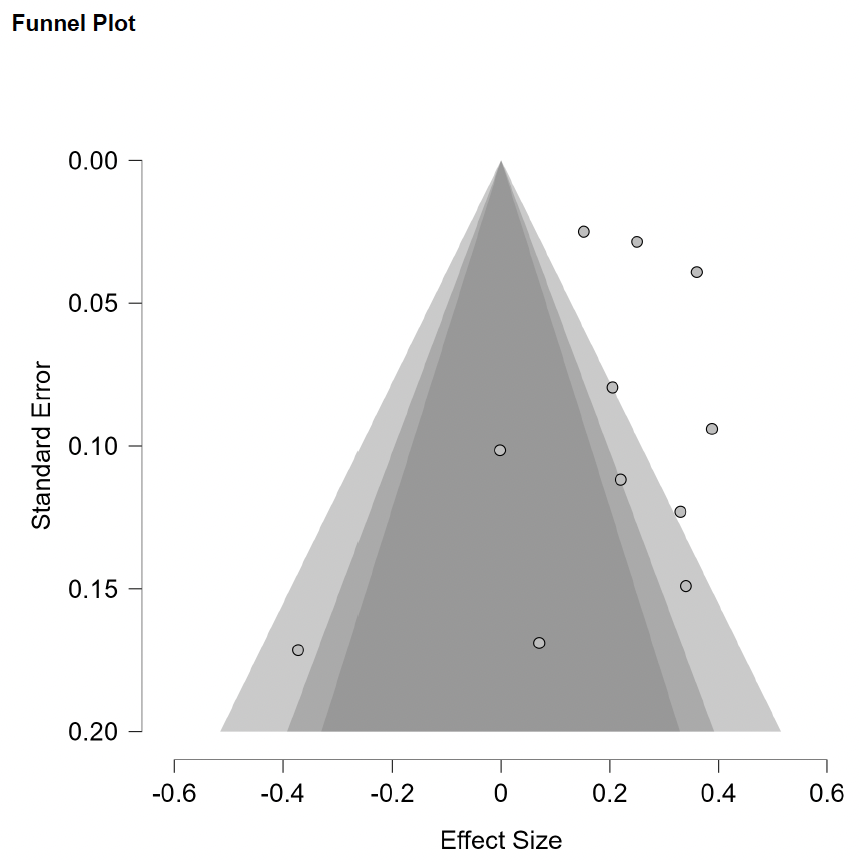

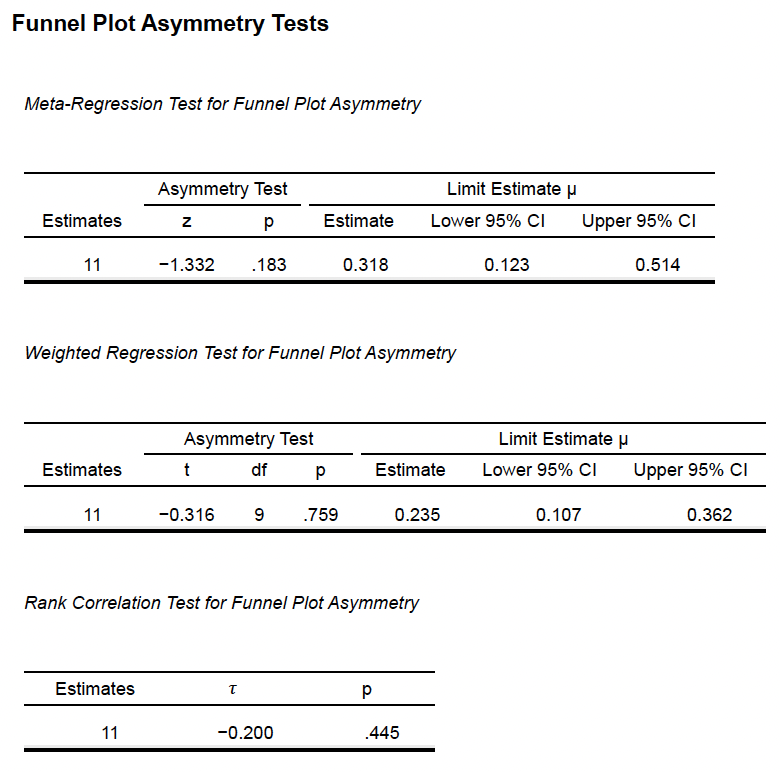

**Supplementary Methods 9d (Neuroticism vs Personal Accomplishment)**

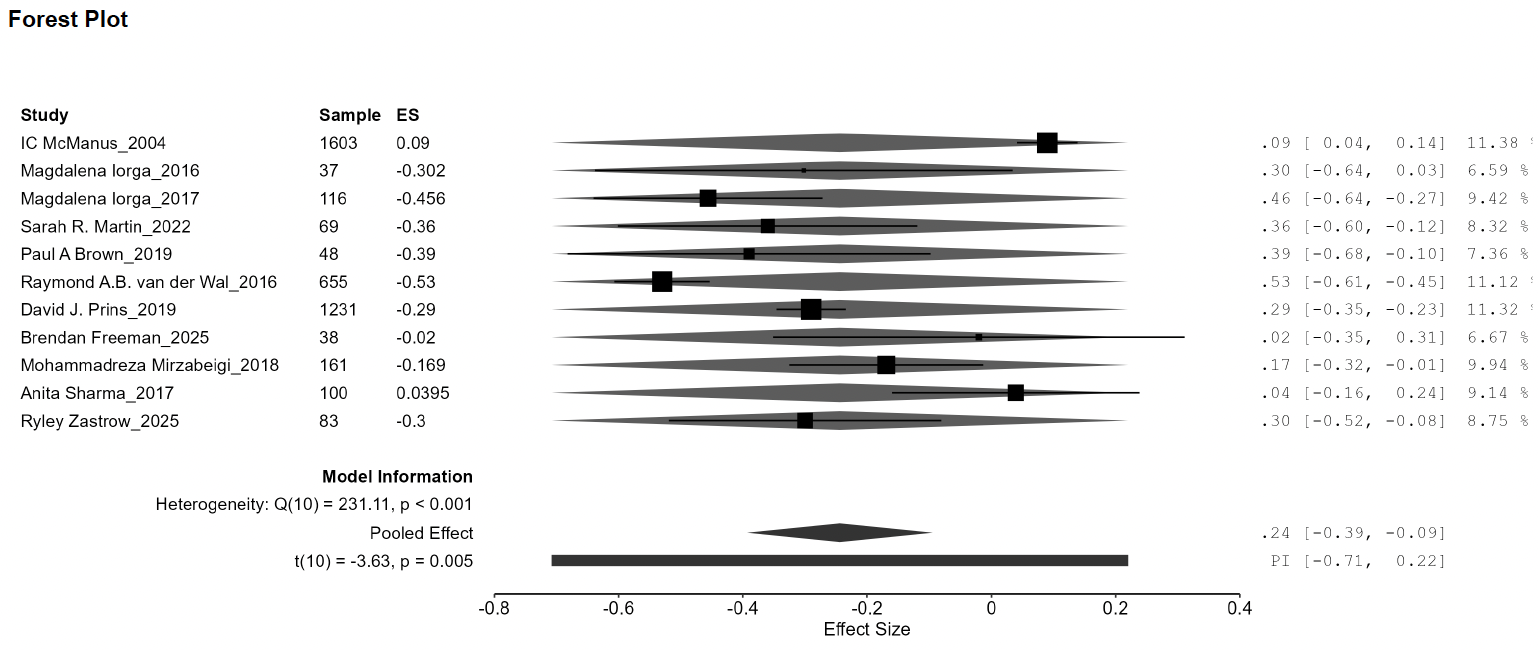

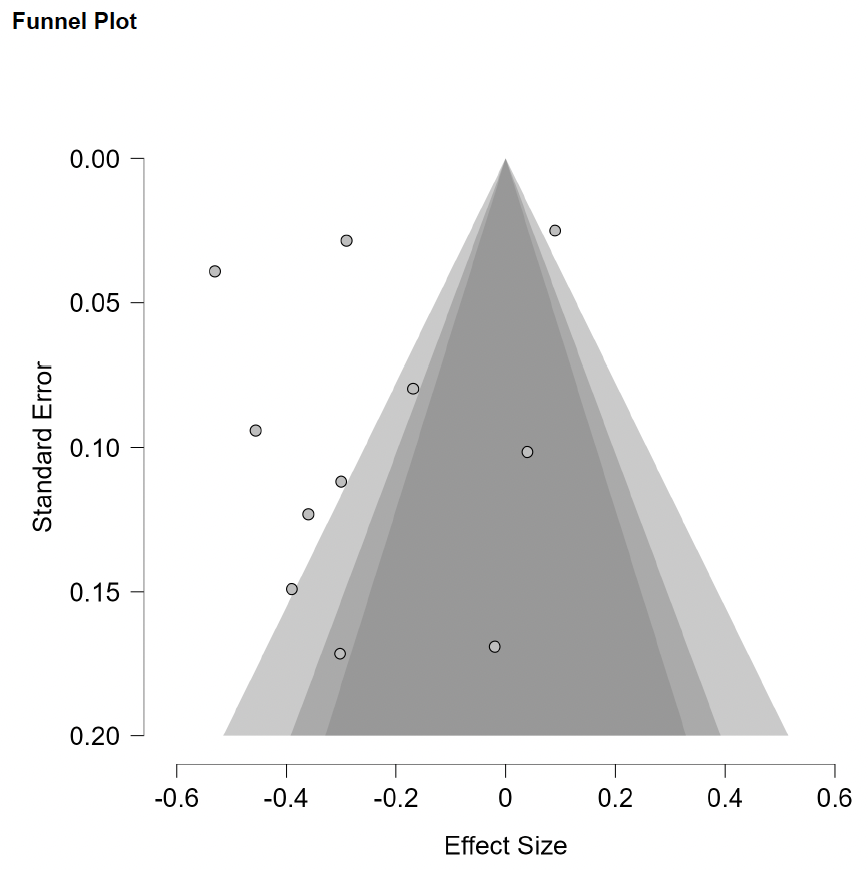

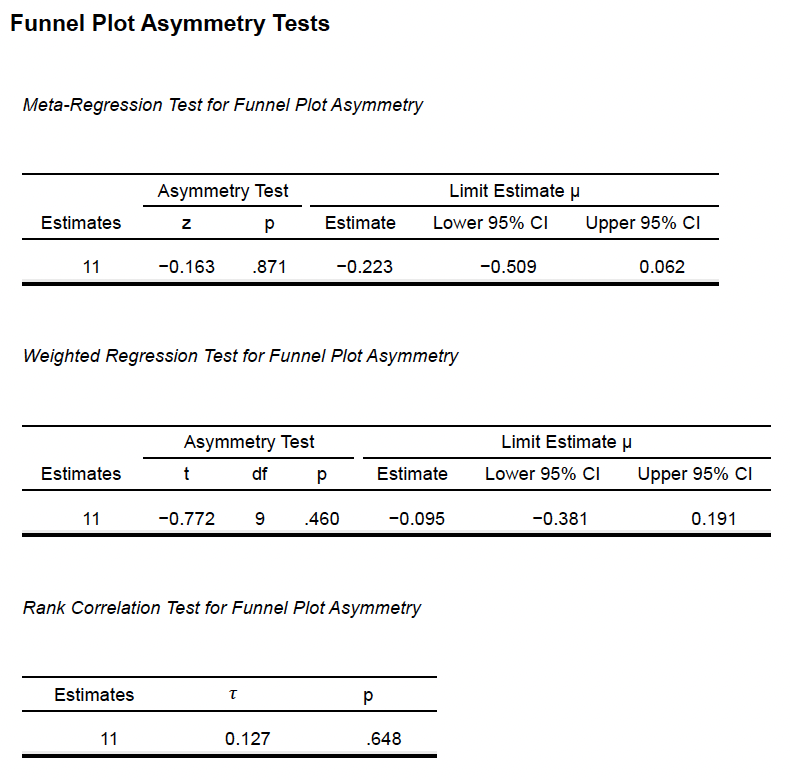

**Supplementary Methods 9e (Openness vs Personal Accomplishment)**

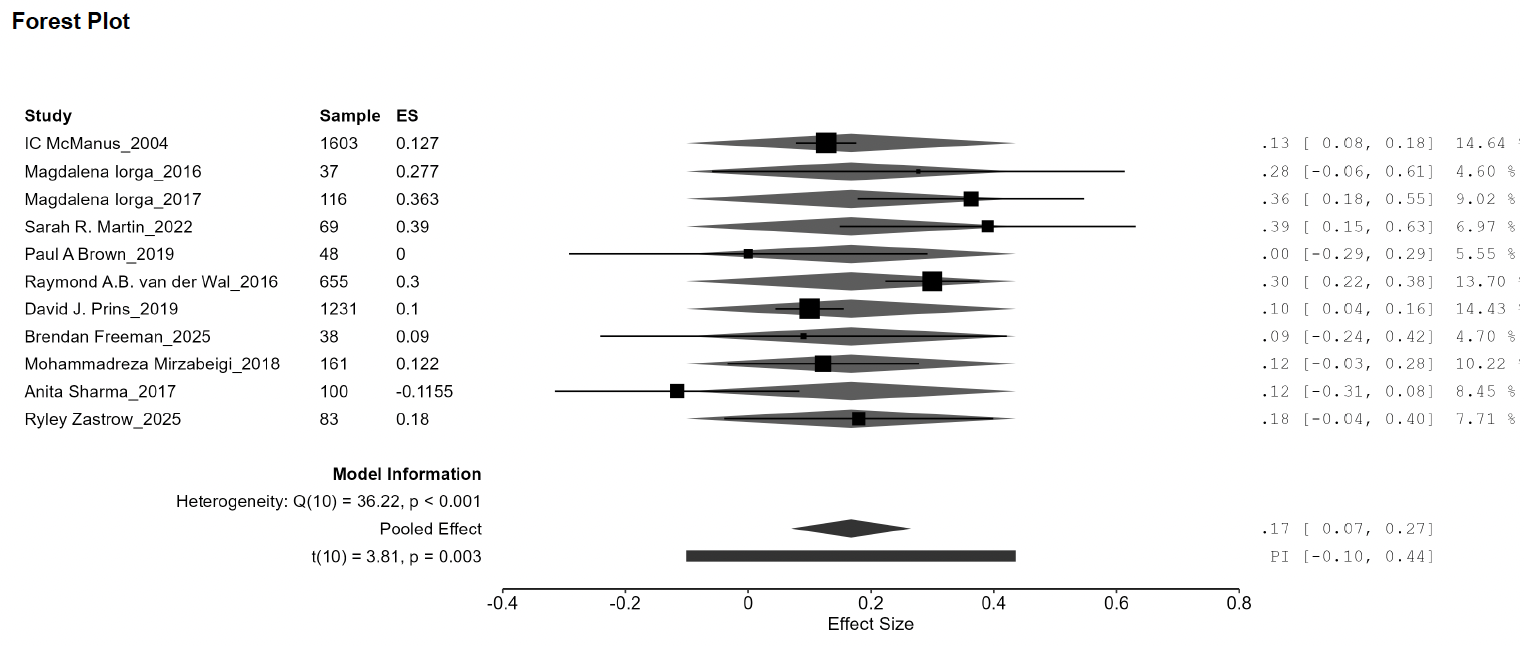

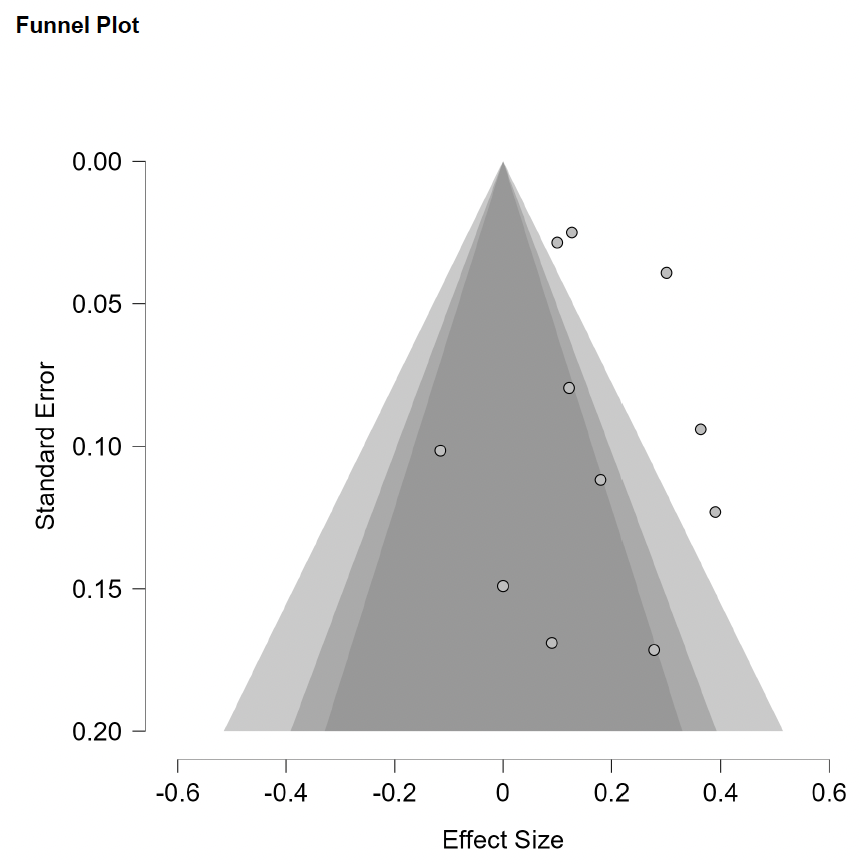

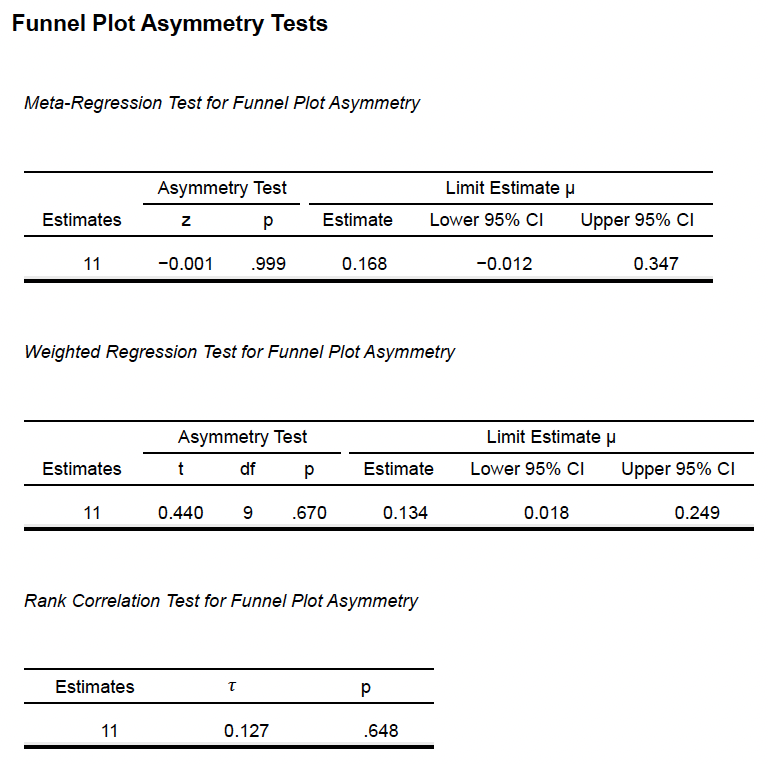

**Supplementary Methods 10a (Agreeableness vs Stress)**

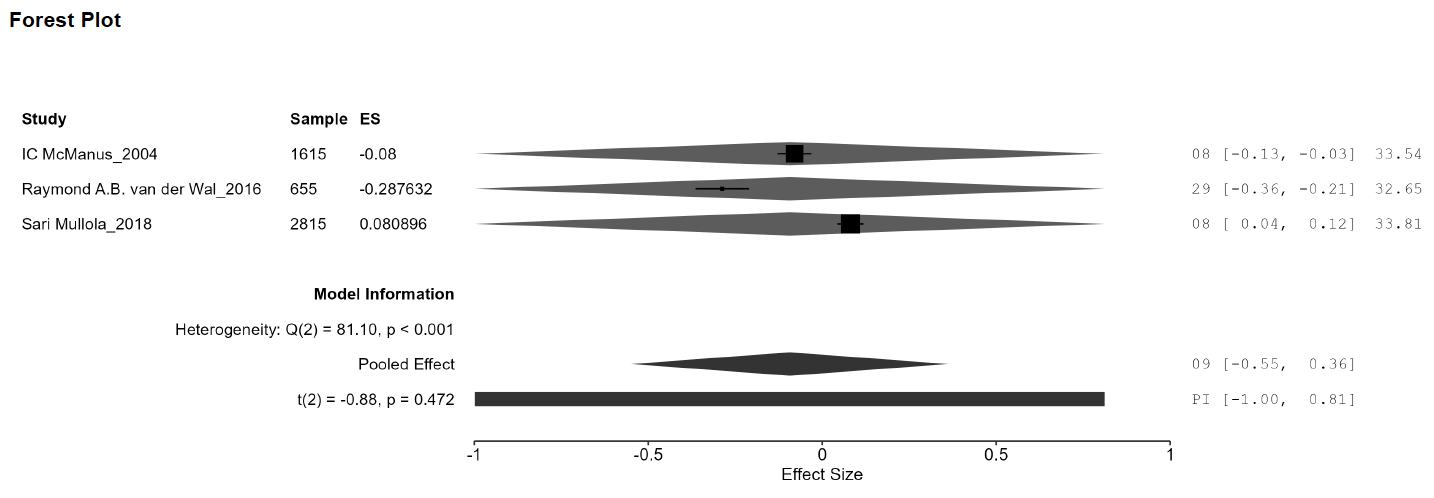

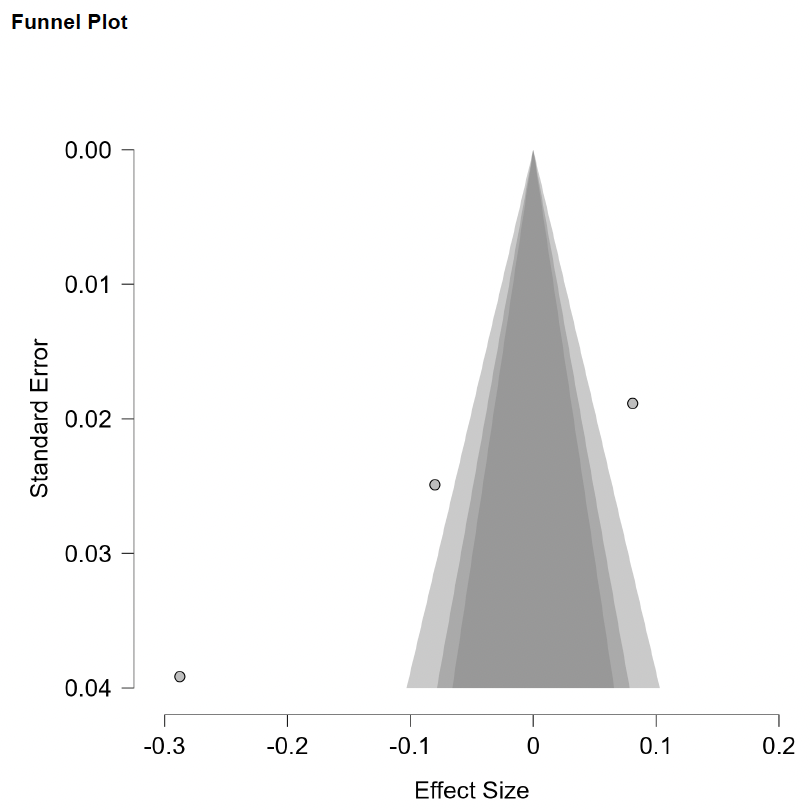

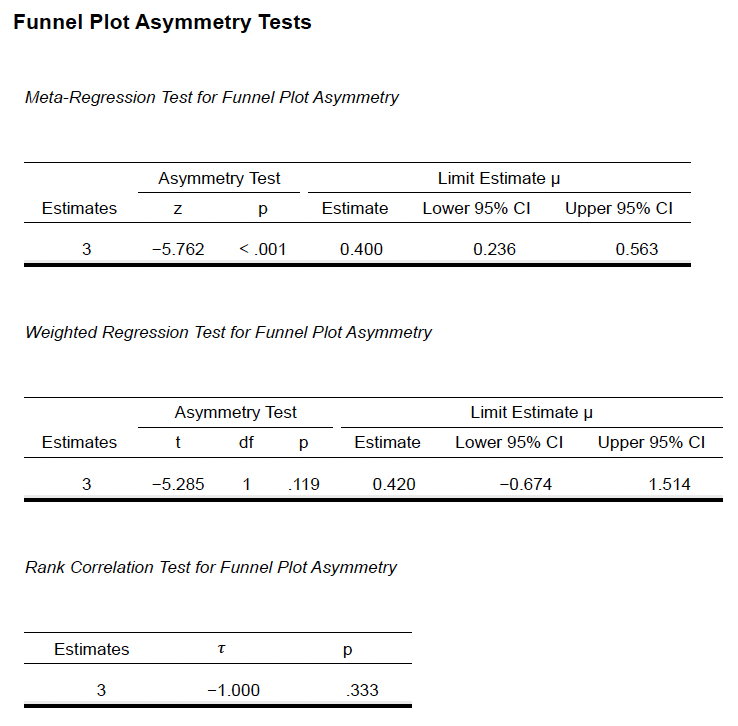

**Supplementary Methods 10b (Conscientiousness vs Stress)**

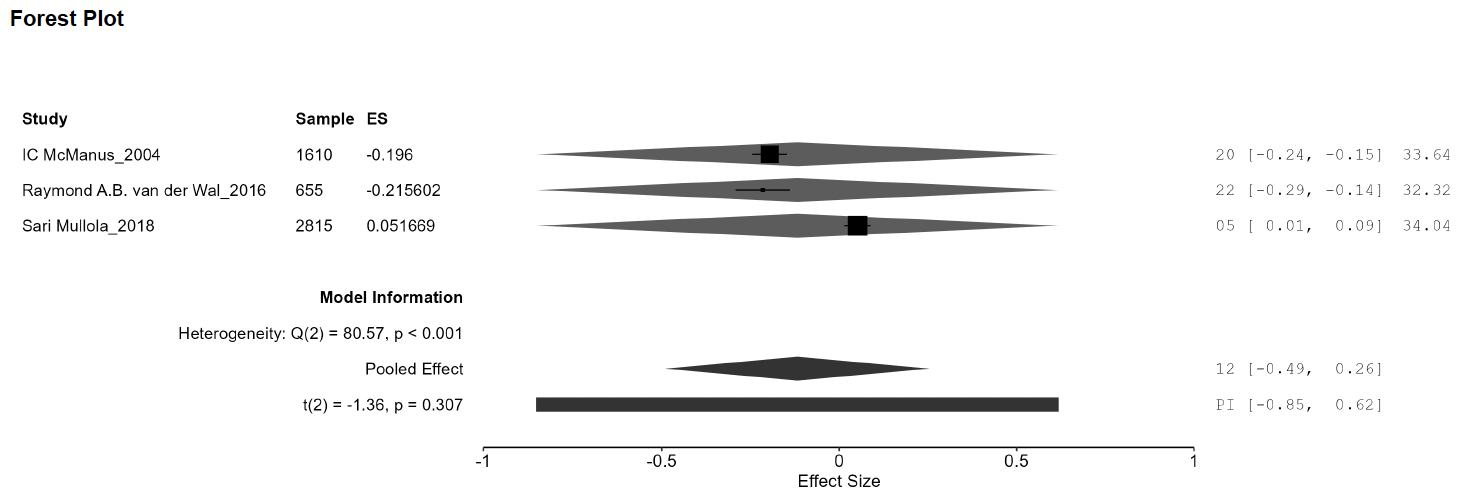

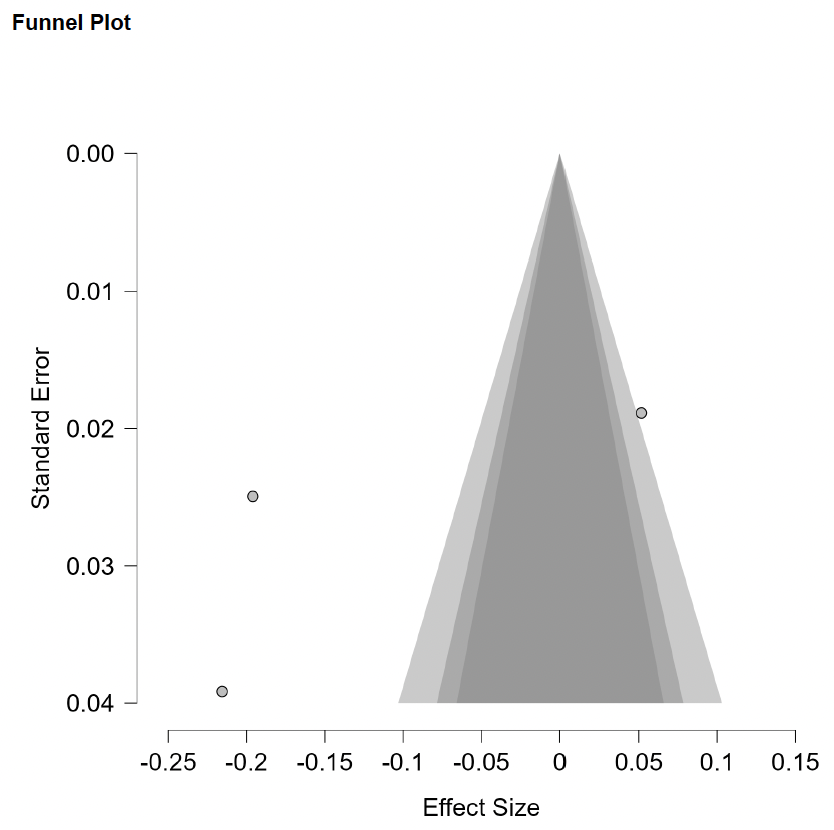

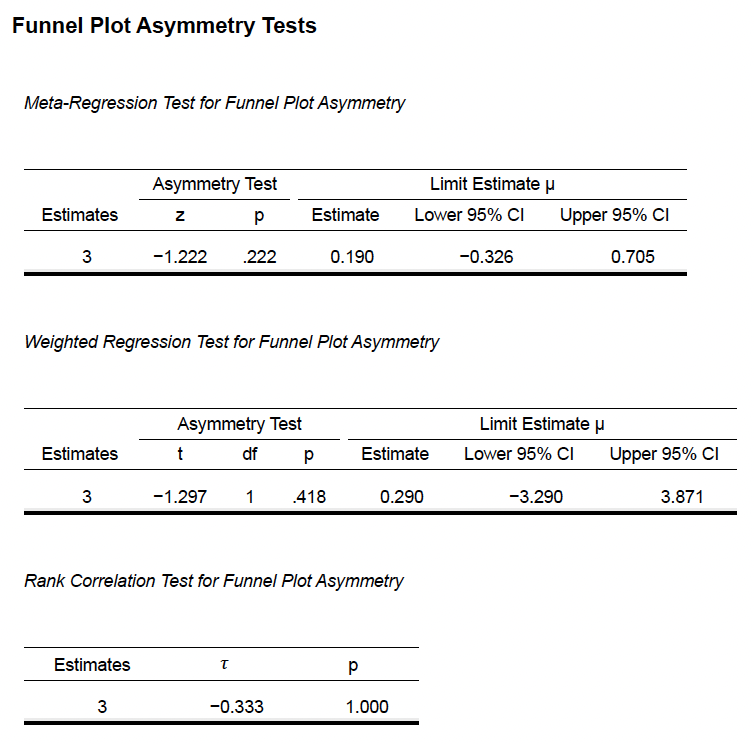

**Supplementary Methods 10c (Extraversion vs Stress)**

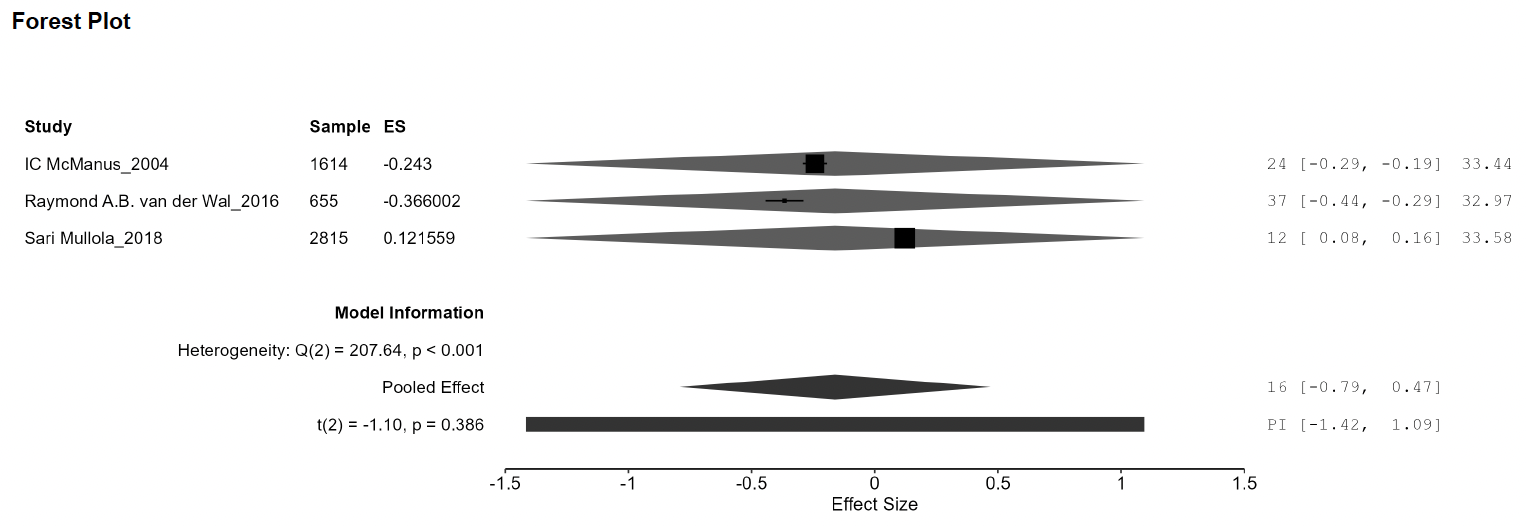

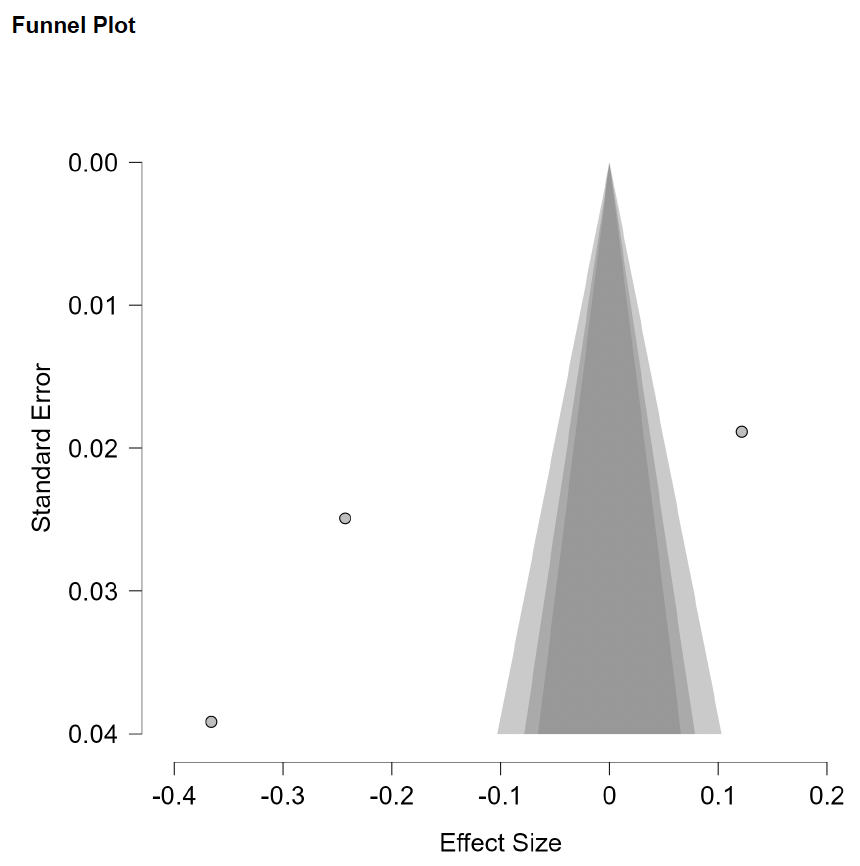

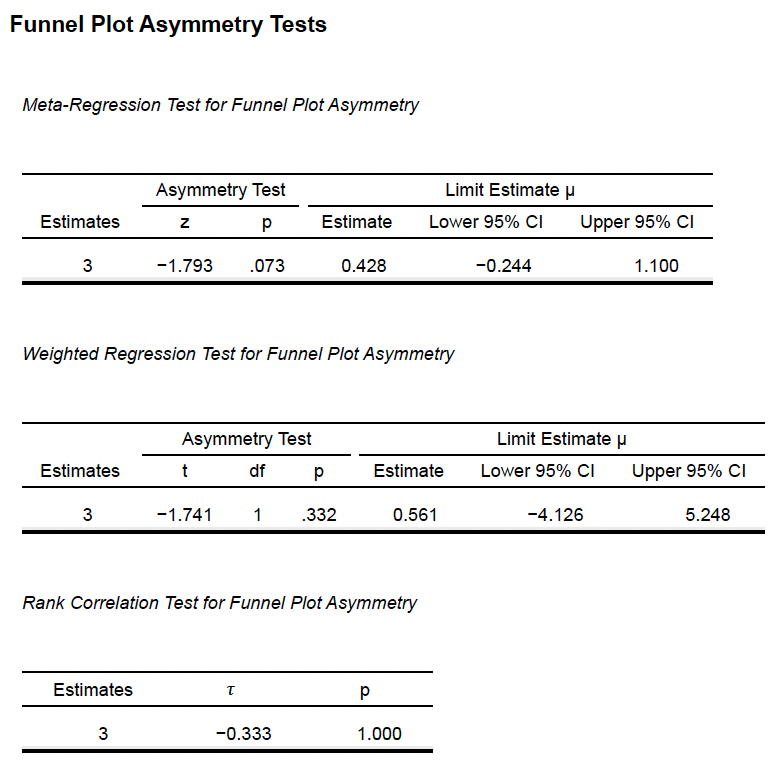

**Supplementary Methods 10d (Neuroticism vs Stress)**

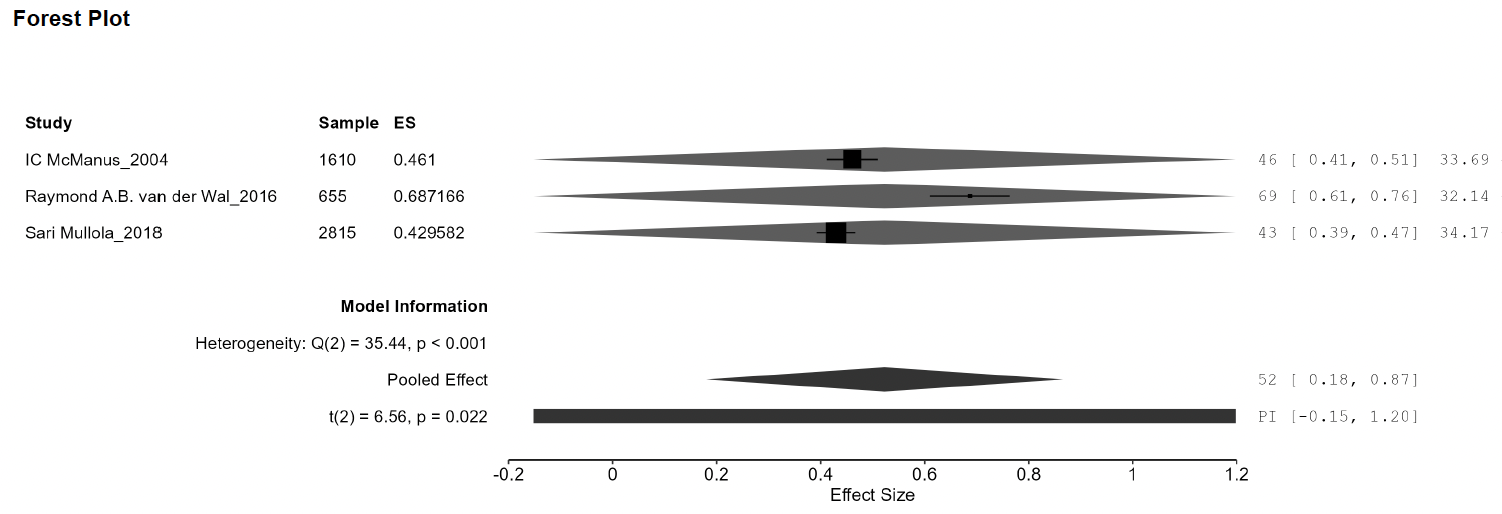

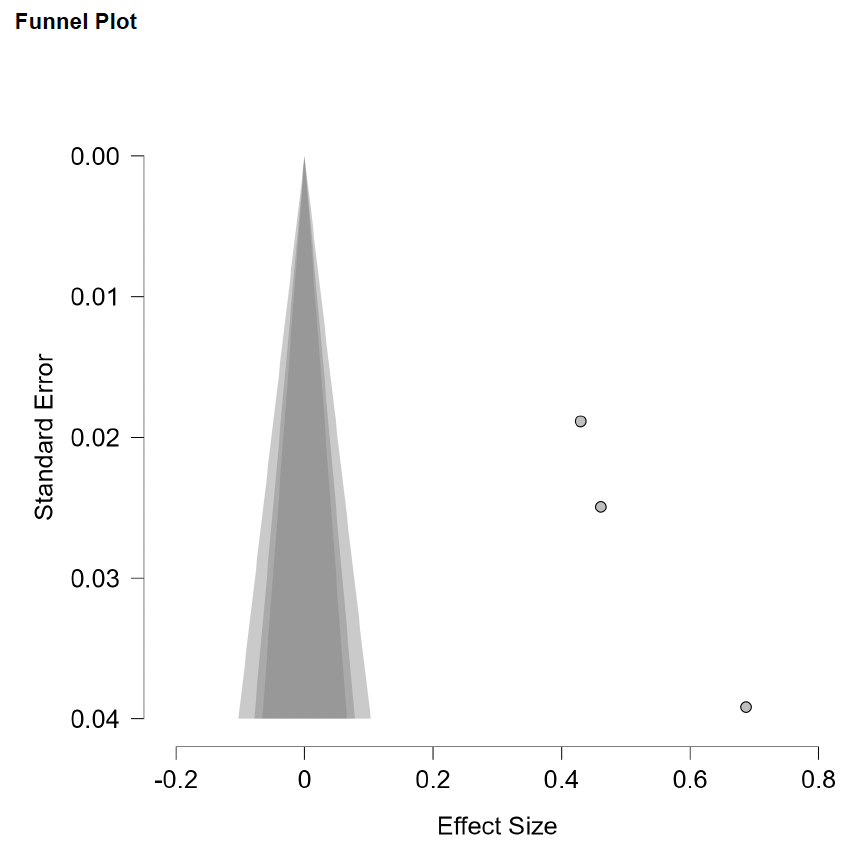

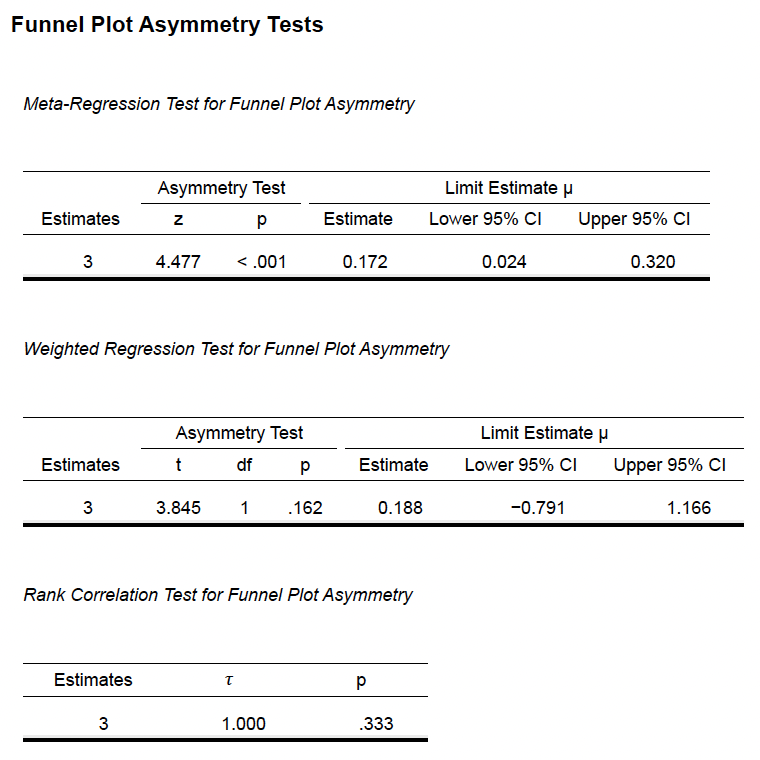

**Supplementary Methods 10e (Openness vs Stress)**

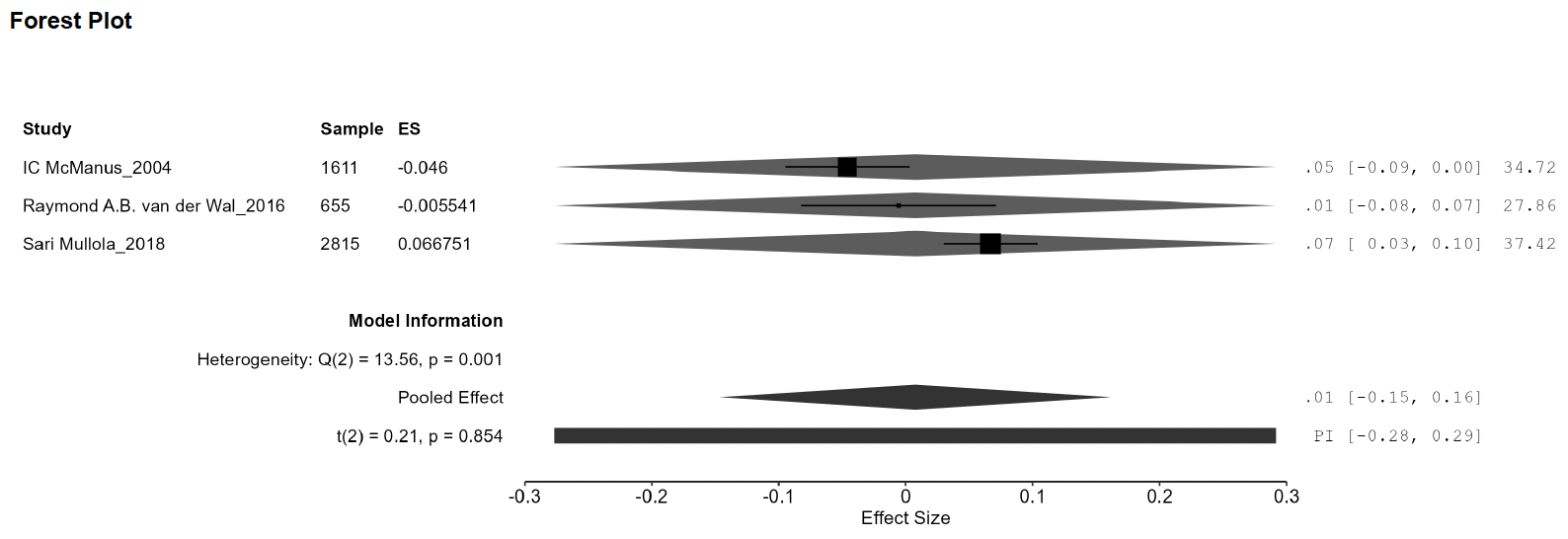

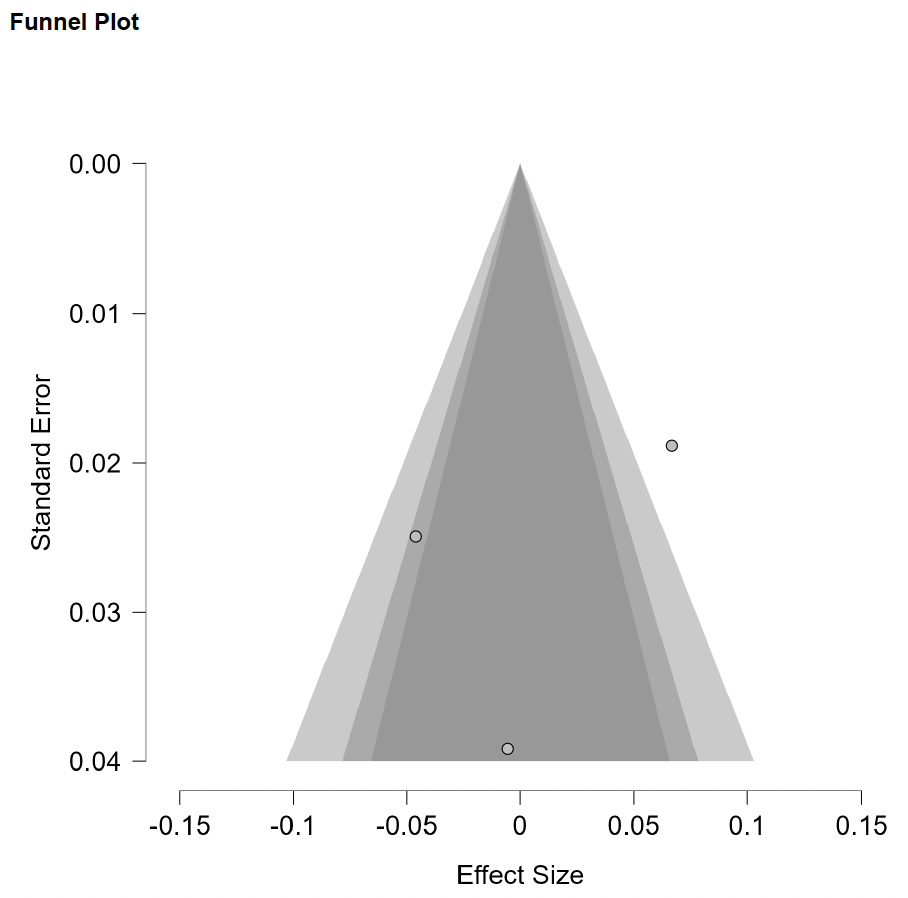

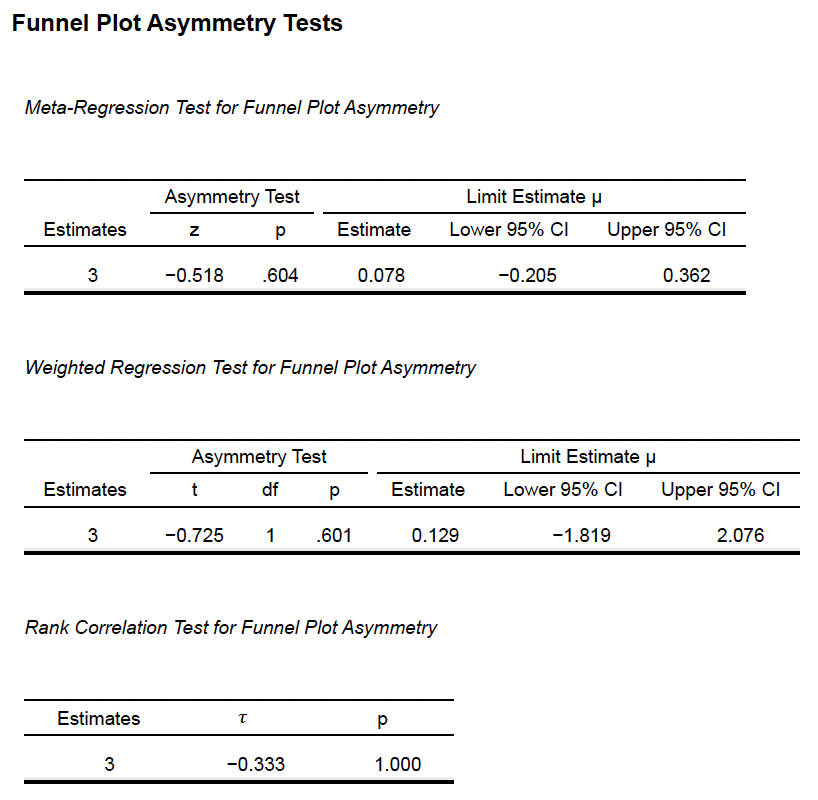

**Supplementary Methods 11a (Experience vs Agreeableness vs Depersonalization)**

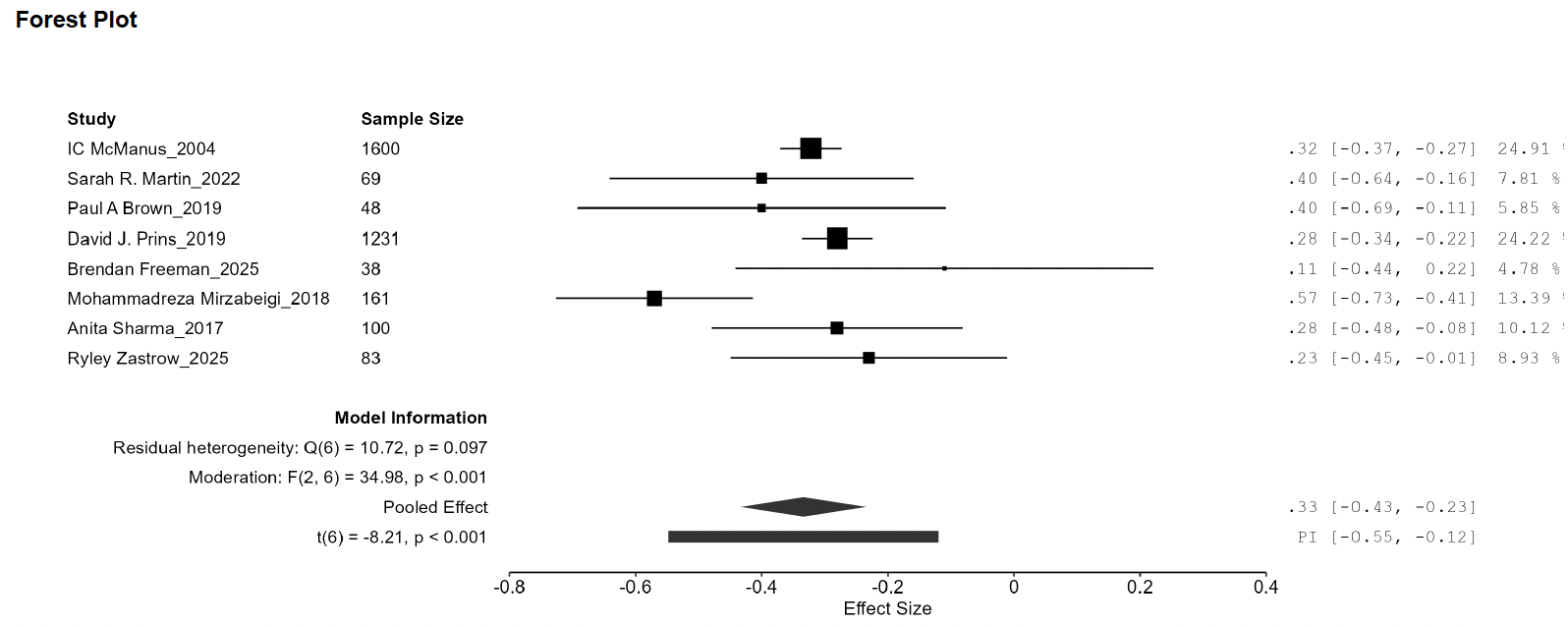

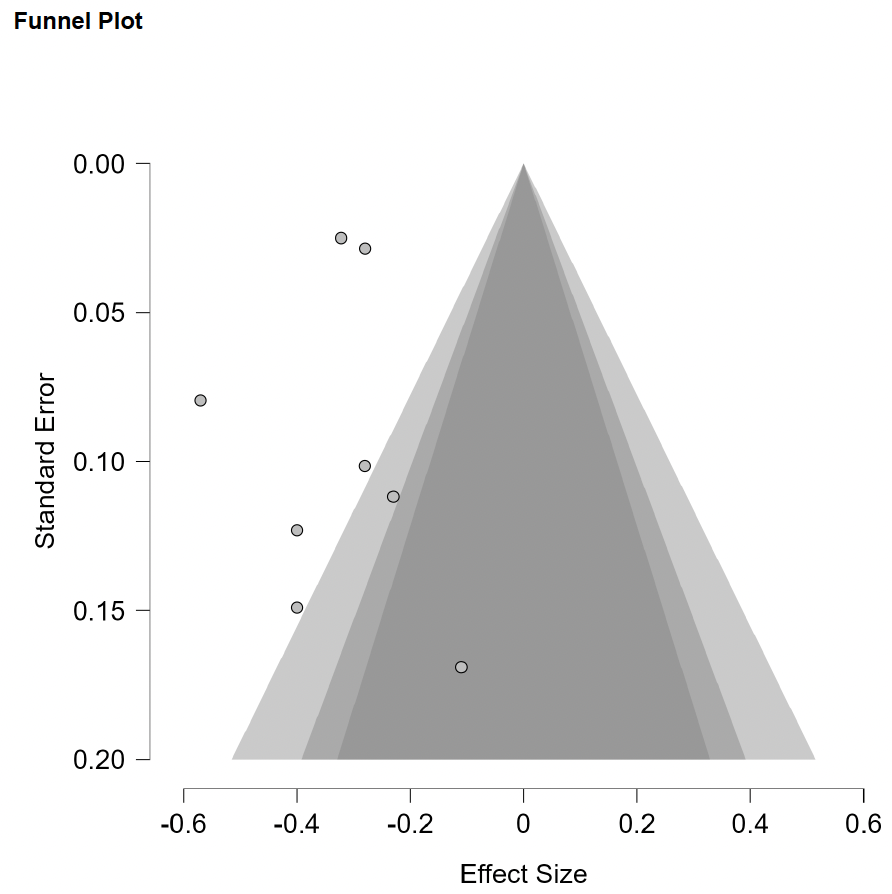

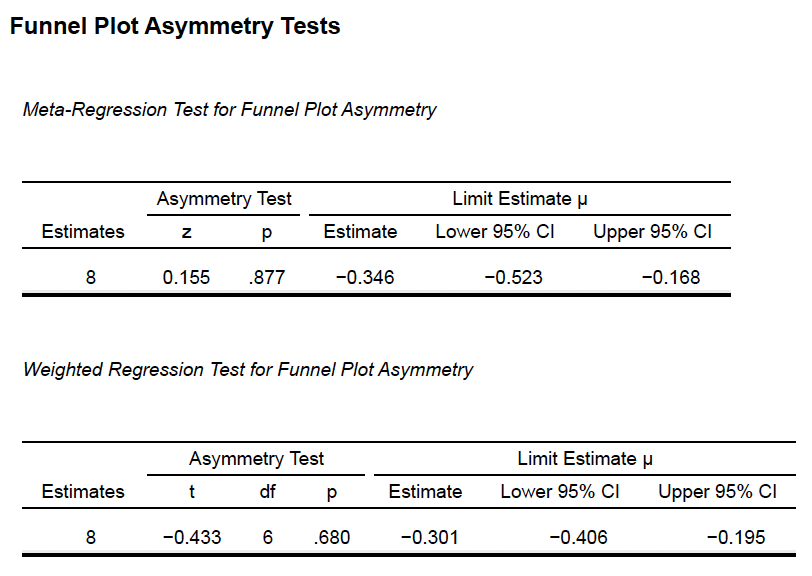

**Supplementary Methods 11b (Experience vs Conscientiousness vs Depersonalization)**

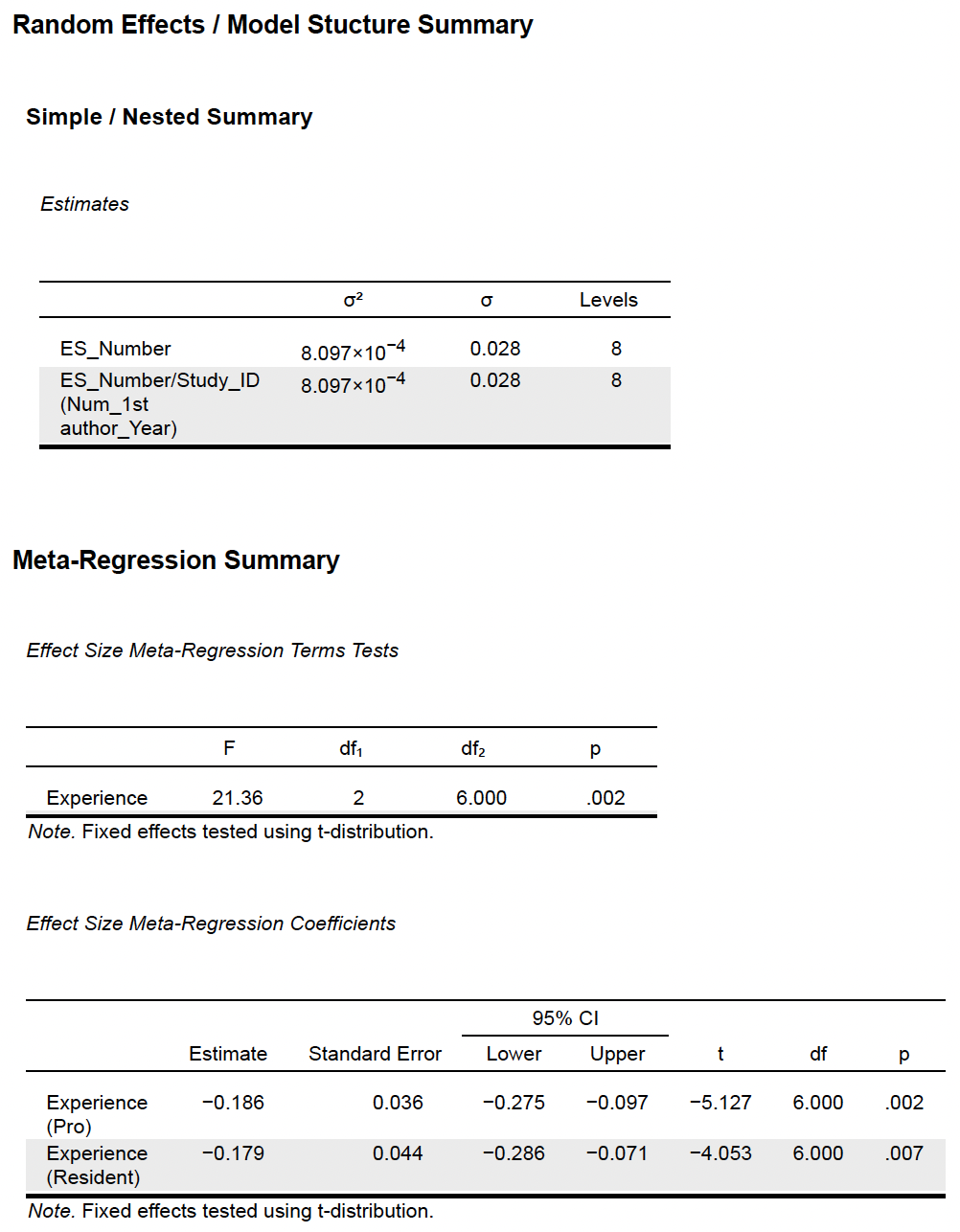

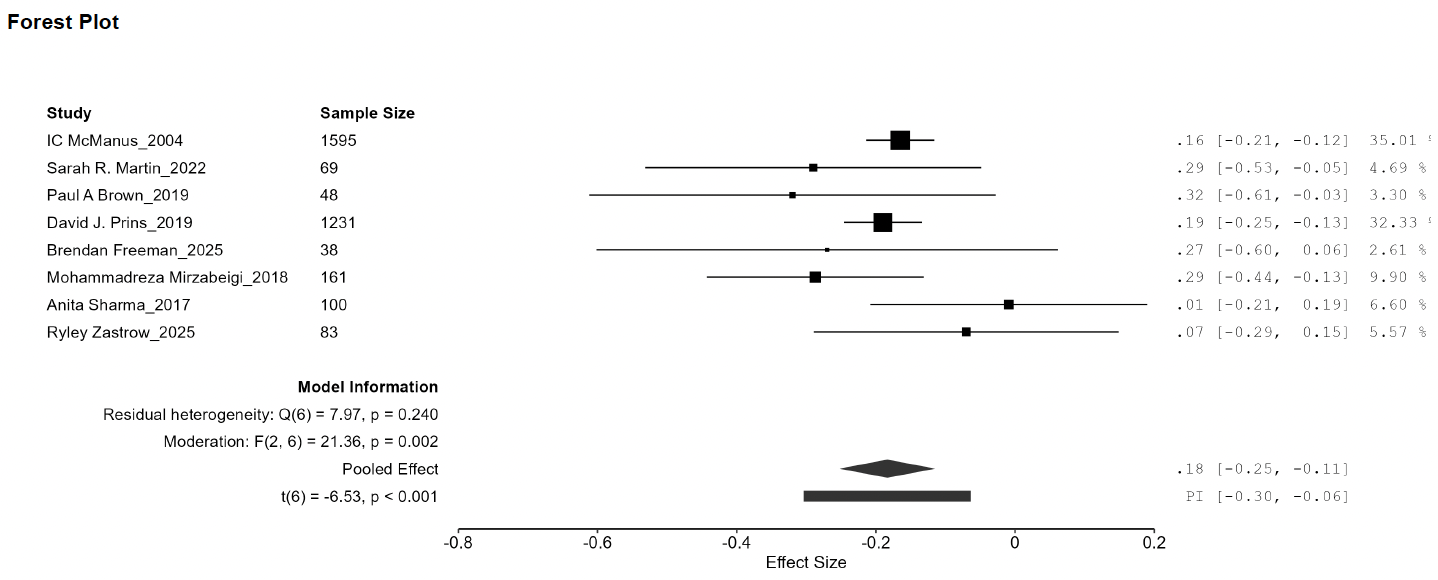

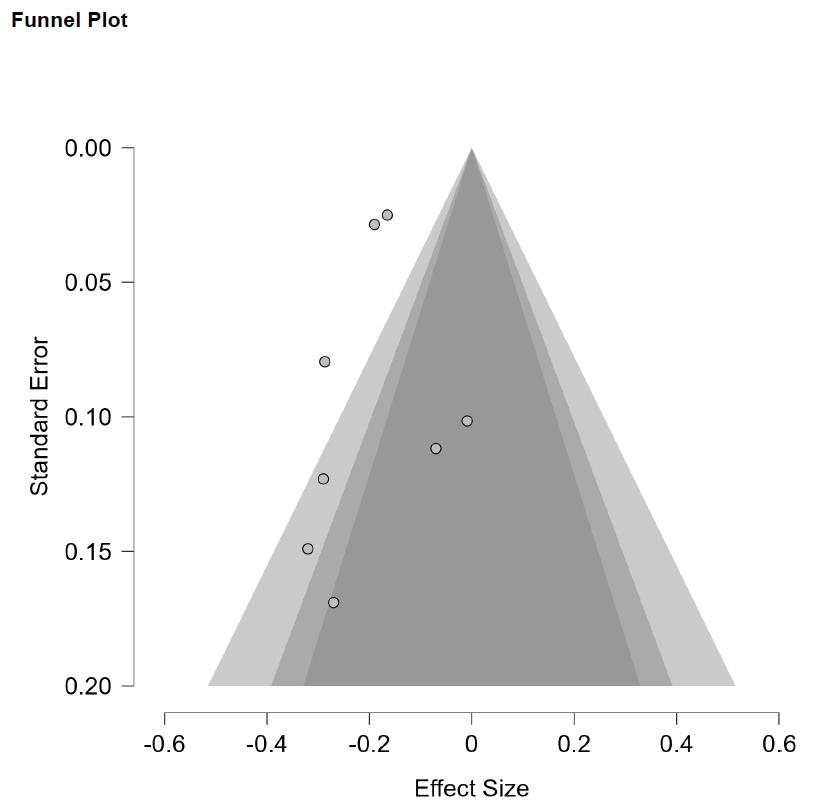

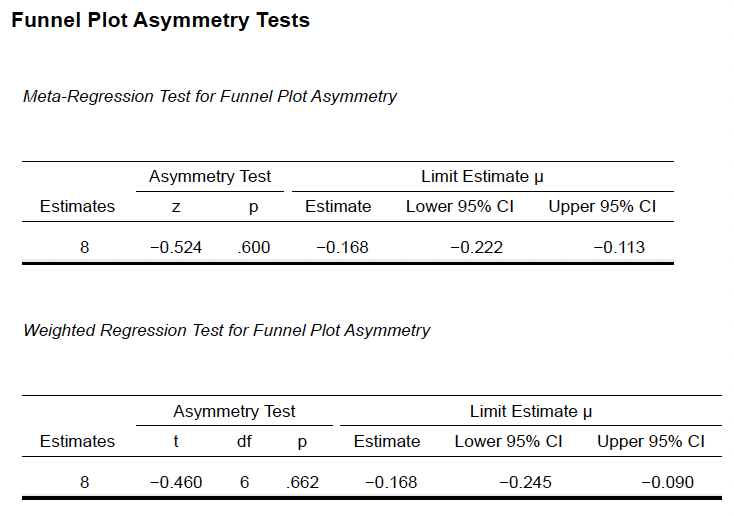

**Supplementary Methods 11c (Experience vs Extraversion vs Depersonalization)**

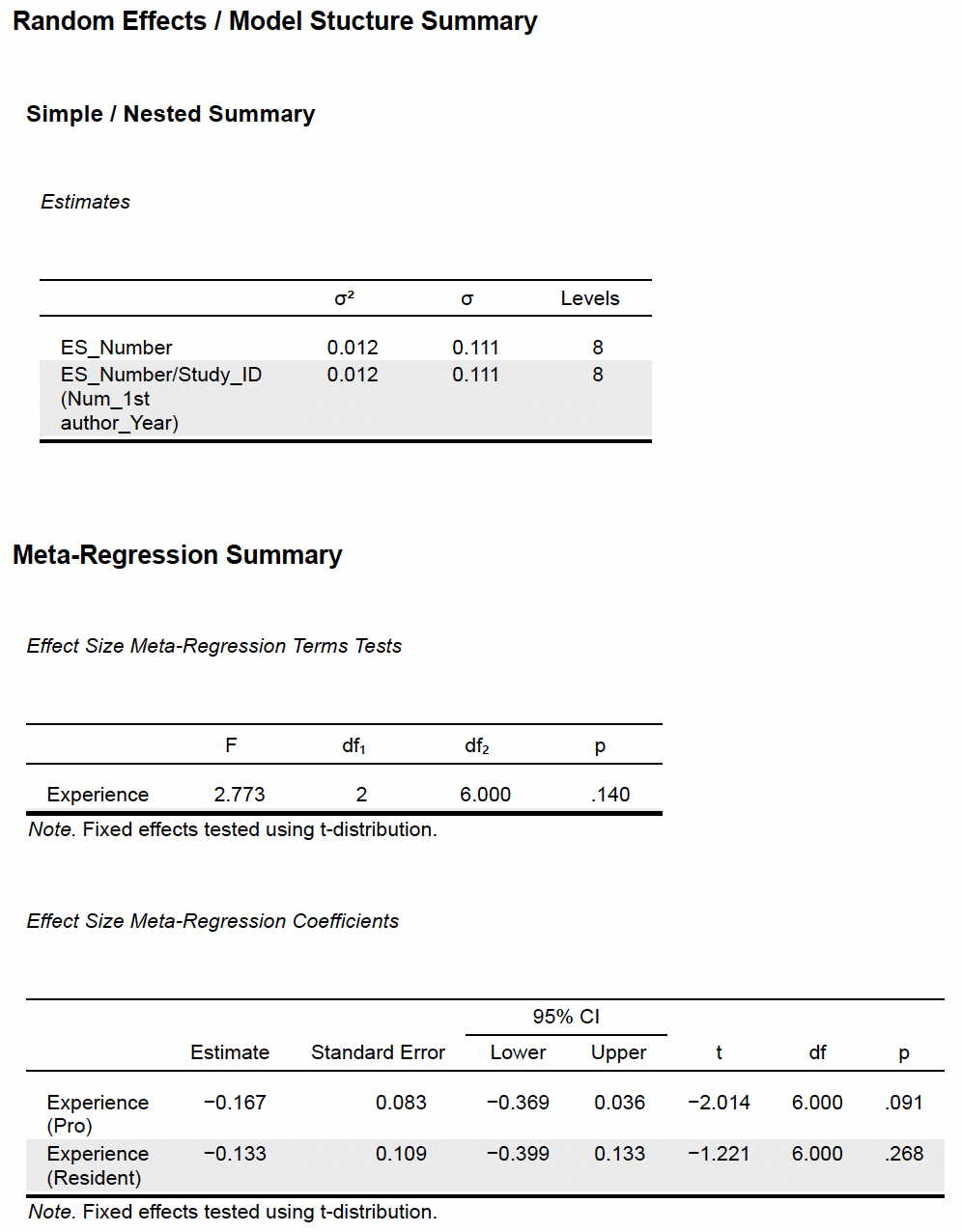

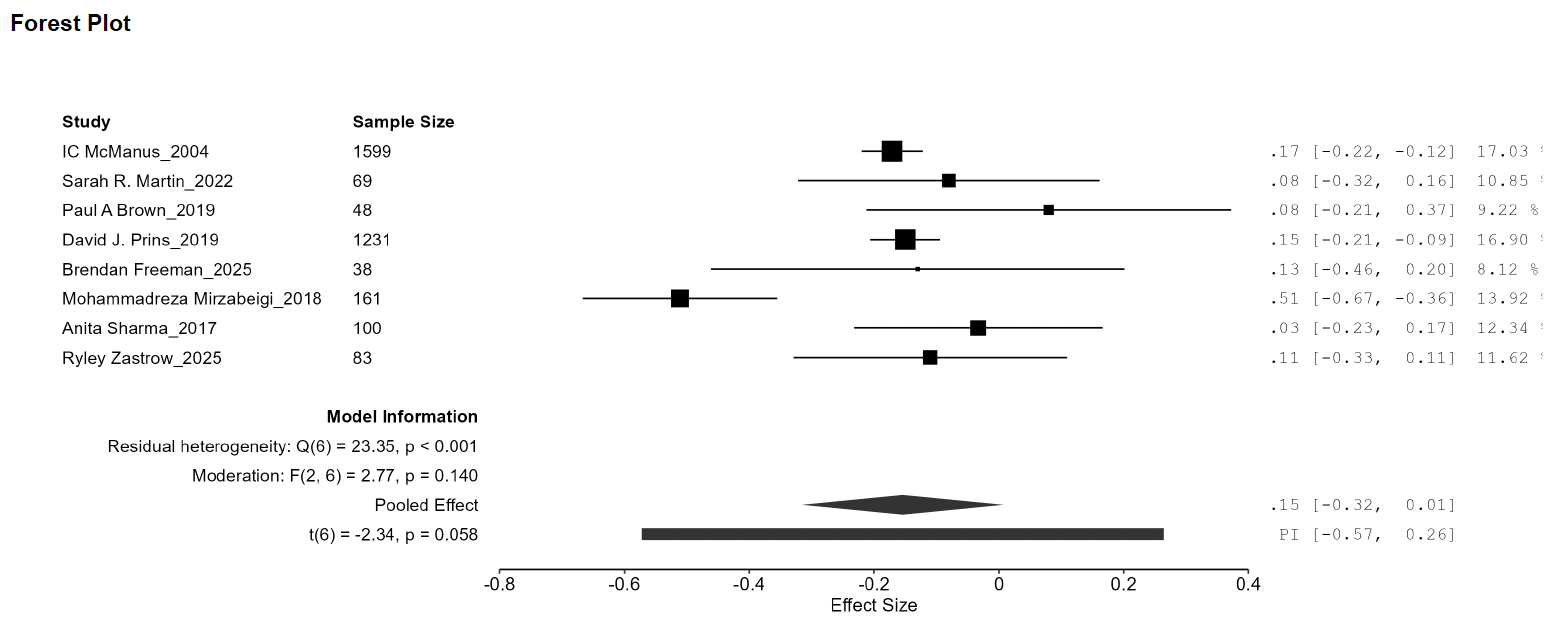

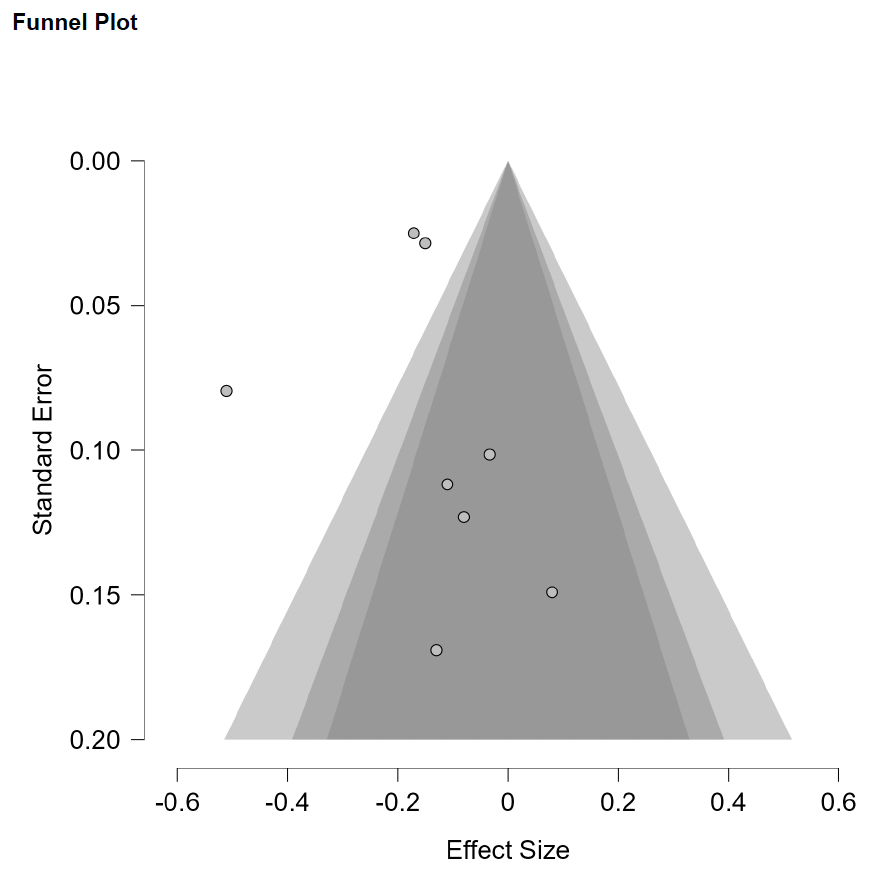

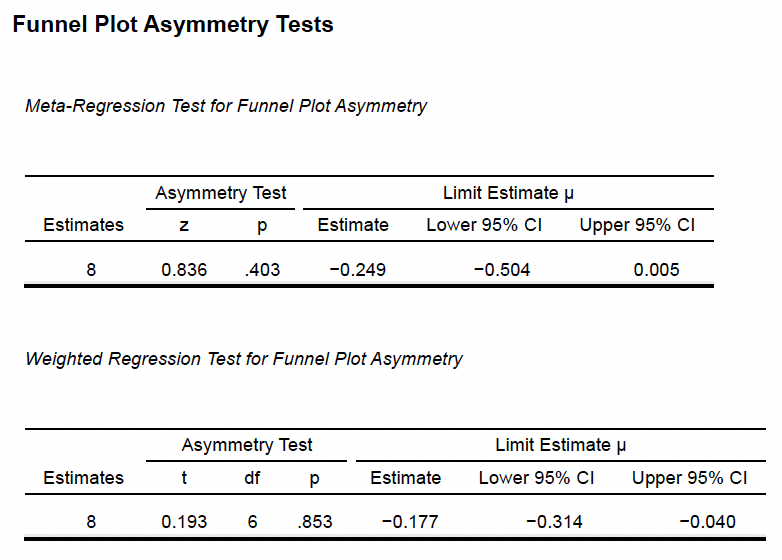

**Supplementary Methods 11d (Experience vs Neuroticism vs Depersonalization)**

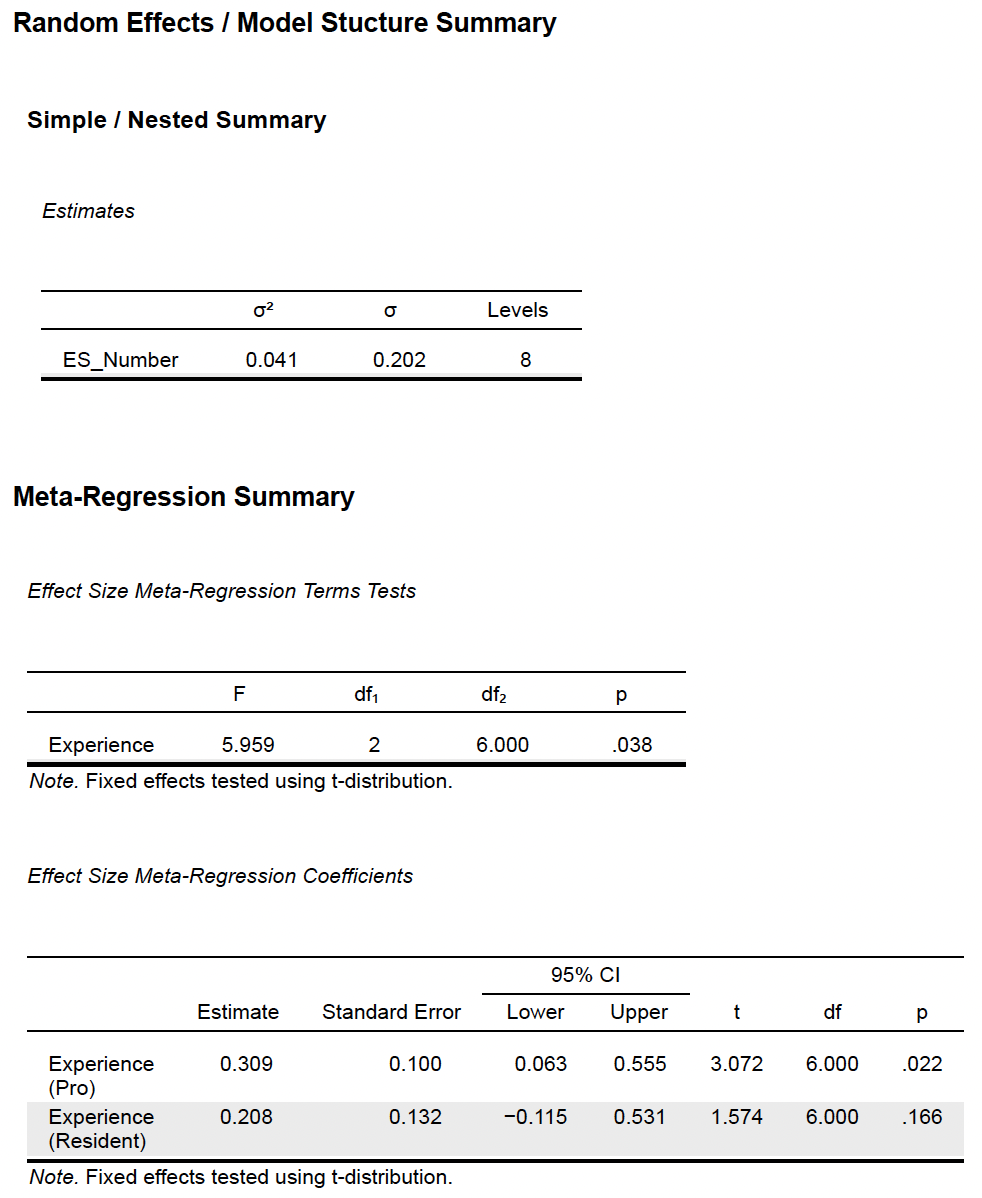

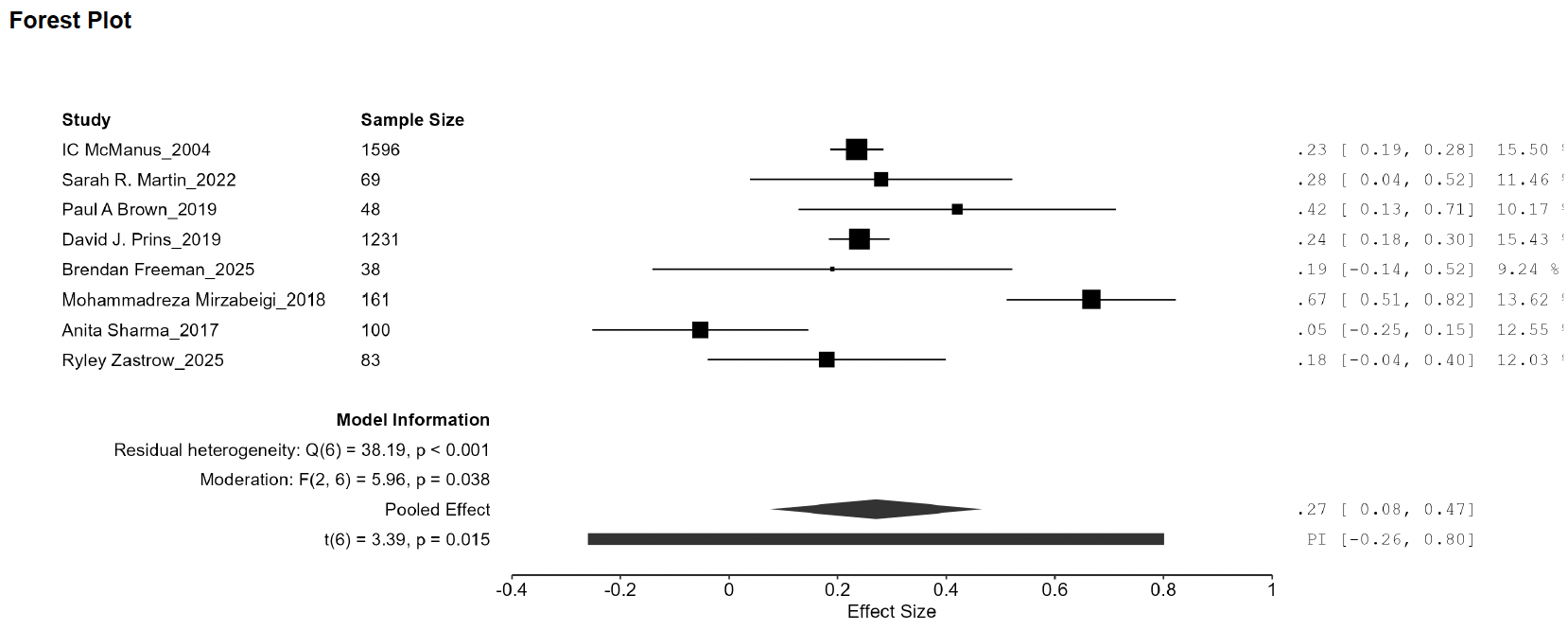

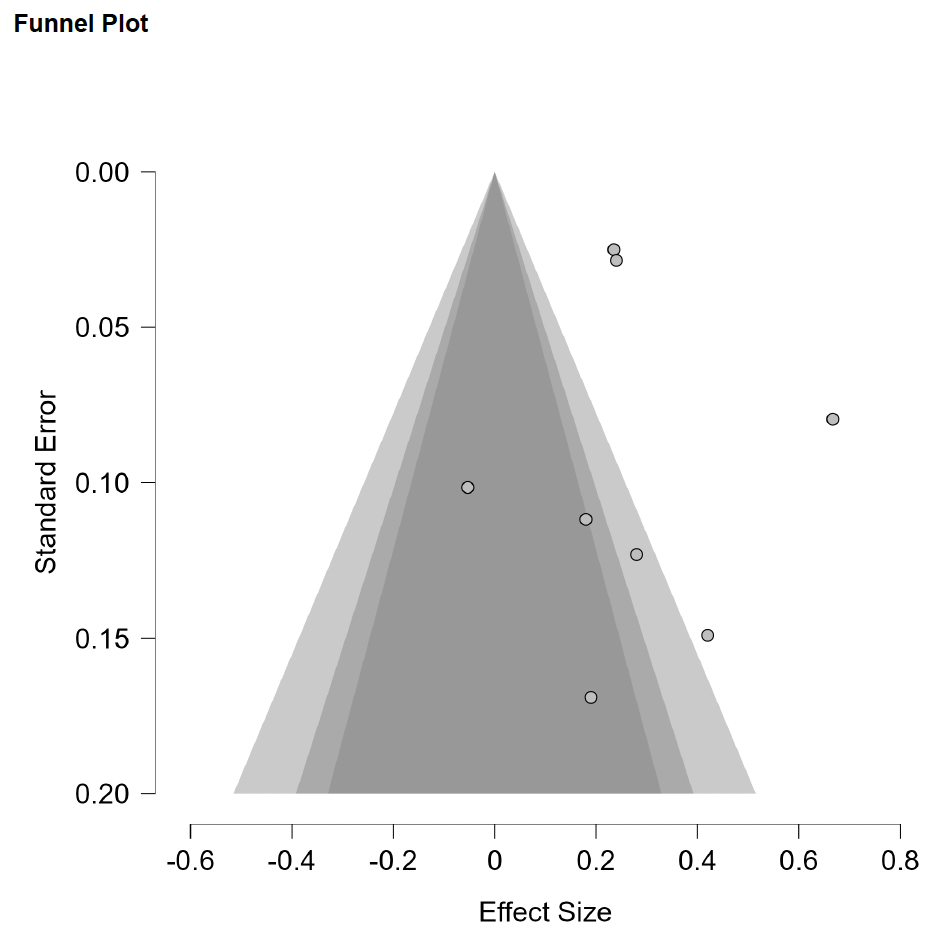

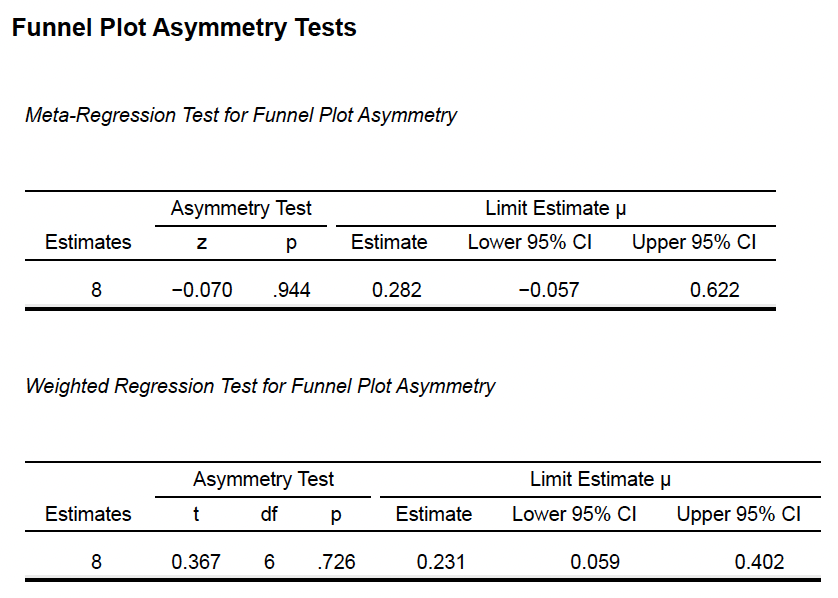

**Supplementary Methods 11e (Experience vs Openness vs Depersonalization)**

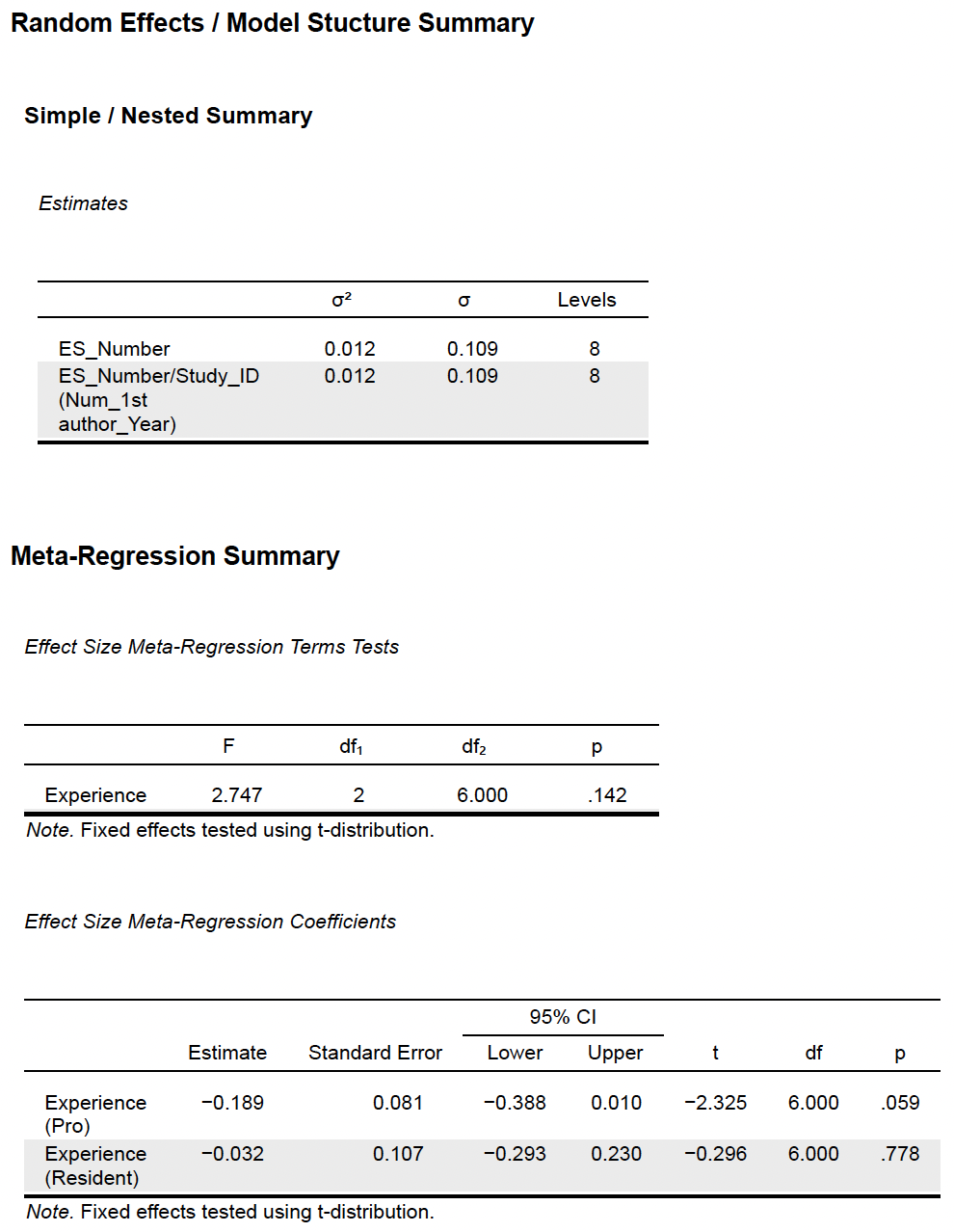

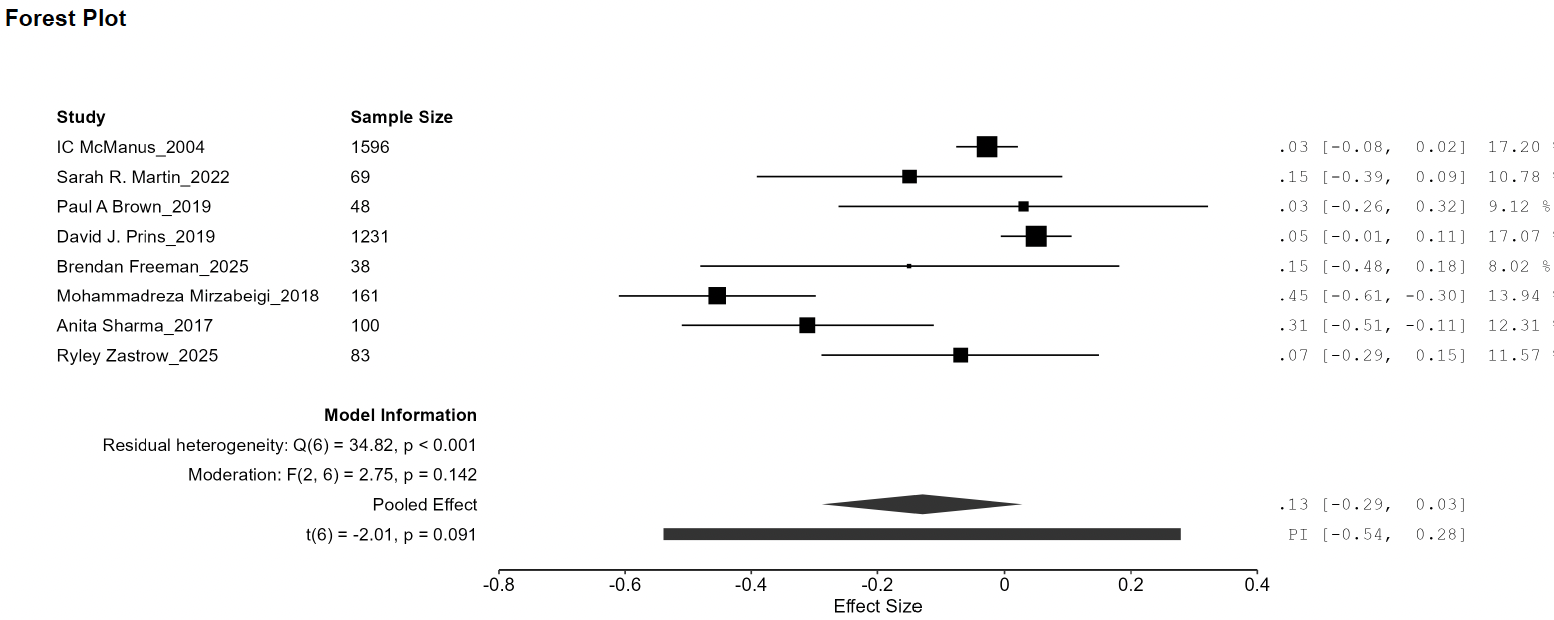

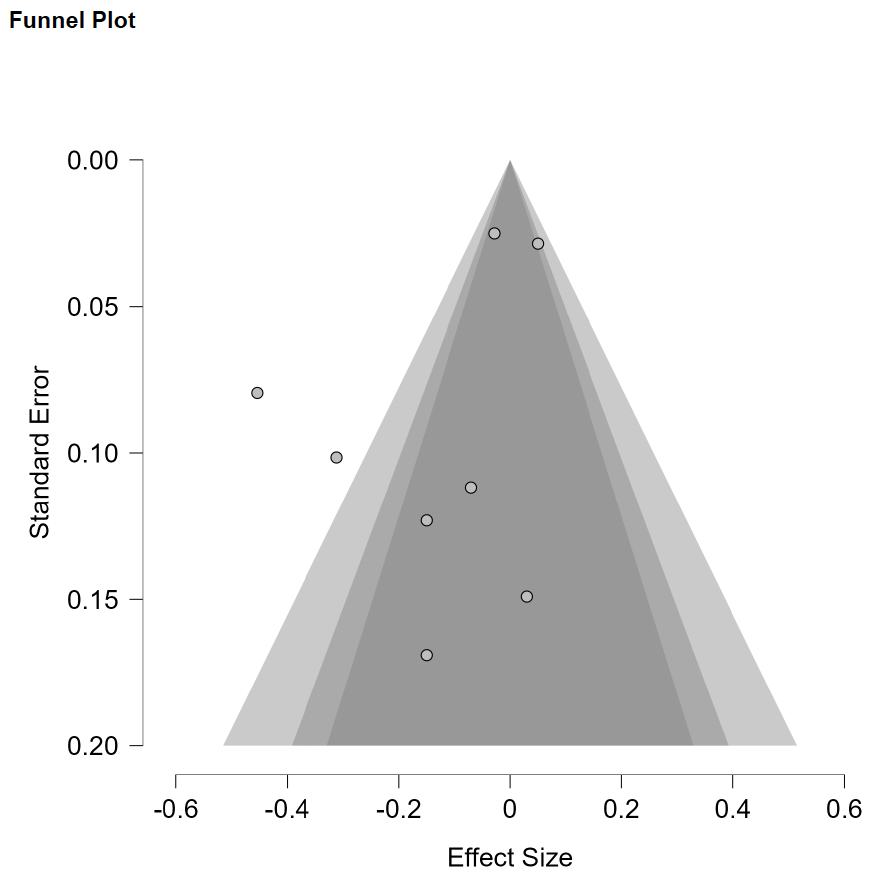

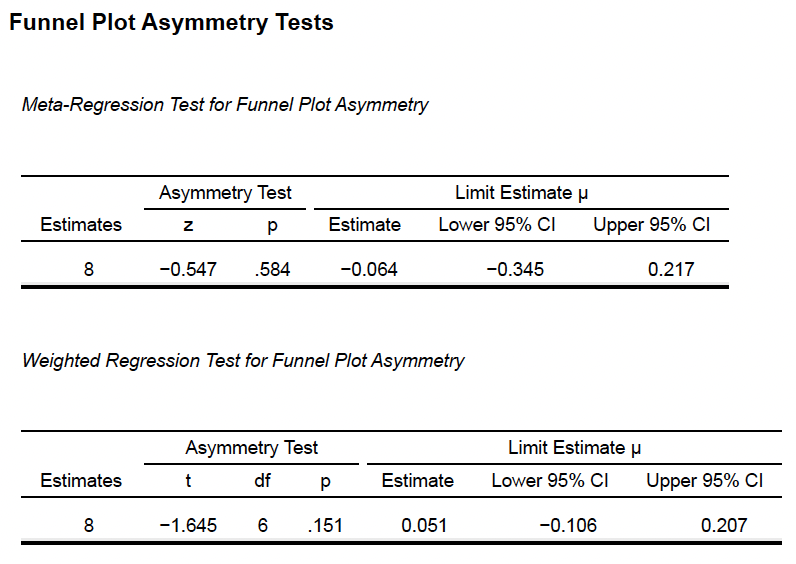

**Supplementary Methods 12a (Experience vs Agreeableness vs Emotional Exhaustion)**

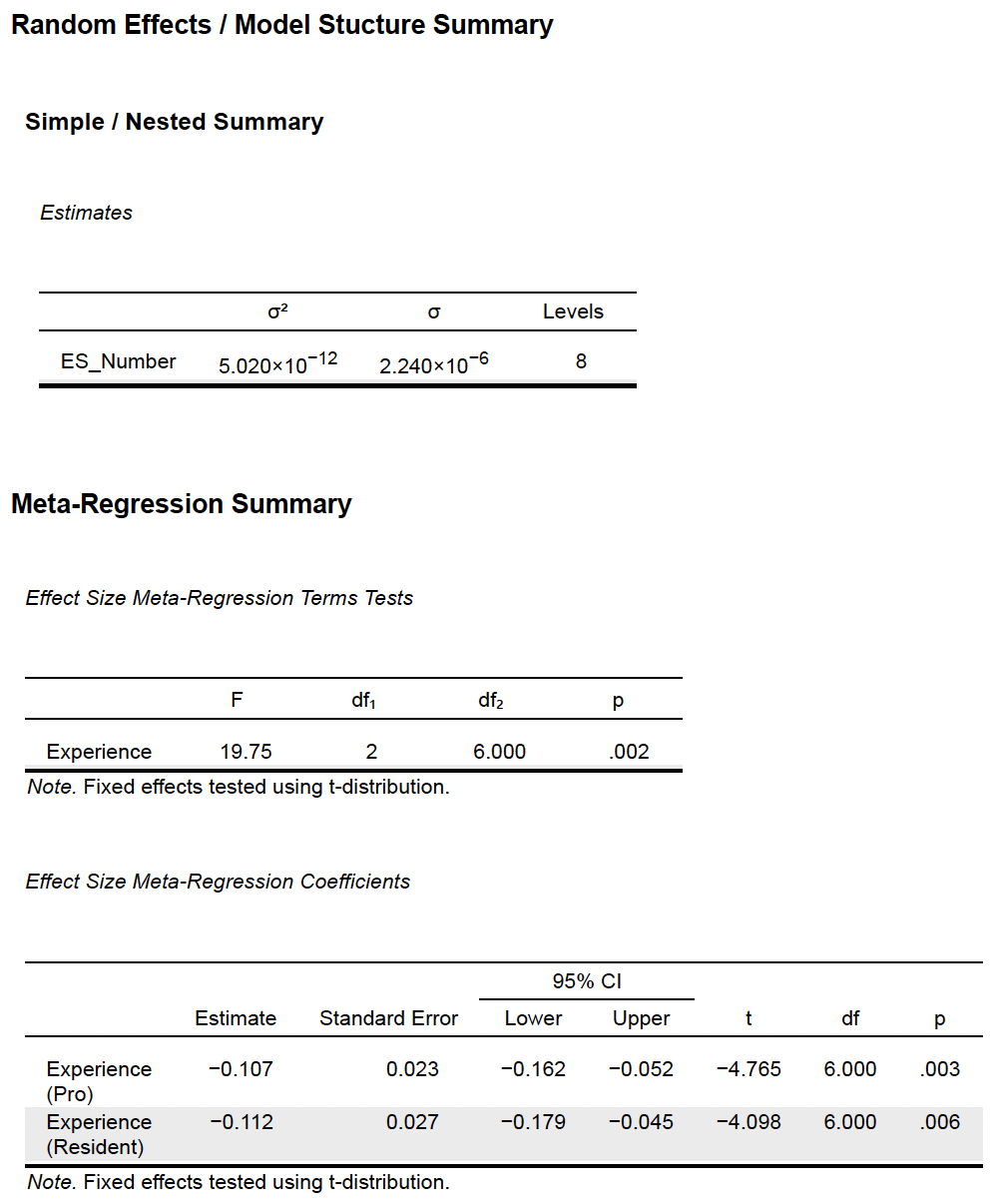

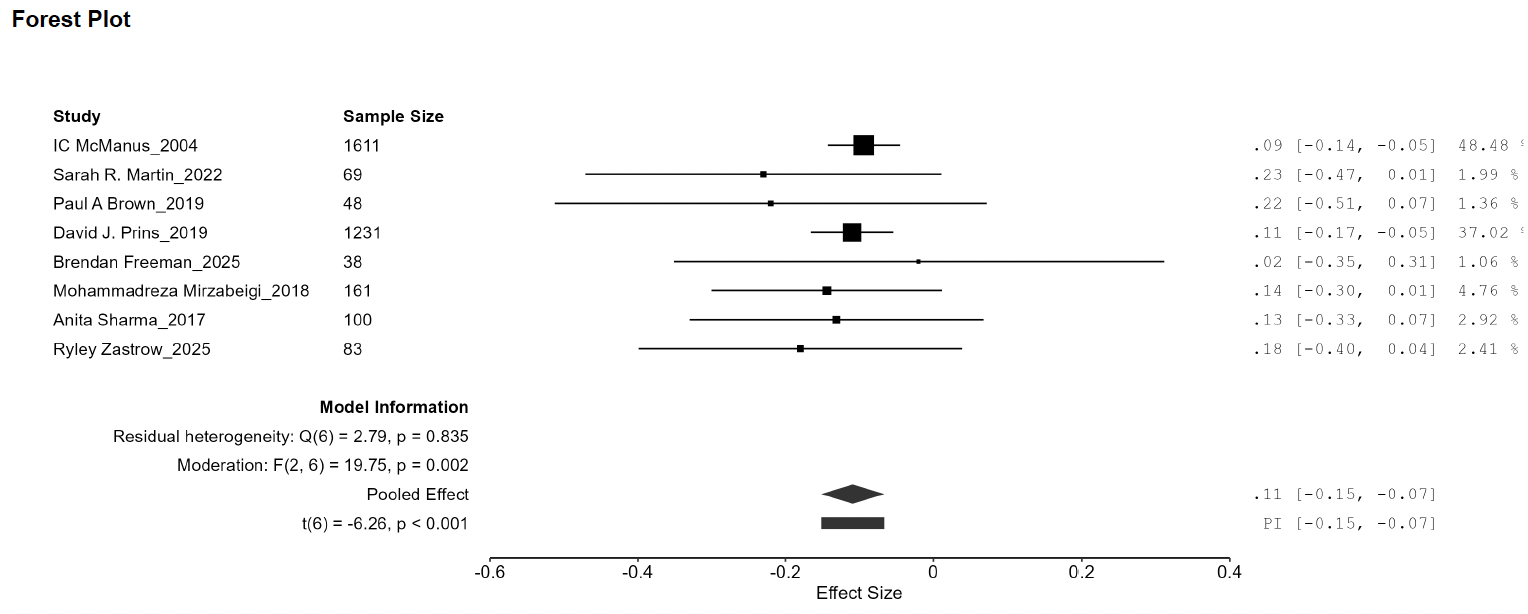

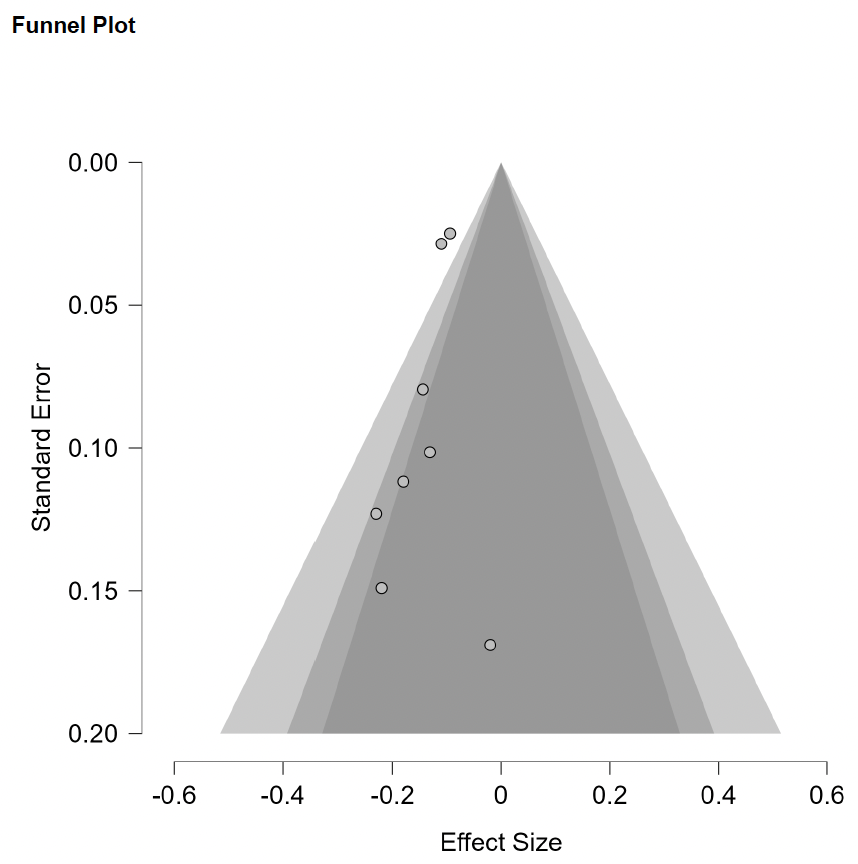

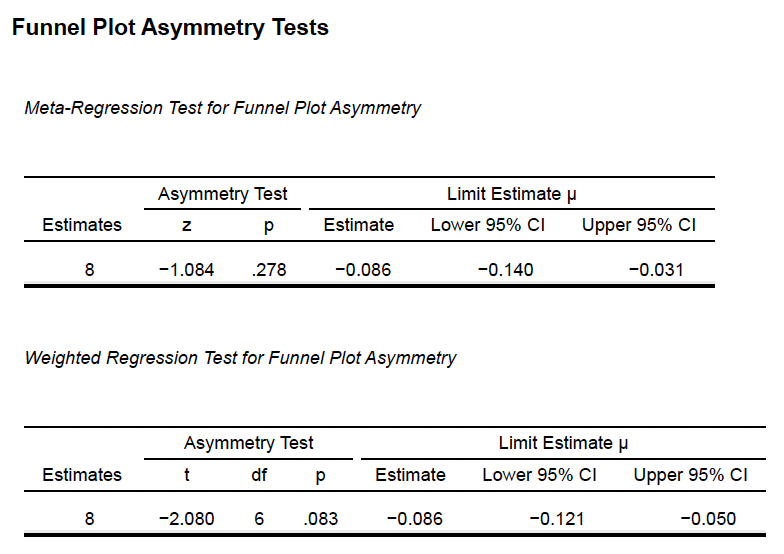

**Supplementary Methods 12b (Experience vs Conscietiousness vs Emotional Exhaustion)**

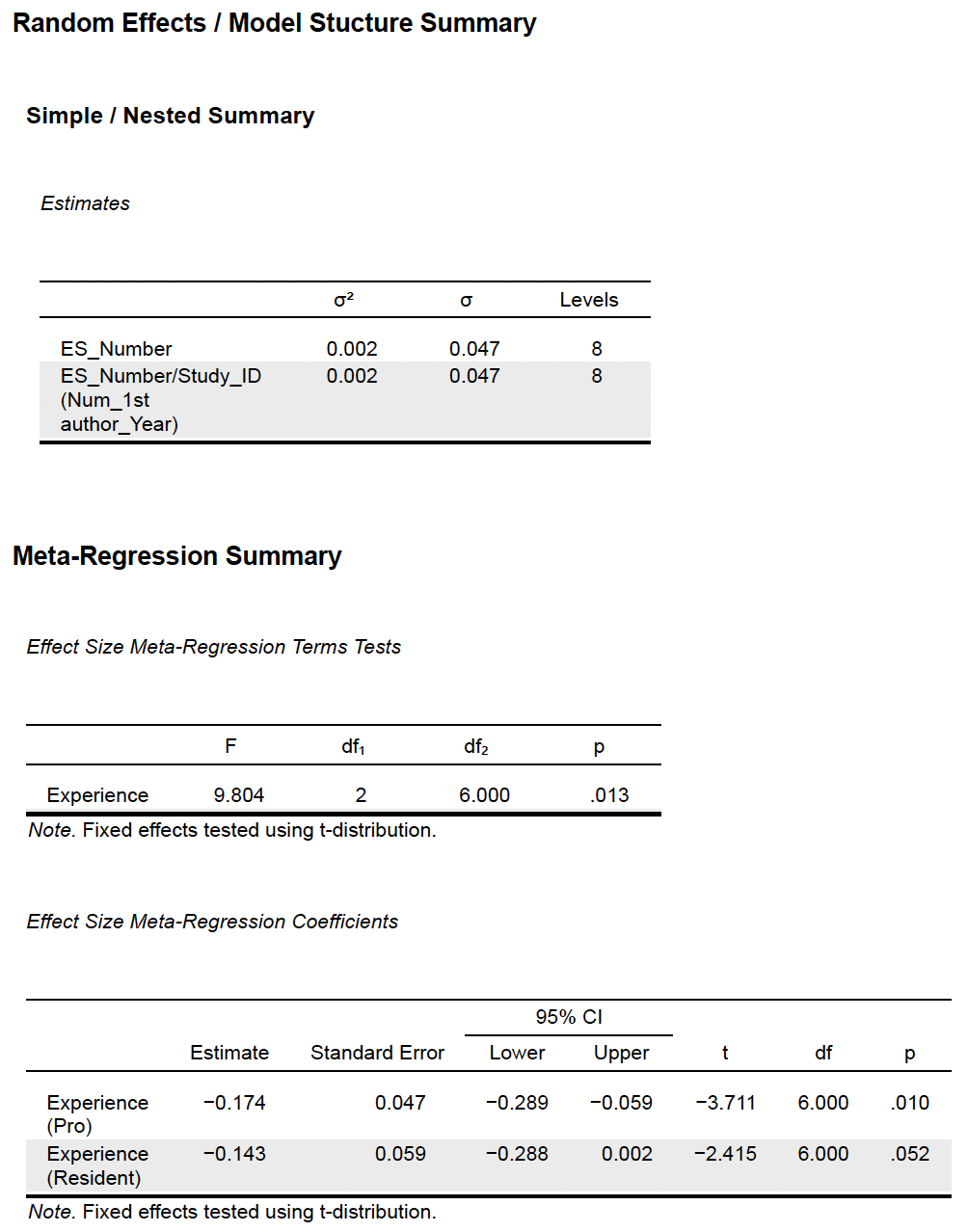

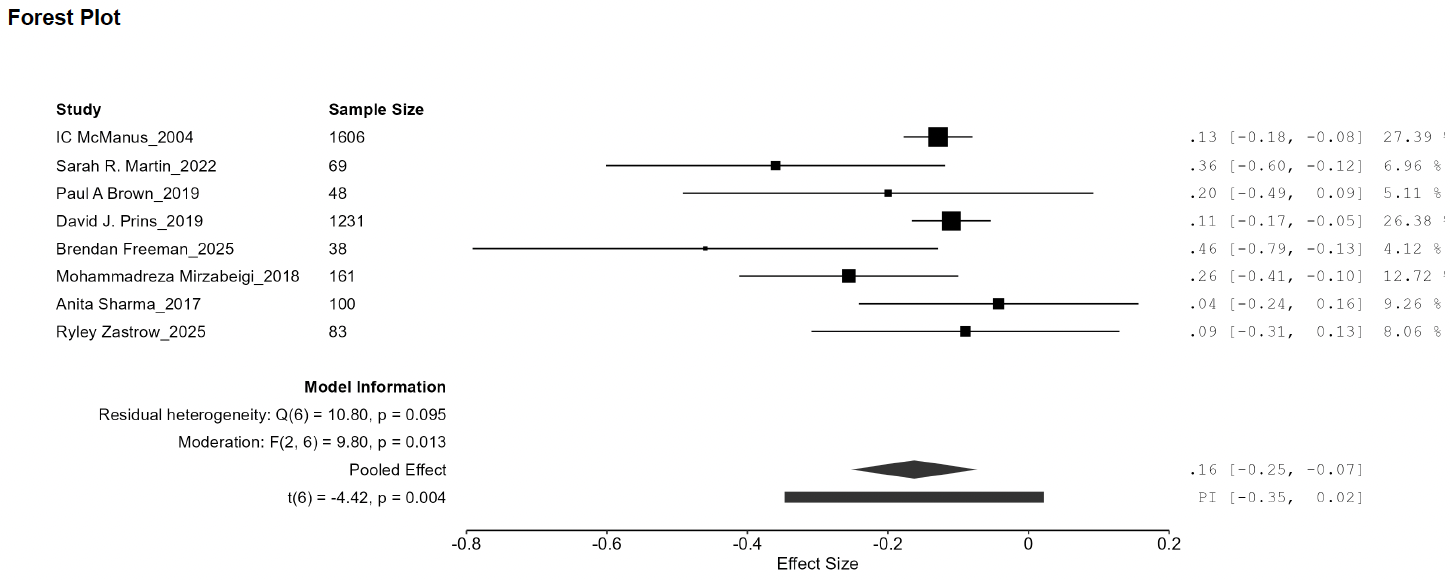

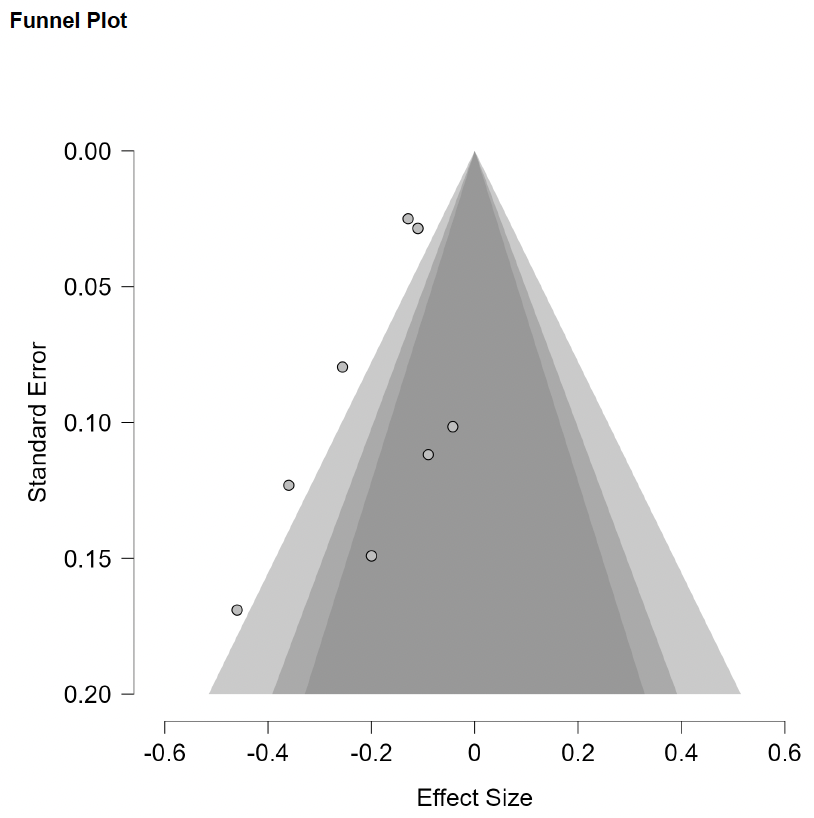

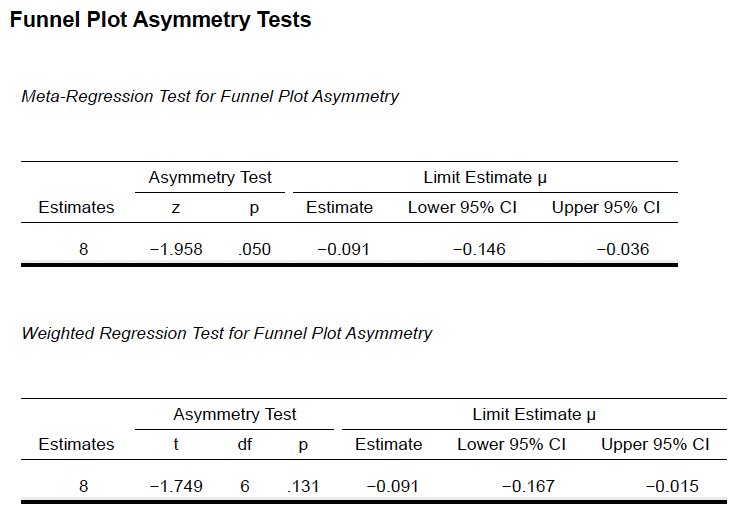

**Supplementary Methods 12c (Experience vs Extraversion vs Emotional Exhaustion)**

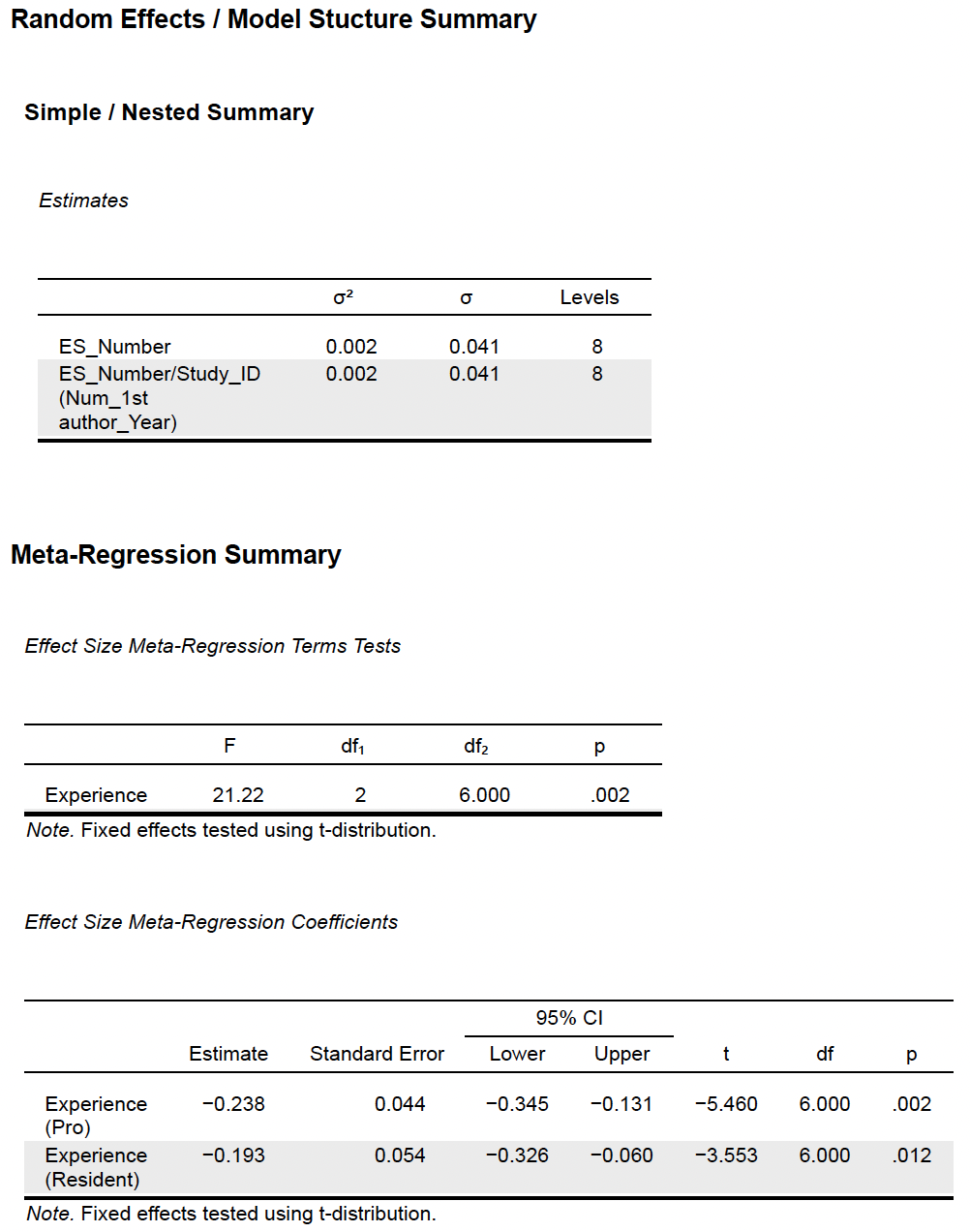

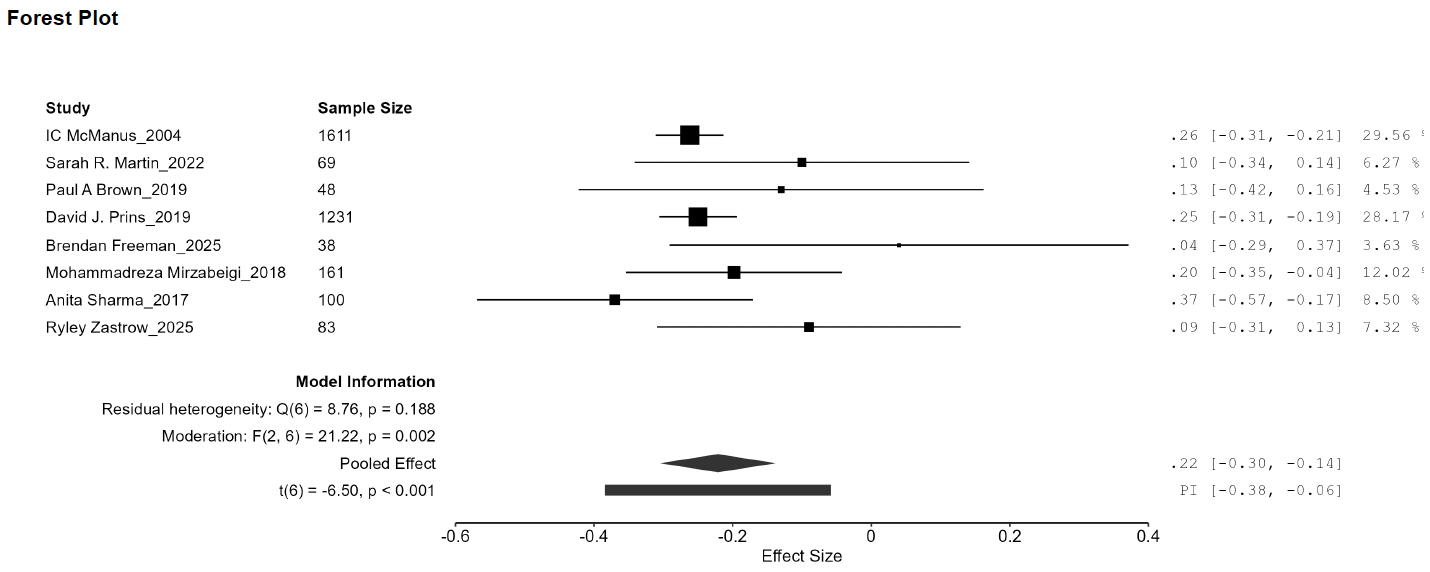

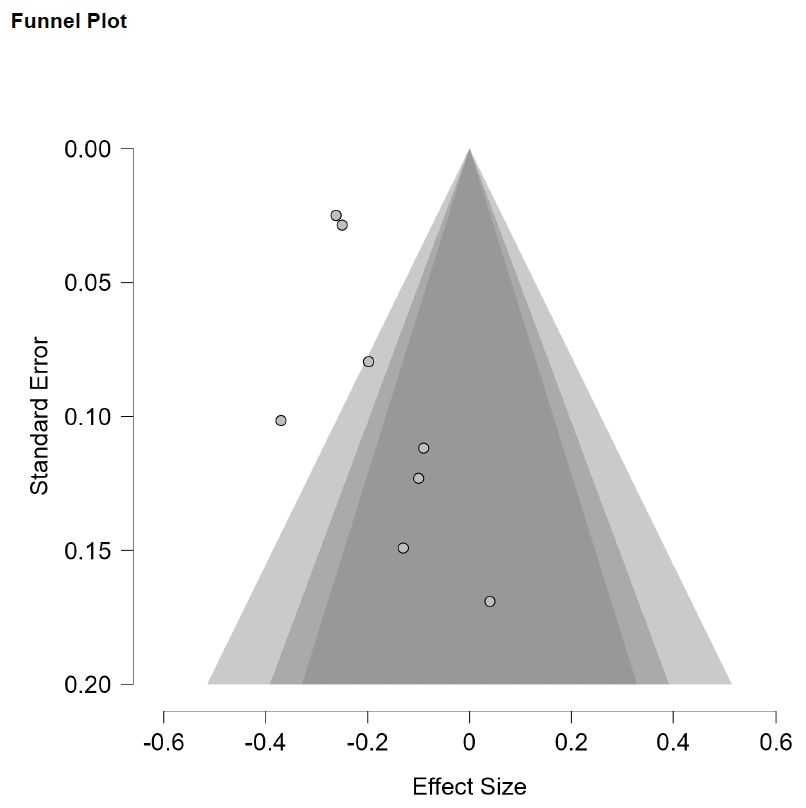

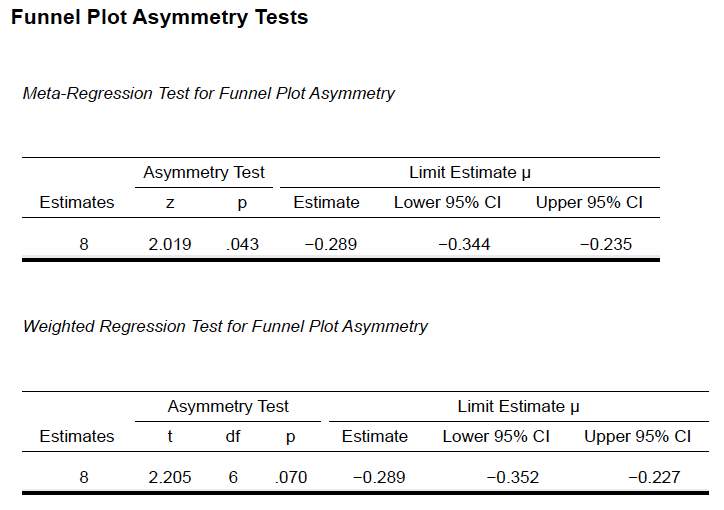

**Supplementary Methods 12d (Experience vs Neuroticism vs Emotional Exhaustion)**

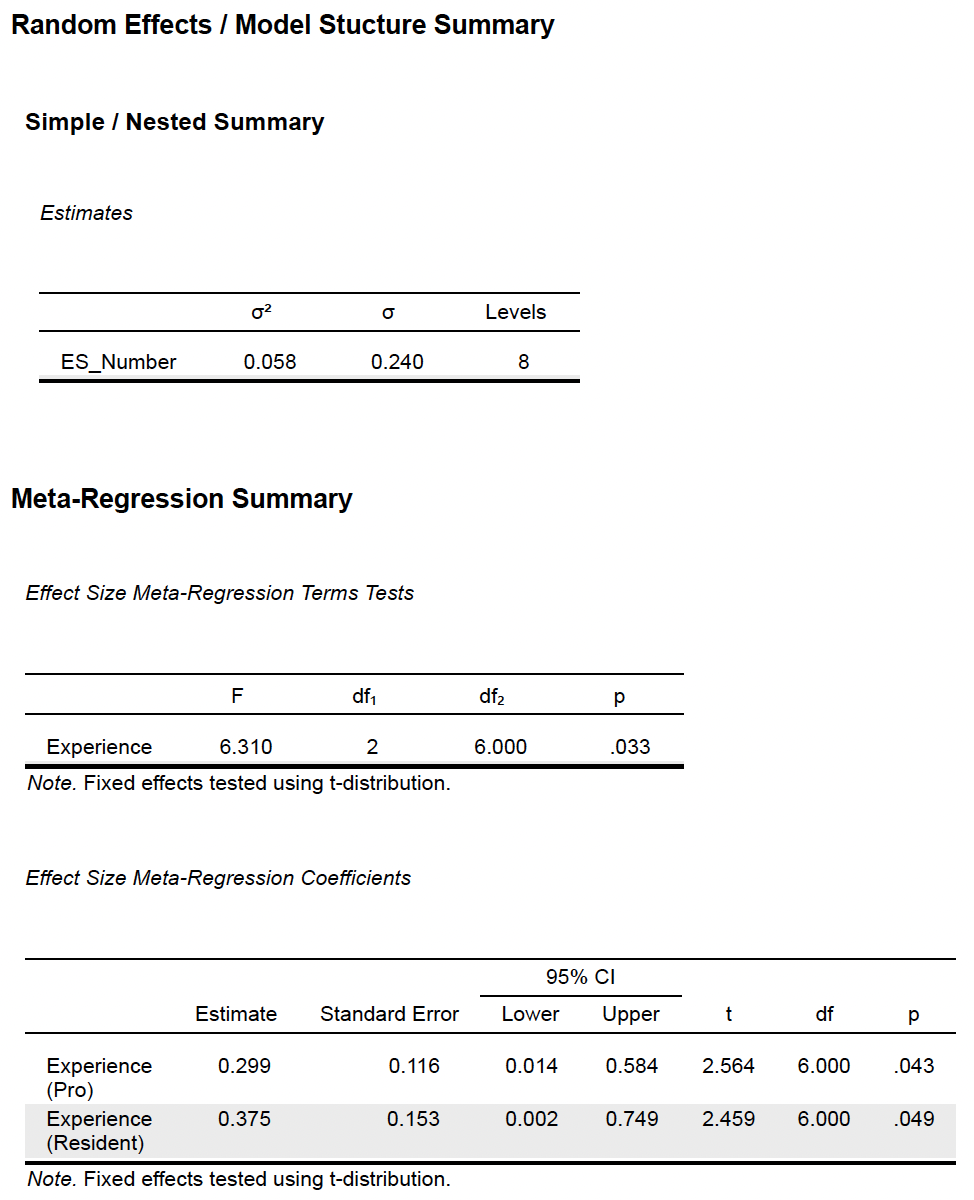

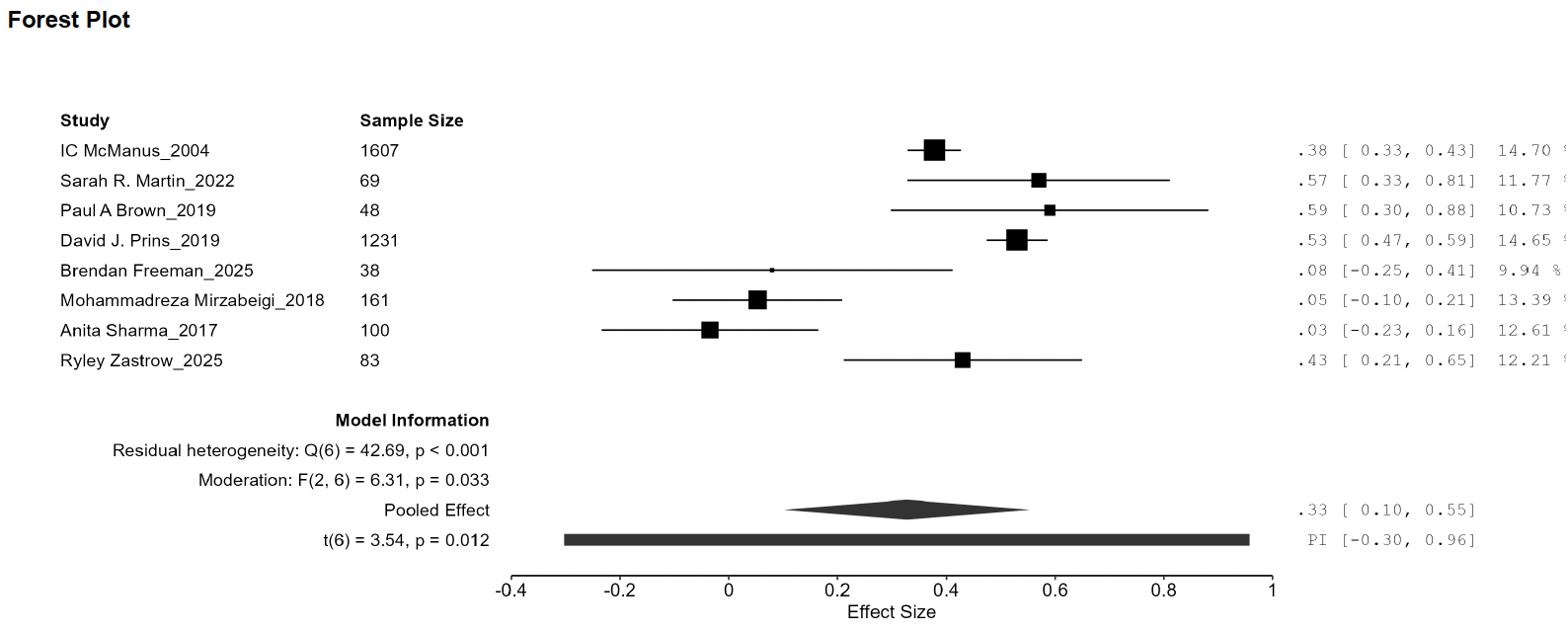

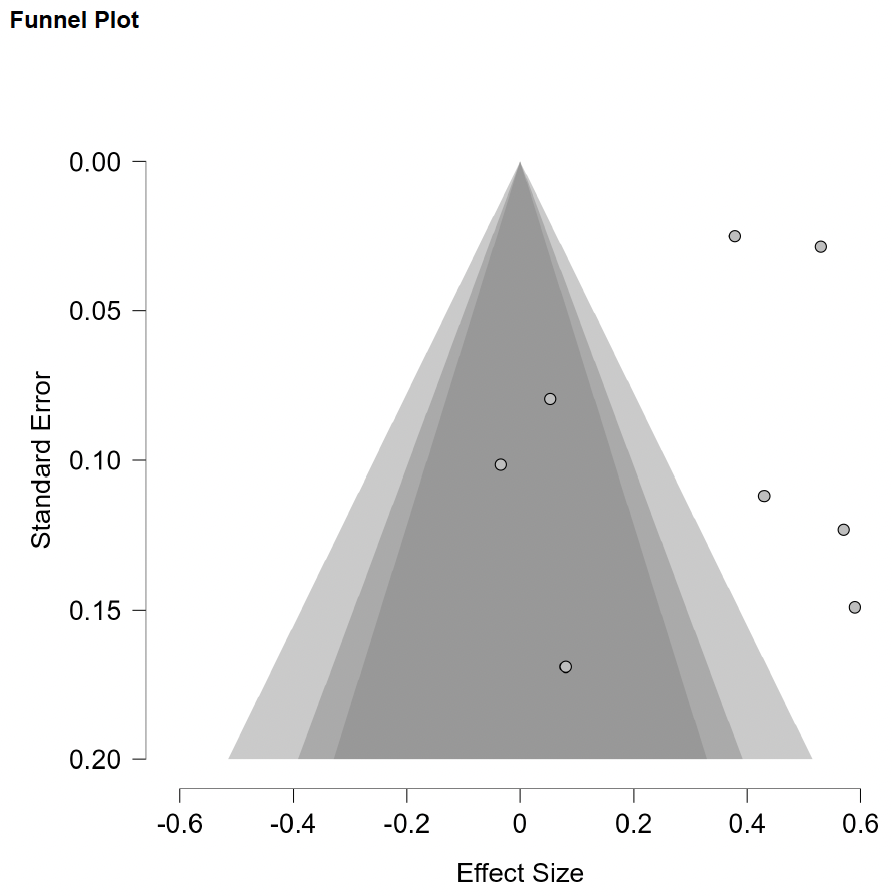

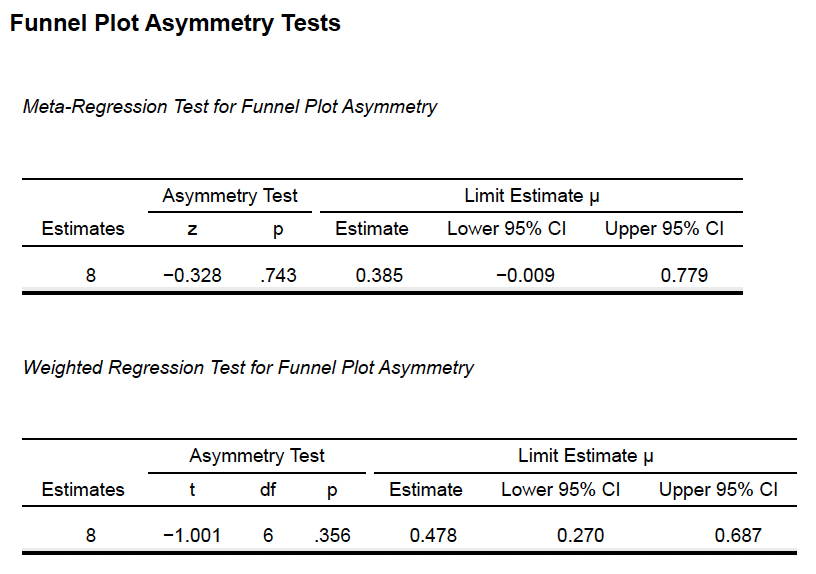

**Supplementary Methods 12e (Experience vs Openness vs Emotional Exhaustion)**

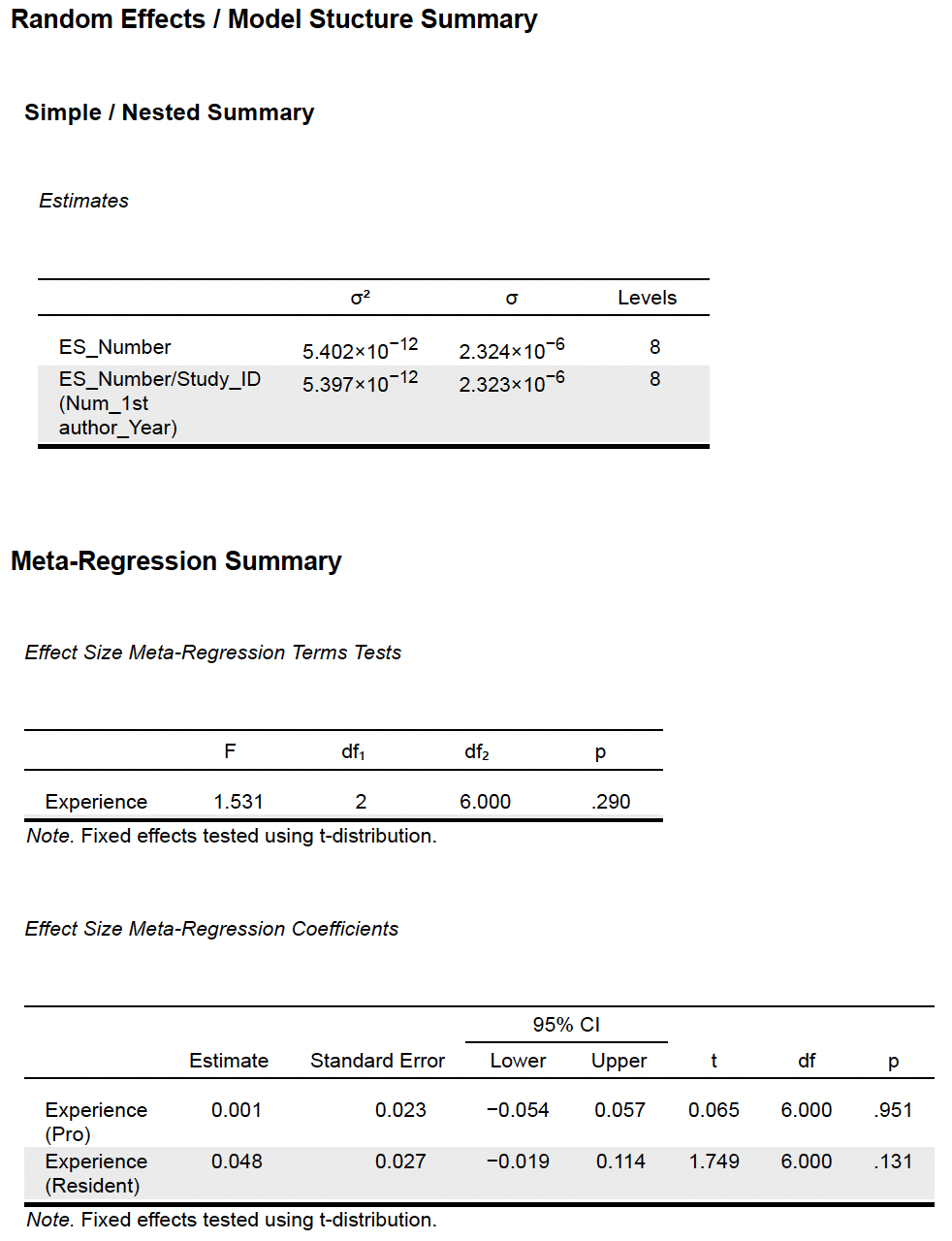

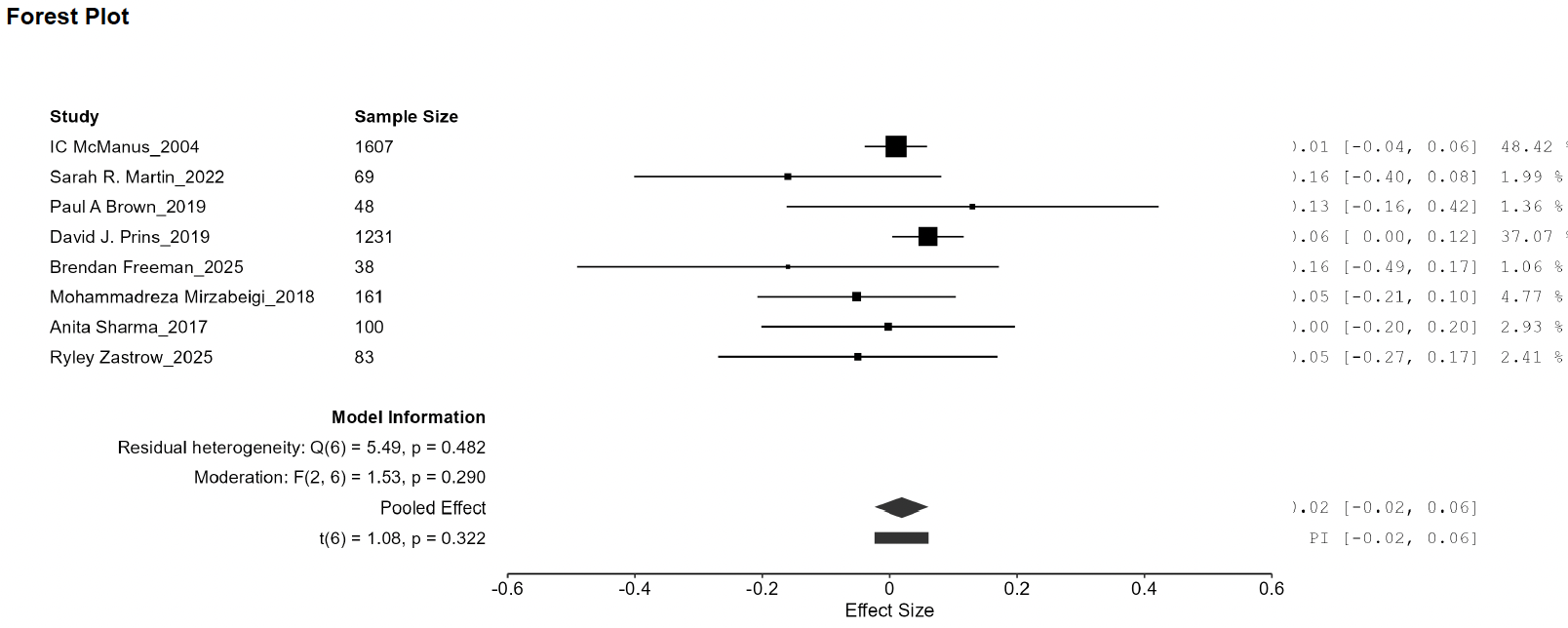

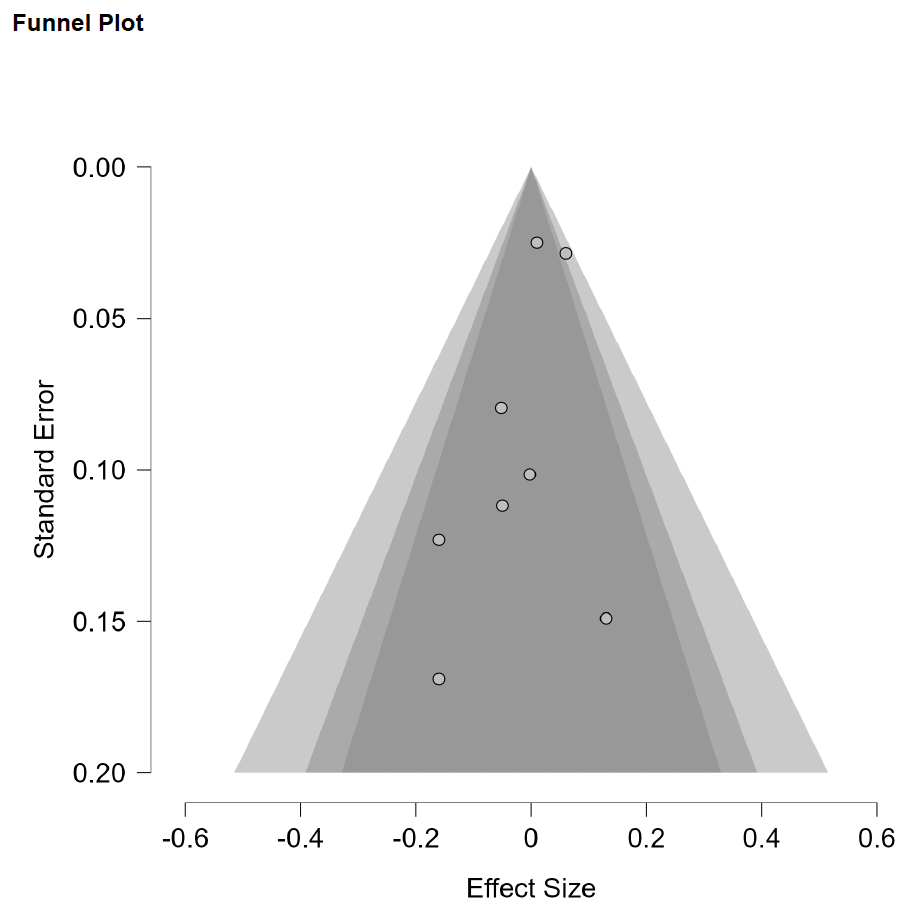

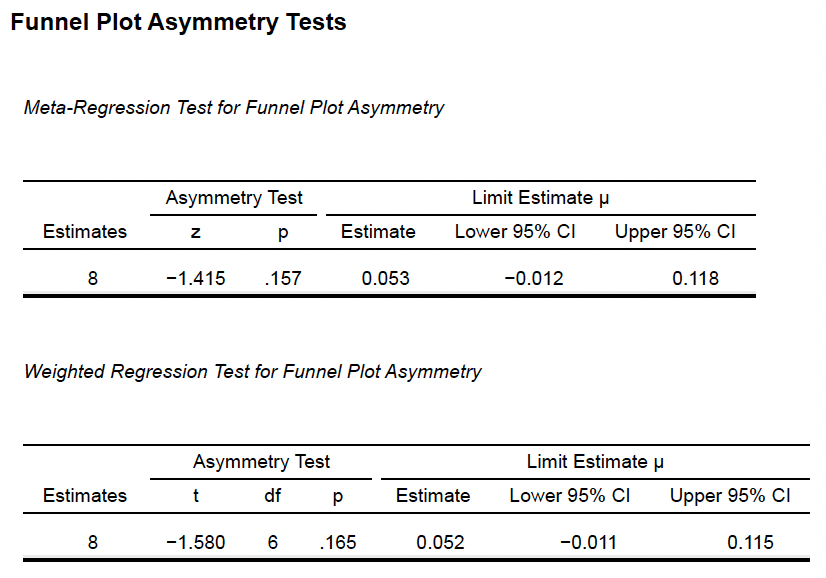

**Supplementary Methods 13a (Experience vs Agreeableness vs Personal Accomplishment)**

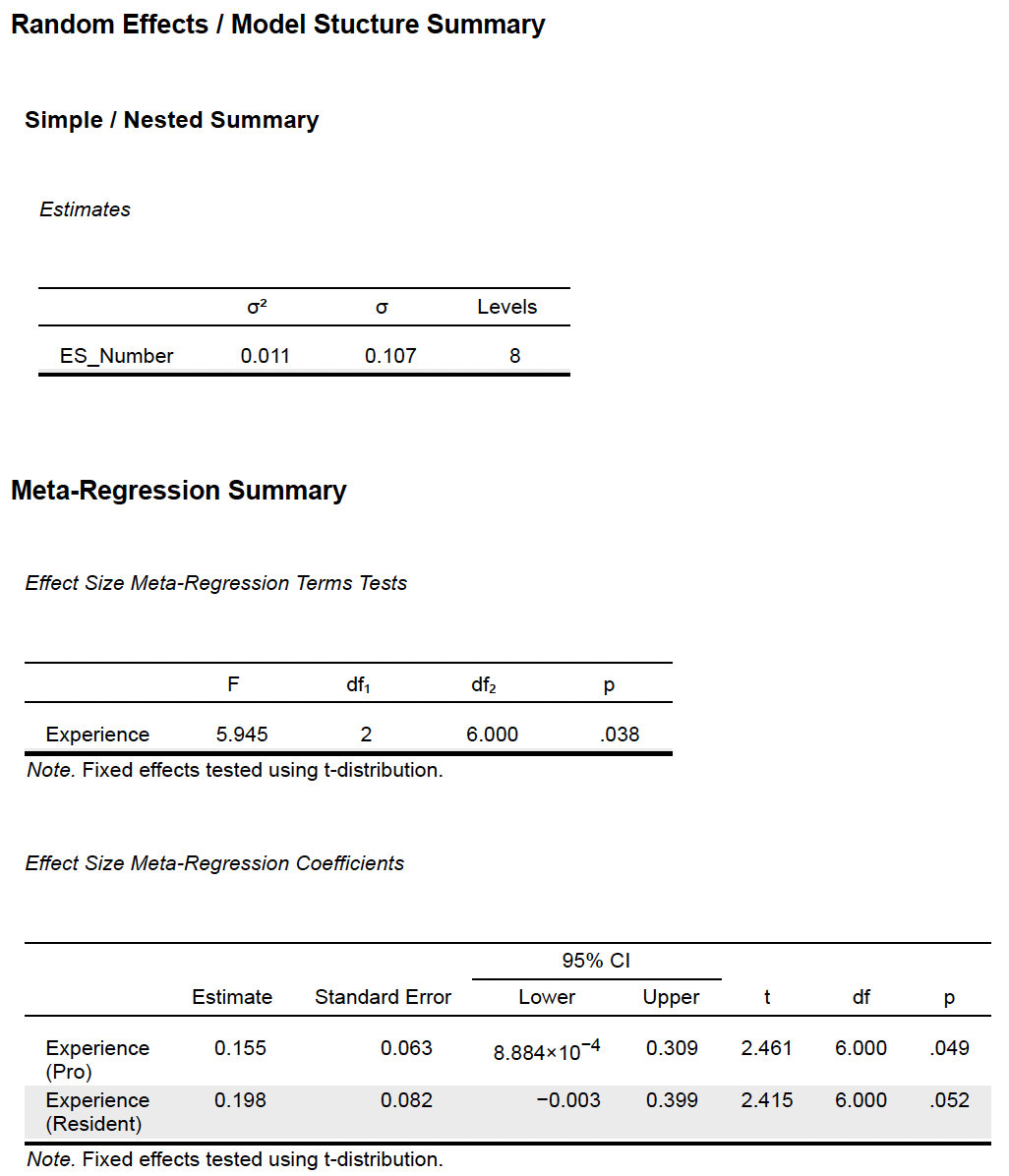

**Supplementary Methods 13b (Experience vs Conscientiousness vs Personal Accomplishment)**

**Supplementary Methods 13c (Experience vs Extraversion vs Personal Accomplishment)**

**Supplementary Methods 13d (Experience vs Neuroticism vs Personal Accomplishment)**

**Supplementary Methods 13e (Experience vs Openness vs Personal Accomplishment)**

**Supplementary Methods 14a (Region vs Agreeableness vs Depersonalization)**

**Supplementary Methods 14b (Region vs Conscientiousness vs Depersonalization)**

**Supplementary Methods 14c (Region vs Extraversion vs Depersonalization)**

**Supplementary Methods 14d (Region vs Neuroticism vs Depersonalization)**

**Supplementary Methods 14e (Region vs Openness vs Depersonalization)**

**Supplementary Methods 15a (Region vs Agreeableness vs Emotional Exhaustion)**

**Supplementary Methods 15b (Region vs Conscientiousness vs Emotional Exhaustion)**

**Supplementary Methods 15c (Region vs Extraversion vs Emotional Exhaustion)**

**Supplementary Methods 15d (Region vs Neuroticism vs Emotional Exhaustion)**

**Supplementary Methods 15e (Region vs Openness vs Emotional Exhaustion)**

**Supplementary Methods 16a (Region vs Agreeableness vs Personal Accomplishment)**

**Supplementary Methods 16b (Region vs Conscientiousness vs Personal Accomplishment)**

**Supplementary Methods 16c (Region vs Extraversion vs Personal Accomplishment)**

**Supplementary Methods 16d (Region vs Neuroticism vs Personal Accomplishment)**

**Supplementary Methods 16e (Region vs Openness vs Personal Accomplishment)**

**Supplementary Methods 17a (Specialty vs Agreeableness vs Depersonalization)**

**Supplementary Methods 17b (Specialty vs Conscientiousness vs Depersonalization)**

**Supplementary Methods 17c (Specialty vs Extraversion vs Depersonalization)**

**Supplementary Methods 17d (Specialty vs Neuroticism vs Depersonalization)**

**Supplementary Methods 17e (Specialty vs Openness vs Depersonalization)**

**Supplementary Methods 18a (Specialty vs Agreeableness vs Emotional Exhaustion)**

**Supplementary Methods 18b (Specialty vs Conscientiousness vs Emotional Exhaustion)**

**Supplementary Methods 18c (Specialty vs Extraversion vs Emotional Exhaustion)**

**Supplementary Methods 18d (Specialty vs Neuroticism vs Emotional Exhaustion)**

**Supplementary Methods 18e (Specialty vs Openness vs Emotional Exhaustion)**

**Supplementary Methods 19a (Specialty vs Agreeableness vs Personal Accomplishment)**

**Supplementary Methods 19b (Specialty vs Conscientiousness vs Personal Accomplishment)**

**Supplementary Methods 19c (Specialty vs Extraversion vs Personal Accomplishment)**

**Supplementary Methods 19d (Specialty vs Neuroticism vs Personal Accomplishment)**

**Supplementary Methods 19e (Specialty vs Openness vs Personal Accomplishment)**
